# Germline loss-of-function *PAM* variants are enriched in subjects with pituitary hypersecretion

**DOI:** 10.1101/2023.01.20.23284646

**Authors:** Giampaolo Trivellin, Adrian F. Daly, Laura C. Hernández-Ramírez, Elisa Araldi, Christina Tatsi, Ryan K. Dale, Gus Fridell, Arjun Mittal, Fabio R. Faucz, James R. Iben, Tianwei Li, Eleonora Vitali, Stanko S. Stojilkovic, Peter Kamenicky, Chiara Villa, Bertrand Baussart, Prashant Chittiboina, Camilo Toro, William A. Gahl, Erica A. Eugster, Luciana A. Naves, Marie-Lise Jaffrain-Rea, Wouter W. de Herder, Sebastian JCMM Neggers, Patrick Petrossians, Albert Beckers, Andrea G. Lania, Richard E. Mains, Betty A. Eipper, Constantine A. Stratakis

**Author notes:** **Corresponding author and person to whom reprints should be addressed:** Dr. Giampaolo Trivellin, PhD, Department of Biomedical Sciences, Humanitas University, Via Rita Levi Montalcini 4, 20072 Pieve Emanuele – Milan, Italy. **Author contributions:** GT and CAS conceived the study. GT, LCHR, AFD, RKD, GF, AM, BAE, REM, EA, and CAS formulated study hypotheses and conceived and designed the experiments. GT, LCHR, GF, AM, JRI, TL, CT (Camilo Toro), EA, FRF, CT, BAE, and REM performed the experiments. GT, LCHR, AFD, GF, EA, FRF, CT (Camilo Toro), CT (Christina Tatsi), BAE, and REM analyzed the data. GT, AFD, EA, BAE, and REM constructed the figures. GT wrote the original manuscript draft. All authors reviewed and edited the manuscript.

## Abstract

Pituitary adenomas (PAs) are common, usually benign tumors of the anterior pituitary gland which, for the most part, have no known genetic cause. PAs are associated with major clinical effects due to hormonal dysregulation and tumoral impingement on vital brain structures. Following the identification of a loss-of-function variant (p.Arg703Gln) in the *PAM* gene in a family with pituitary gigantism, we investigated 299 individuals with sporadic PAs and 17 familial isolated pituitary adenomas kindreds for *PAM* variants. *PAM* encodes a multifunctional protein responsible for the essential C-terminal amidation of secreted peptides.

Genetic screening was performed by germline and tumor sequencing and germline copy number variation (CNV) analysis. No germline CNVs or somatic single nucleotide variants (SNVs) were identified. We detected seven likely pathogenic heterozygous missense, truncating, and regulatory SNVs. These SNVs were found in sporadic subjects with GH excess (p.Gly552Arg and p.Phe759Ser), pediatric Cushing disease (c.-133T>C and p.His778fs), or with different types of PAs (c.-361G>A, p.Ser539Trp, and p.Asp563Gly). The SNVs were functionally tested *in vitro* for protein expression and trafficking by Western blotting, for splicing by minigene assays, and for amidation activity in cell lysates and serum samples. These analyses confirmed a deleterious effect on protein expression and/or function. By interrogating 200,000 exomes from the UK Biobank, we confirmed a significant association of the *PAM* gene and rare *PAM* SNVs to diagnoses linked to pituitary gland hyperfunction.

Identification of *PAM* as a candidate gene associated with pituitary hypersecretion opens the possibility of developing novel therapeutics based on altering PAM function.

## Introduction

The anterior pituitary gland plays a critical role in the dynamic control of major hormonal systems, including growth, fertility, and stress responses. Anterior pituitary adenomas (PAs), also called pituitary neuroendocrine tumors (PitNETs) [1], can be comprised of any of the secretory cell subtypes, such as lactotropes that secrete prolactin, somatotropes (growth hormone (GH)), corticotropes (adrenocorticotropic hormone (ACTH)), gonadotropes (follicle stimulating hormone (FSH) and luteinizing hormone (LH)), and thyrotropes (thyroid stimulating hormone (TSH)) [2]. Although they are usually benign lesions, PAs can have a major impact through hormonal dysregulation and direct mass effects or invasion of brain structures (optic chiasm, cavernous sinus). Epidemiologically, PAs are one of the most frequent intracranial tumor types, and lead to clinically apparent disease with a frequency of approximately 1 per 1,000 in the general population [3, 4]. Approximately 95% of PAs occur sporadically. Hereditary PAs are often distinguished by a more severe clinical presentation, such as an earlier age at onset, aggressive growth, larger size, and greater resistance to treatment, and they might coexist with other syndromic components. Therefore, identification of germline genetic causes can have implications for earlier identification and treatment of affected individuals via genetic and clinical screening [5].

Germline changes like single nucleotide variants (SNVs) and/or copy number variants (CNVs) in several genes have been implicated in familial isolated pituitary adenomas - FIPA (*AIP* and *GPR101* genes) [6, 7], familial syndromic pituitary adenomas (*MEN1*, *CDKN1B*, *PRKAR1A*, *PRKACB*, *SDHx*, *MAX*, *NF1*, *DICER1*, *TSC2,* among others) and sporadic PAs (*AIP*, *GPR101*, *CABLES1*). Recurrent somatic SNVs are found most frequently in *GNAS* and *USP8*, in sporadic GH and ACTH-secreting PAs, respectively, and rarely in other genes [8]. Despite these advances in establishing gene-disease links, the etiology of the overwhelming majority of inherited and sporadic pituitary adenomas remains unknown. Hence, identification and characterization of novel pathological genetic and genomic variants in PA cohorts is medically significant.

In a large, international cohort of individuals with PAs, we identified germline loss-of-function (LOF) variants of *PAM* (peptidylglycine α-amidating monooxygenase, MIM: 170270) in subjects with familial and sporadic GH and ACTH hypersecretion. PAM is a highly conserved, multifunctional protein that is increasingly recognized as an important regulator of peptide amidation and secretion, among many other functions, in health and disease [9]. We functionally evaluated 36 SNVs, demonstrating a deleterious effect on PAM function/expression in eight variants. We also report a statistically significant association of rare *PAM* SNVs with diagnoses of hyperfunction or tumors of the anterior pituitary gland. These results suggest that pathogenic *PAM* variants can predispose and/or contribute to the development of pathological pituitary hypersecretion.

## Results

### Identification of a pathogenic SNV in the *PAM* gene in a family with pituitary gigantism

The discovery cohort consisted of a three-member, non-consanguineous FIPA family that was enrolled in the UDP of the NIHCC. The kindred consisted of monozygotic twin brothers (individuals II-2 and II-3) and the eldest child of individual II-2, all of whom were affected by childhood-onset pituitary gigantism (Table 1 and Figure 1A).

**Figure 1.**
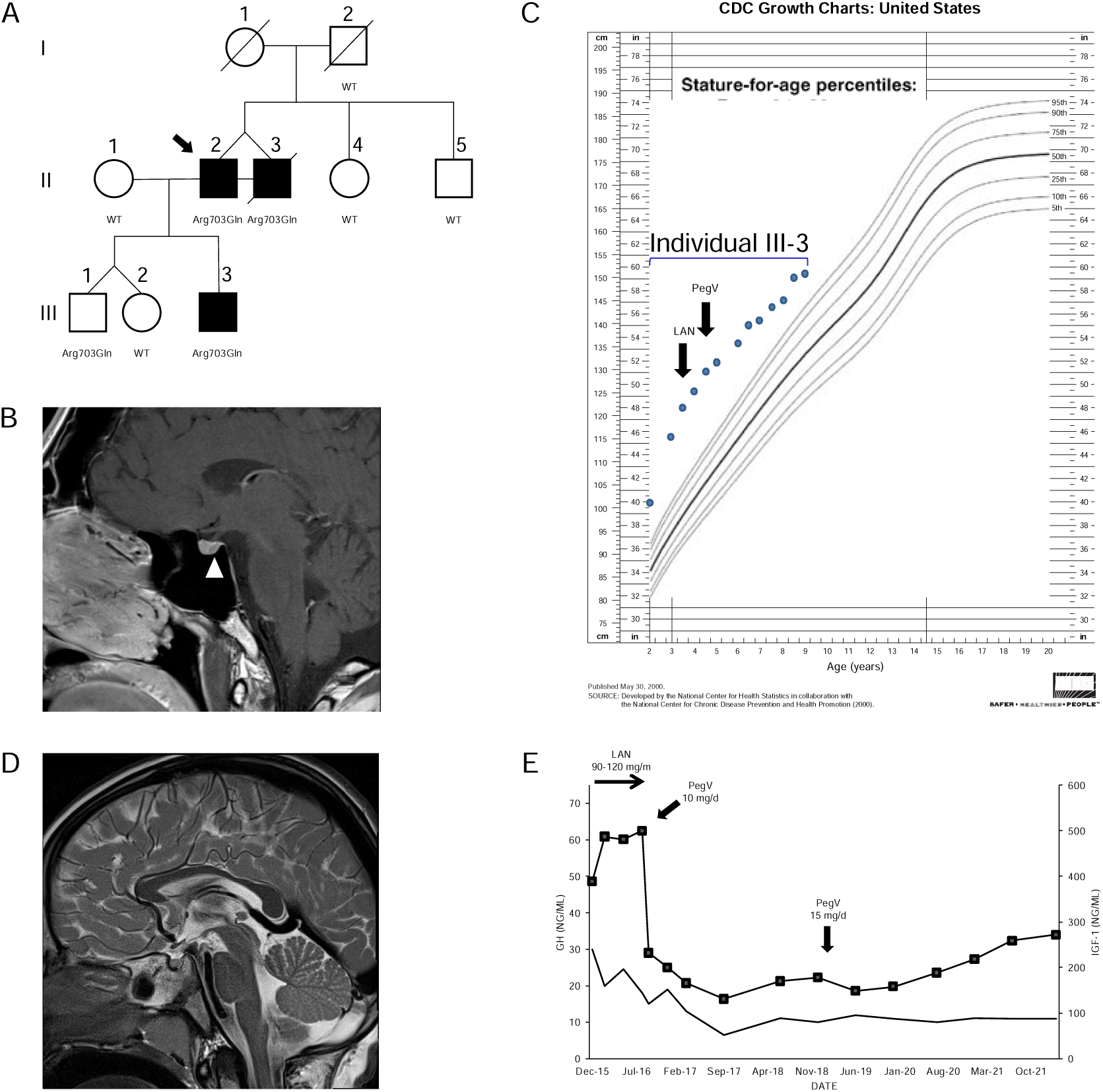
Clinical findings in the index family with pituitary gigantism. (A) Pedigree. Generation numbers are represented by Roman numerals, and individual numbers are in Arabic numerals. The proband is II-2, indicated by the black arrow. Open square/circle, unaffected male/female; filled square, affected male. *PAM* mutational status is shown under each screened individual. In (B) the white arrowhead points to a possible 4 mm lesion, seen in II-2. (C) Growth chart for individual III-3 before and after medical interventions. (D) A sagittal T2-weighted MRI of individual III-3 that was performed before medical therapy began did not reveal a pituitary lesion. (E) Time course of the effects of treatment modalities on GH (left axis) and IGF-1 (right axis) in individual III-3. The rapid decrease of IGF-1 after switching from lanreotide (LAN) to pegvisomant (PegV) is evident.

**Table 1.** Summary of characteristics of seven cases with rare pathogenic PAM variants.

| ID | Diagnosis | Gender | Pathogenic *PAM* variant(s) | Ethnicity | Age at disease onset (years) | Age at PA diagnosis (years) | Tumor size (mm) | Treatment | Other clinical diagnoses |
| --- | --- | --- | --- | --- | --- | --- | --- | --- | --- |
| II-2* | GH excess (gigantism) | M | c.2108G>A (p.Arg703Gln) | Caucasian | <10 | 35-40 | 4 | PegV | hypogonadism, adrenal nodules, pleural masses, severe osteopenia, muscular atrophy, diverticulosis |
| II-3* | GH excess (gigantism) | M | c.2108G>A (p.Arg703Gln) | Caucasian | <10 | 35-40 | 5x6 | TSS, SSA, PV | hypogonadism, pancreatic and colon adenocarcinoma |
| III-3* | GH excess (gigantism) | M | c.2108G>A (p.Arg703Gln) | Caucasian | 1-5 | 1-5 | n.a. | SSA, PegV | hypotonia |
| NIH26 | pediatric CD | F | c.2332-2A>T (p.His778fs) | Caucasian | 10-15 | 10-15 | 3 | TSS | scleroderma |
| NIH36 | pediatric CD | F | c.-133T>C | African-American | 10-15 | 10-15 | 10 | TSS (x3) | no |
| Belgium128 | GH excess (gigantism) | M | c.2276T>C (p.Phe759Ser) | Latino | 15-20 | 20's | Macro | TSS, SRL | none |
| Belgium197 | GH excess (acromegaly) | M | c.1654G>A (p.Gly552Arg), c.1688A>G (p.Asp563Gly) | Caucasian | 40's | 40's | 40 | TSS, SRL, RTx | none |

F, female; M, male; n.a., not applicable; PA, pituitary adenoma; PegV, pegvisomant; RTx, radiotherapy; SRL, somatostatin receptor ligand; TSS, transsphenoidal surgery.
\*Member of the index FIPA kindred.

The monozygotic twins were born at 31 weeks gestation; their birth weights and lengths were 1.96 kg and 48.3 cm (II-2) and 2.3 kg and 53.3 cm (II-3), respectively. They had two normally sized siblings. Their father was 190 cm in height. Their mother was very tall [187 cm; +4.1 SDS using Centers for Disease Control (CDC) charts] but had no known history of pituitary disease or established medical diagnosis of overgrowth; she died of colorectal cancer in her 70’s. The precise onset of the twins’ excessive growth is uncertain, but it began during childhood, such that by in their 10’s they both measured >200 cm in height (>+6.2 SDS). They were formally diagnosed with pituitary gigantism as adults (35-40 years), at which time their final height was 231 cm (+7.8 SDS). At diagnosis they had clinical signs of acromegaly including coarse facial features, frontal bossing, and enlarged extremities. Elevated random GH (26 and 23.1 ng/mL) and IGF-1 (603 and 1130 ng/mL) levels were noted, but prolactin levels were below normal levels. Only minor abnormalities were noted on 3 Tesla pituitary magnetic resonance imaging (MRI), with possible small pituitary microadenomas, 4 mm and 5 mm in maximum diameter (Figure 1B). Individual II-3 underwent transsphenoidal surgery, but no somatotropinoma tissue was identified by histopathology. Both brothers required medical therapy with the GH-receptor antagonist, pegvisomant, to reduce excess IGF-1. Colorectal and pancreatic adenocarcinomas were diagnosed in patient II-3, and he subsequently died of the latter tumor in his 40’s.

The third affected member was III-3, a male (son of II-2) who was born with normal length and weight but developed marked overgrowth by the age of 12 months (Figure 1C). His siblings are dizygotic twins with normal height. III-3 was diagnosed with GH and IGF-1 excess (random GH 30 ng/mL and IGF-1 389 ng/mL) before five years of age. An MRI demonstrated a normal pituitary gland (Figure 1D). To control the excessive growth, the patient was started on the long-acting somatostatin receptor ligand lanreotide autogel at a dose of 90-120 mg/month sc. Despite this adult level dosing, the GH remained uncontrolled and the IGF-1 rose to 487 ng/mL. During the lanreotide autogel treatment period, the growth velocity remained accelerated, at 15.9-20 cm/year. Similar to II-2 and II-3, his condition markedly improved on switching to pegvisomant (10-15 mg/day sc): IGF-1 rapidly fell from 500 ng/mL to 232 ng/mL and growth velocity decreased to 3.6-5.6 cm/year during approximately five years of follow-up. His growth curves for height and weight remain above the 99^th^ centile but are no longer diverging (Figure 1E).

We conducted WES in peripheral blood-derived DNA isolated from the index cases (see the Methods section for details). We applied a variant prioritization strategy (Figure S1 and Table S3) and selected for further analysis a heterozygous missense SNV in *PAM* (c.2108G>A, p.Arg703Gln), which was shared by both twins from generation II (Figure 1A). DNA of their deceased mother was unavailable, so we could not confirm whether they inherited the *PAM* variant from her or if it arose *de novo*; the father was WT. The p.Arg703Gln variant was present in the other individual with gigantism, III-3, but also in III-1, his younger brother (Figure 1A). Endocrine and growth studies showed that III-1 had no evidence of growth excess or hormonal dysregulation at the time of study (age < 10 years). All other family members studied were WT for *PAM* and had normal growth. Overall, these results suggest incomplete penetrance. No other potentially pathogenic alterations in genes were shared among the three affected members.

The *PAM* variant was notable among the 27 candidates due to its strong expression in neuroendocrine tissues and gene function. As reported in *The Human Protein Atlas*, the pituitary is among the tissues displaying the strongest *PAM* mRNA and protein expression signal in human and other mammals. *PAM* is highly expressed in all pituitary cell types, including folliculostellate cells (Figure S2A), where it plays a crucial function in post-translational hormone processing and secretion [10, 11]. Staining for PAM and GM130, a cis-Golgi marker, in normal adult human pituitary cells showed diffuse cytoplasmic expression for PAM that partially overlapped the Golgi (Figure S2B), reflecting the established trafficking of PAM within the secretory pathway observed in rat pituitary cells. PAM is a multifunctional protein that contains two enzymatic domains, peptidylglycine α-hydroxylating monooxygenase (PHM) and peptidyl-α-hydroxyglycine α-amidating lyase (PAL); acting sequentially, the two domains generate C-terminally amidated peptides [10]. The p.Arg703Gln variant – located in the PAL domain (Figure 2A) – has an extremely low minor allele frequency in controls (MAF: 0.0013%), no homozygous variant entries in gnomAD, and is bioinformatically predicted to be pathogenic (Table 2).

**Figure 2.**
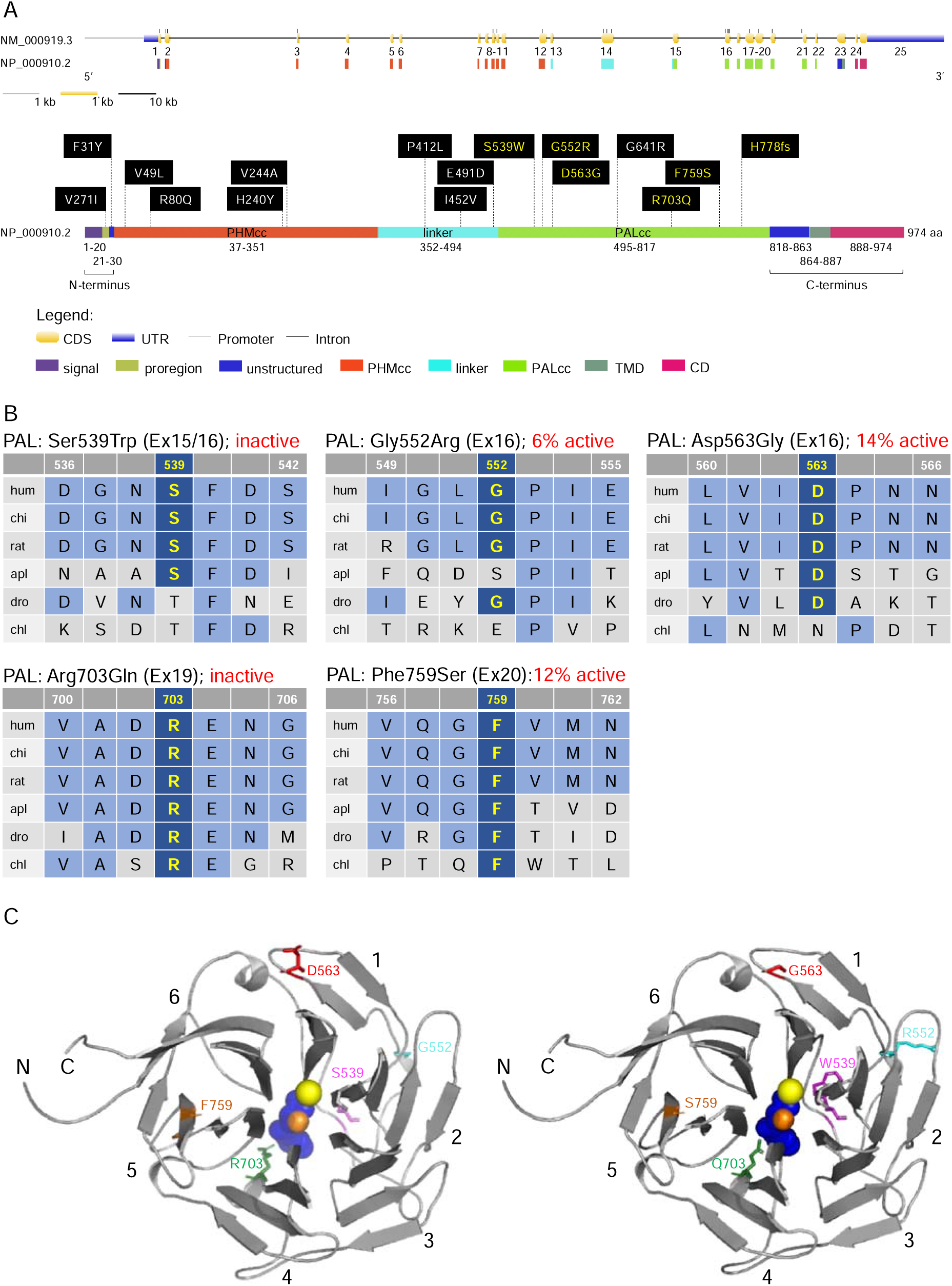
Location and evolutionary conservation of the missense and frameshift *PAM* SNVs that were functionally tested. (A) Schematic representation of the *PAM* gene (GenBank: NM_000919.3, PAM-1, 25 exons) and encoded protein (NP_000910.2, 974 amino acids), including functional domains, with 15 missense and a frameshift SNVs. Gene and protein structures were drawn with the Gene Structure Display Server (GSDS ver. 2.0) [14]. Variants found to have deleterious effects on PAM function/expression (p < 0.01) are shown in yellow lettering, while those without major effects are shown in white lettering. Brackets identify the non-catalytic regions that precede PHMcc (N-Terminus) and follow PALcc (C-Terminus); CD, cytosolic domain; CDS, coding sequence; PALcc, catalytic core of peptidyl-α-hydroxyglycineα-amidating lyase; PHMcc, catalytic core of peptidylglycine α-hydroxylating monooxygenase; TMD, transmembrane domain; UTR, untranslated region. (B) Protein sequence alignments for five of the variants with deleterious effects. Conserved affected residues are shown in yellow. PAL activities, indicated in red, refer to functional experiments in PEAKrapid cells. (C) The crystal structure of rat PALcc (PDB entry 3FW0) was used to contextualize the missense variants categorized as likely pathogenic based on *in silico* analyses; the WT residue is shown on the left and the mutant residue on the right. PAL folds as a β-propeller, with six blades (numbered 1 to 6) positioned around a central cavity. The calcium and mercury ions are depicted as yellow and orange spheres, respectively. The mercury ion was used instead of zinc to capture the binding of a nonpeptide substrate, α-hydroxyhippuric acid, depicted in blue. The affected residues are highlighted in purple (Ser or Trp 539), cyan (Gly or Arg 552), red (Asp or Gly 563), green (Arg or Gln 703), and orange (Phe or Ser 759), along the ribbon visualization of WT rat PALcc in grey. Arg703 is positioned at the active site and participates in substrate binding. Interestingly, p.Gly552Arg and p.Asp563Gly are located on the same face of the β-propeller. C, C-terminus; N, N-terminus.

**Table 2.**
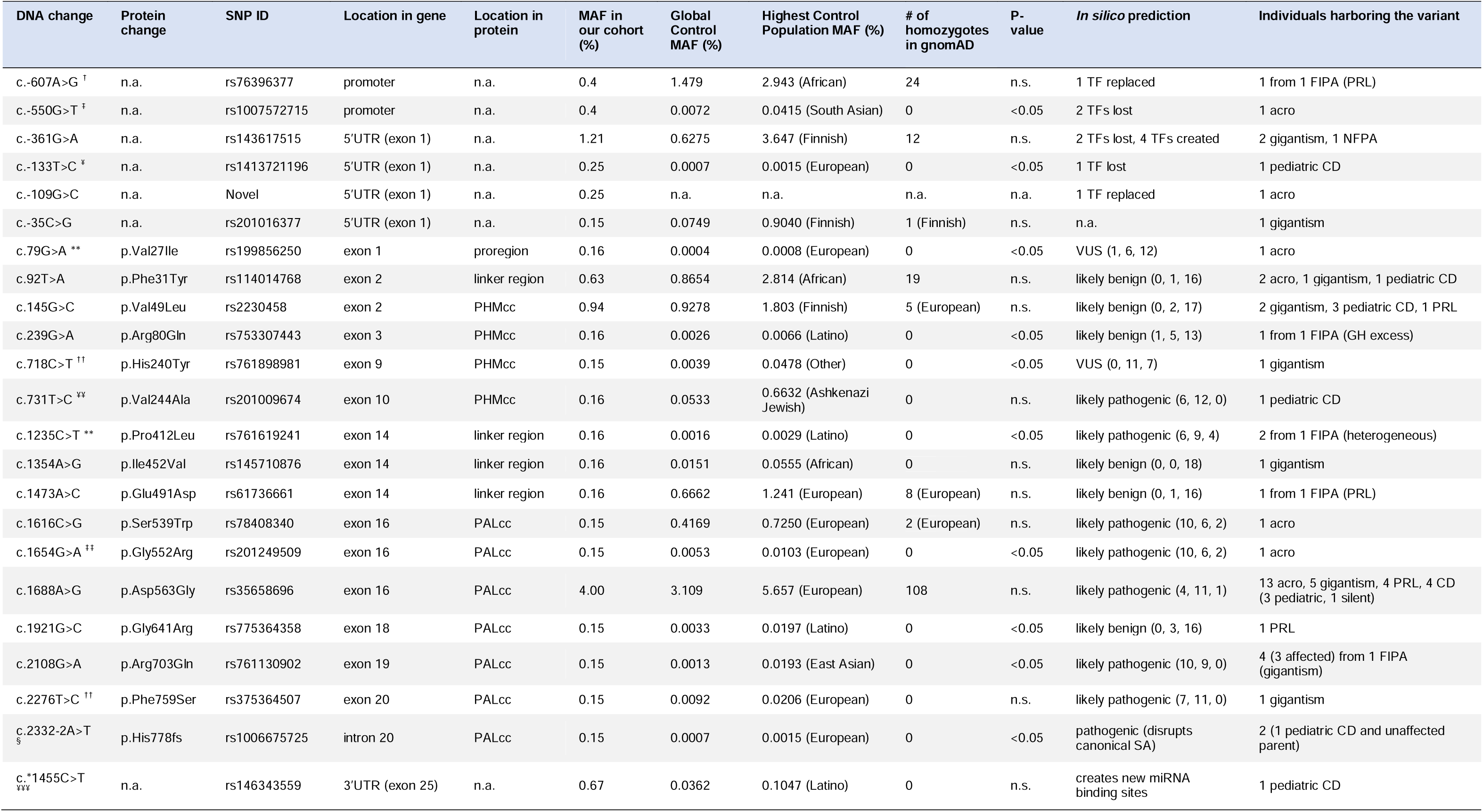
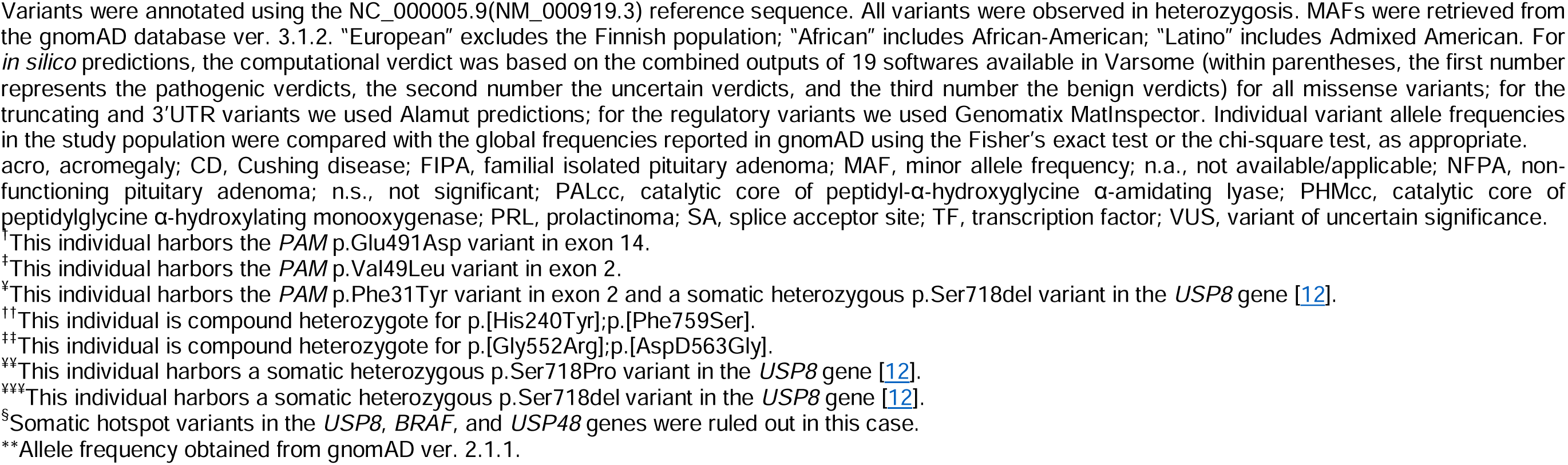
Prioritized heterozygous missense, truncating, and regulatory PAM variants that were functionally analyzed.

Arg703 is conserved throughout evolution (Figure 2B). Along with Tyr651, crystallographic studies identified Arg703 as part of the PALcc catalytic dyad; the catalytic activity of soluble recombinant rat PALcc, in which Arg703 was replaced by Ala or Gln, was greatly reduced [13], a result consistent with its location at the active site (Figure 2C). Altogether, these data suggested that a LOF *PAM* variant could be associated with pituitary gigantism, prompting us to pursue this lead.

### *PAM* screening in a validation cohort revealed multiple likely pathogenic SNVs by *in silico* prediction

We next determined whether predicted pathogenic *PAM* variants are associated with other types of PAs, both hereditary and sporadic. To do so, we screened a diverse group of 326 germline and 60 tumor DNAs from PA patients for *PAM* SNVs (see the Methods section and Table S1 for details). Sequencing covered the *PAM* CDS and exon–intron junctions. Germline CNVs at the *PAM* locus were analyzed by ddPCR in a cohort of 137 individuals with PAs (16 with gigantism and 121 with different types of PAs). Germline DNA sequencing detected 51 SNVs (Table 2 and Table S4). There were no germline CNVs, or rare/pathogenic somatic DNA SNVs (data not shown). A subset of the germline SNVs (15 of 51) was prioritized for further screening together with the p.Arg703Gln variant (Table 2). This selection was based on the following non-mutually exclusive criteria: MAF < 1%, *in silico* analysis with multiple algorithms predicting a deleterious effect on PAM function or splicing, and published functional data indicating pathogenicity [15]. To complement these data, we used Clustal and structure-based analyses to identify variants that could impact protein structure/function (Figure 2B and 2C, Figure S3). The remaining, non-characterized germline SNVs are listed in Table S4.

Four of the 16 heterozygous nonsynonymous and splice site-affecting SNVs prioritized for functional studies were in PHMcc and seven were in PALcc; the remainder were in the non-catalytic regions of PAM (Figure 2A). Based on *in silico* predictions, one of the four SNVs in PHMcc and six of the seven in PALcc were predicted to be pathogenic (Table 2). The prioritized variants were not significantly spatially clustered when we considered the geometric mean distance between all pairs of variants normalized to cDNA length (p = 0.35). However, when we classified the variants based on their location within the N-terminus, PHMcc, linker, PALcc or C-terminus (Figure 2A), the N-terminus harbored more variants than expected by random chance (p = 0.02, Table S5).

### Expression of prioritized *PAM* SNVs revealed deficits in protein expression, enzymatic activity, and glycosylation

To understand the impact of the prioritized *PAM* variants on the function of this membrane enzyme, we transiently expressed each PAM-1 variant in PEAKrapid (HEK-293 derivative) cells. To facilitate accurate assessment of the PHM and PAL activity of each *PAM* variant, a non-ionic detergent (TX-100) was used to solubilize membrane proteins from a crude particulate fraction (Figure 3A). PAM expression levels and protein integrity were evaluated by Western blot analysis (Figure 3B) and PAM trafficking was assessed by analysis of N- and O-linked oligosaccharide maturation (Figure 3C and 3D).

**Figure 3.**
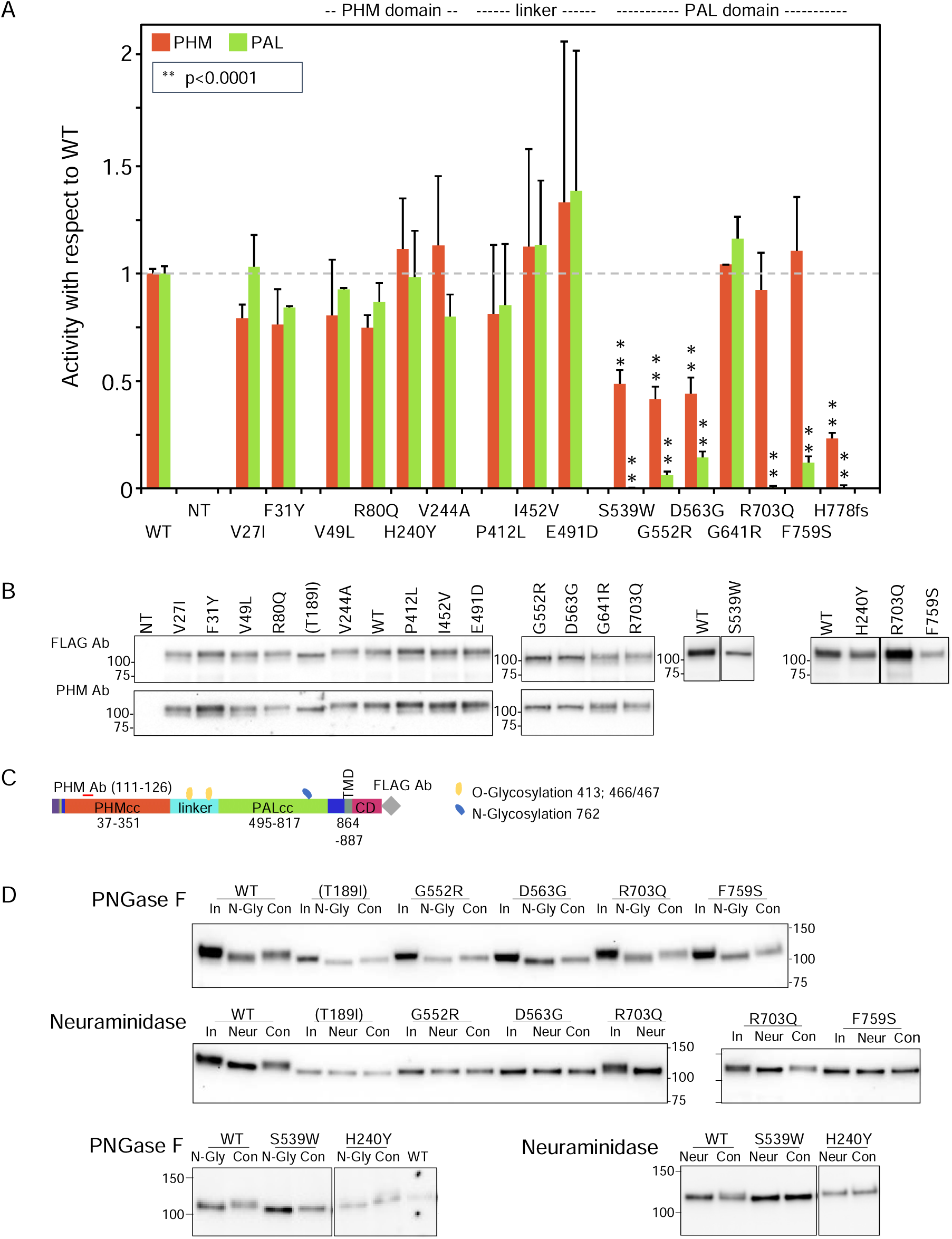
Enzymatic activity, protein expression, and glycosylation pattern of *PAM* variants. (A) PHM and PAL activity. As described in Methods, TMT solubilized particulate fractions prepared from transiently transfected PEAKrapid cells were assayed for PHM activity and for PAL activity. Data for the level of expression of WT PAM and each full-length variant were determined by quantifying the FLAG-tag signal. Levels of p.His778fs were assessed as described in Methods. NT, not transfected; WT, wild-type; **p < 0.0001. (B) PAM protein expression. SNV expression was assessed using a FLAG tag antibody and an antibody to a peptide contained in PHMcc (JH246). The lines separating WT and p.His240Tyr from p.Arg703Gln and p.Phe759Ser indicate that data for two intervening samples were removed. Molecular weight standards are indicated. (C) PAM protein diagram, indicating the location of the JH246 epitope (red horizontal line), the FLAG tag (grey diamond), and the expected location of N- and O-glycans (blue and yellow freeform shapes, respectively). (D) Glycosylation is altered in a subset of PAM variants. Cell lysates were treated with PNGase F or neuraminidase as described in Methods. Proteins were visualized using the FLAG antibody. The samples treated with Neuraminidase were analyzed on two separate gels, with the p.Arg703Gln samples appearing in part on both gels. Con, control; In, Input; N-Gly, PNGase; Neur, neuraminidase; NT, not transfected.

To establish the efficacy of our expression system, we compared the properties of WT PAM to those of missense PAM variants designed to inactivate PHMcc (p.Thr189Ile) or PALcc (p.His529Arg and p.Gly796Glu, Figure S4). PHM and PAL activity measurements for WT PAM and for each engineered control were normalized to the expression of that PAM protein [(PHM or PAL activity)/PAM protein], providing a measure of its specific activity; the PHM and PAL specific activities of each engineered control were then compared to the PHM and PAL specific activities of WT PAM. As expected, the PHM activity of the p.Thr189Ile mutant and the PAL activity of the p.His529Arg and p.Gly796Glu mutants were reduced more than ten-fold compared to WT PAM (Figure S4 and Table 3).

**Table 3.** Summary of effects on enzymatic activity and glycosylation for all *PAM* variants that were functionally tested.

| PAM variant | PHM activity (% WT) | PAL activity (% WT) | PNGaseF (N-sugars) | Neuraminidase (sialic acid) |
| --- | --- | --- | --- | --- |
| WT | 100 ± 2 | 100 ± 3 | yes | yes |
| Val27Ile | 79 ± 6 | 103 ± 15 | (yes) | (yes) |
| Phe31Tyr | 76 ± 16 | 84 ± 1 | (yes) | (yes) |
| Val49Leu | 80 ± 26 | 93 ± 1 | (yes) | (yes) |
| Arg80Gln | 75 ± 6 | 87 ± 9 | (yes) | (yes) |
| Thr189Ile<sup>†</sup> | 6 ± 3** | 23 ± 5** | yes | no |
| His240Tyr | 111 ± 24 | 98 ± 22 | yes | no |
| Val244Ala | 113 ± 32 | 80 ± 10 | (yes) | (yes) |
| Pro412Leu | 81 ± 32 | 85 ± 28 | (yes) | (yes) |
| Ile452Val | 112 ± 45 | 113 ± 30 | (yes) | (yes) |
| Glu491Asp | 133 ± 75 | 138 ± 55 | (yes) | (yes) |
| His529Arg<sup>†</sup> | 41 ± 13 | 7 ± 1** | n.a. | n.a. |
| Ser539Trp | 49 ± 7** | 0 ± 1** | yes | no |
| Gly552Arg | 41 ± 6** | 6 ± 2** | yes | no |
| Asp563Gly | 44 ± 8** | 14 ± 3** | yes | no |
| Gly641Arg | 104 ± 0 | 116 ± 10 | (yes) | (yes) |
| Arg703Gln | 92 ± 17 | 0 ± 1** | yes | yes |
| Phe759Ser | 110 ± 25 | 12 ± 3** | yes | no |
| His778fs | 23 ± 3** | 0 ± 1** | n.a. | n.a. |
| Gly796Glu<sup>†</sup> | 55 ± 8 | 4 ± 1** | (yes) | (yes) |

Values are presented as mean ± SEM.
n.a., not assessed; WT, wild-type; (yes), based on SDS-PAGE profile.
<sup>†</sup>Engineered inactive mutants.
\*\* p < 0.0001.

The structure of bifunctional PAM has not yet been experimentally determined, but for all three engineered variants, inactivating mutations placed into one domain resulted in a modest decrease in the activity of the other domain.

Activity and Western blot data for all studied SNVs are shown in Figure 3A and 3B, respectively. No significant decrease in PHM activity was seen for SNVs p.Val27Ile, p.Phe31Tyr, p.Val49Leu, p.Arg80Gln, p.His240Tyr or p.Val244Ala. The three non-catalytic linker domain SNVs tested exhibited no significant change in PHM or PAL activity and were not extensively assayed. Six of the seven SNVs located in PALcc exhibited a dramatic decrease in PAL activity. While some PALcc variants reduced PHM activity, others did not. Although the PAL activity of p.Arg703Gln was undetectable and the PAL activity of p.Phe759Ser was 12% of WT, their PHM activities were equal to that of WT. In contrast, the dramatic reductions in PAL activity observed for p.Ser539Trp, p.Gly552Arg, p.Asp563Gly, and p.His778fs were accompanied by at least a two-fold decrease in PHM activity. Normal levels of both PHM and PAL activity were detected for p.Gly641Arg.

Maturation of N-linked oligosaccharides is generally completed only when the newly synthesized protein exits the Golgi complex. Sialylation of both the N- and O-linked oligosaccharides attached to rat PAM-1 occurs just before it exits the Golgi complex. The stepwise manner in which the maturation of N- and O-linked glycans occurs often results in glycoprotein heterogeneity, contributing to the diffuse bands observed for WT PAM and many of the prioritized SNVs (Figure 3B, FLAG Ab and PHM Ab). In contrast, we consistently observed a slightly smaller, more compact band for p.Thr189Ile (engineered variant), p.His240Tyr, p.Ser539Trp, p.Gly552Arg, p.Asp563Gly, and p.Phe759Ser.

Since altered access to the enzymes responsible for the maturation of N- and O-linked glycans can reflect altered protein trafficking, we utilized PNGase F to remove N-linked oligosaccharides and neuraminidase to remove sialic acid (Figure 3D). The 2 to 3 kDa reduction in apparent molecular mass caused by PNGase F treatment demonstrated that the engineered control lacking PHM activity (Thr189Ile) and that each of these six variants had been N-glycosylated. As expected, neuraminidase treatment brought about a slight decrease in the apparent molecular mass of WT PAM. Strikingly, neuraminidase treatment failed to reduce the mass of any of these six SNVs, indicating that their glycans had not undergone normal sialylation. Despite this, p.His240Tyr, one of the variants exhibiting altered sialylation, had normal levels of both PHM and PAL activity. Altered trafficking in the secretory and/or endocytic pathway could limit sialylation.

A summary of the effect of each prioritized and engineered SNV on PAM expression, enzymatic activity and oligosaccharide maturation is presented in Table 3.

Three SNVs with diminished PAL activity were found in subjects with GH excess leading to sporadic acromegaly (p.Ser539Trp, p.Gly552Arg) or gigantism (p.Phe759Ser). The relatively common p.Asp563Gly variant (3.11% global MAF) was observed in persons with different types of PAs (Table 2). Interestingly, two males with GH excess – one with acromegaly, and one with gigantism – were compound heterozygotes for p.[Gly552Arg];[Asp563Gly] and p.[His240Tyr];[Phe759Ser], respectively (Figure S5, Table 1). Although neither the PHM nor PAL activity of p.His240Tyr (located in PHMcc) differed from WT, its heterogeneous band pattern and lack of sensitivity to neuraminidase suggest structural alterations sufficient to alter its trafficking (Figures 3A, 3B, and 3D); its significantly higher prevalence in the cohort of subjects with PAs vs. controls, and the absence of homozygous individuals in gnomAD, argue for its classification as a variant of uncertain significance (VUS). Further studies are necessary to establish or disprove pathogenicity.

Three of the 15 SNVs were identified in individuals from FIPA families (one variant in each family, Table 2). SNVs p.Arg80Gln and p.Pro412Leu, which are very rare (MAF: 0.002%), are significantly more prevalent in our cohort vs. controls, and are without homozygous variant entries in gnomAD. The p.Arg80Gln SNV is located within PHMcc, while the p.Pro412Leu and p.Glu491Asp SNVs are within the linker region. The p.Arg80Gln did not segregate with the phenotype in the FIPA kindred and functional studies showed no significant effect of p.Arg80Gln on PHM enzymatic activity. In contrast, the p.Pro412Leu variant segregated with the FIPA phenotype and was seen in both the proband (acromegaly) and the daughter (prolactinoma). Neither of the p.Pro412Leu or p.Glu491Asp variants in the non-enzymatic linker region had a functional effect on PHM or PAL activity (Figure 3A, Table 3). Altogether, combining allele frequencies in our cohort vs. controls, *in silico* predictions, functional data, and segregation analysis, our results indicate that the p.Arg80Gln and p.Glu491Asp variants should be classified as likely benign, while p.Pro412Leu linker region change is a VUS.

### Functional evaluation of a truncating variant associated with Cushing disease

A variant affecting a canonical splice acceptor site (c.2332-2A>T) was present in a pediatric female (NIH26) with ACTH excess leading to Cushing disease (Table 1). Her parent also carried the variant and had evidence of disrupted circadian cortisol rhythm, based on elevated midnight serum cortisol, but did not present clinically overt Cushing disease. The variant was predicted to cause skipping of exon 21, generating a frameshift that introduces a premature stop codon, eliminating the C-terminal region of PALcc (p.His778fs, Table 2 and Figure S6). Skipping of exon 21 was confirmed by testing both the affected subject and parent blood-extracted RNA (Figure 4A and 4B) and by using a minigene splicing assay (Figure 4C). Transient expression of p.His778fs in PEAKrapid cells demonstrated, as expected, that the truncated protein lacked detectable PAL activity; in addition, significantly reduced levels of PHM activity were also observed (Figure 3A, Table 3). Western blotting indicated that the mutant protein is produced but at lower levels than those observed for WT. Moreover, the observed band was smaller than predicted (75 vs. 90 kDa, respectively), suggesting that the mutant might lack some post-translational modifications and/or is cleaved at a pair of basic residues introduced by the frameshift (Figure 4D).

**Figure 4.**
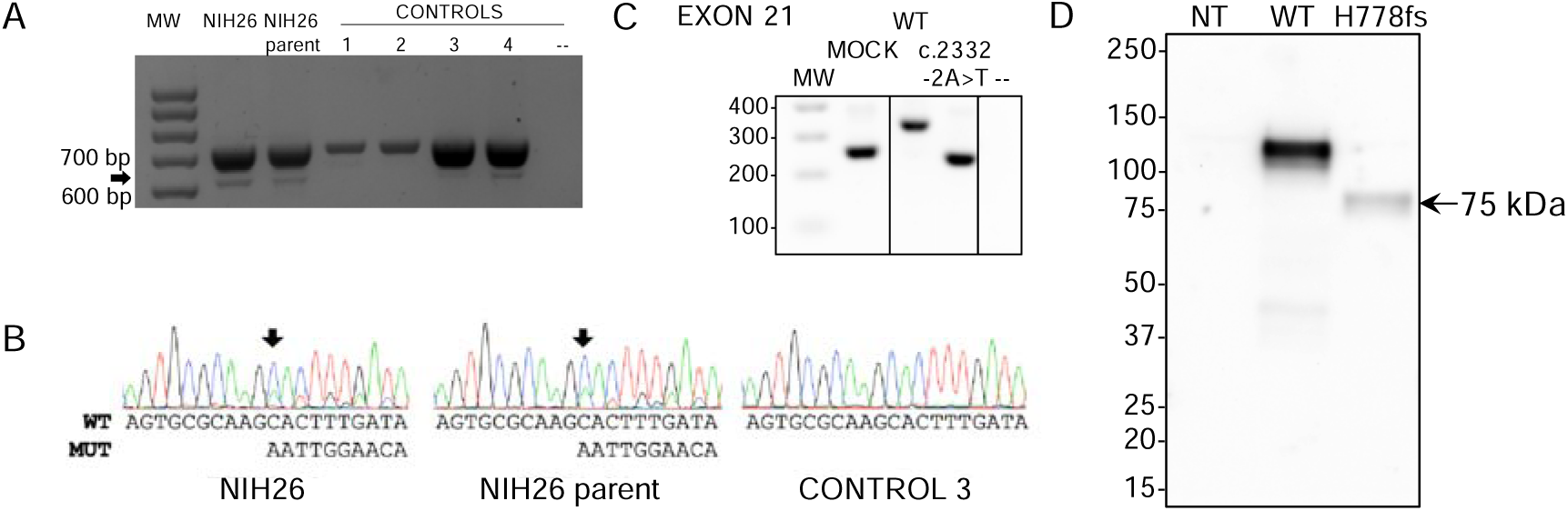
Functional studies of the c.2332-2A>T (p.His778fs) truncating variant. (A) RT-PCR analysis of *PAM* exon 21 splicing was performed using blood-extracted RNA from two family members carrying the c.2332-2A>T variant (NIH26 and parent) and four WT control cDNAs. Primers were designed on exons 17 and 22. Both carriers and controls showed a normally spliced transcript (713 bp, upper band), while only the carriers showed an additional alternatively spliced transcript lacking exon 21 (the 613 bp band, identified by the arrow). MW, molecular weight marker. (B) The identity of the PCR products (panel A) was confirmed by Sanger sequencing. The arrows point to the variant-specific peaks present only in the carriers. MUT, mutated; WT, wild-type. (C) Minigene assay. After transfection into HEK-293 AD cells, mRNA synthesis from the plasmids using the cells’ own transcription and splicing machinery led to mRNA products containing (WT) or lacking (variant) exon 21 of *PAM* flanked by two exons from the pSPL3 vector. The RT-PCR analysis of the minigene transcripts was conducted using vector-specific primers. MW, molecular weight marker; MOCK, cDNA from empty vector-transfected cells consisting of a 260 bp band made up of fragments of pSPL3 exons; --, negative control (RT-PCR without cDNA). The lines separating MOCK and negative control from WT and c.2332-2A>T indicate that data for intervening samples were removed. (D) An expression vector lacking exon 21 of human PAM-1 (H778fs) was transiently expressed in PEAKrapid cells. Proteins were visualized using an antibody to PHMcc (JH246 PHM Ab). Expression of WT PAM produces a major band at 114 ± 1 kDa and a minor one at 105 kDa. The only band visible in the cells expressing p.His778fs migrated at 75 kDa; after signal peptide removal, the mass predicted for this protein – which includes only the first 777 residues of WT PAM-1 but extends 45 residues beyond residue 777 before reaching a stop codon – is 90.31 kDa. NT, not transfected.

### Screening for *PAM* SNVs located in regulatory regions identified two variants that reduce transcription

Next, we extended our screening to regulatory regions of *PAM*, namely the promoter and untranslated regions (UTRs), to investigate whether there are SNVs with the potential to impact *PAM* expression. We identified multiple variants and prioritized seven – two in the promoter, four in the 5’UTR, and one in the 3’UTR – for functional studies (Table 2 and Figure S7). All SNVs in regulatory regions were observed in the heterozygous state. The selection criteria we applied for prioritization were analogous to those used for nonsynonymous variants. *In vitro* evaluation in HEK-293 AD cells using reporter assays, in which we cloned either the promoter-5’UTR upstream or the 3’UTR downstream of the luciferase coding sequence, identified two 5’UTR variants, c.-361G>A and c.-133T>C, that significantly reduced luciferase activity when compared to the WT sequence (Figure 5A). In contrast, the 3’UTR variant c.*1455C>T, which was predicted to create new miRNA binding sites, did not produce any effect on luciferase activity when expressed alongside the identified miRNAs (Figure S8), indicating that it does not impair *PAM* mRNA stability and expression. The c.-361G>A variant was identified in two patients with gigantism and in one individual with a non-functioning PA, while the c.-133T>C variant was present in a pediatric subject who also harbored the p.Phe31Tyr variant and was affected by CD (NIH36, Table 1); this genotype will be identified as c.-133T>C(;)p.Phe31Tyr. Based on *in vitro* enzyme assays, the p.Phe31Tyr missense SNV diminished neither PHM nor PAL activity to a significant extent (Figure 3A and Table 3).

**Figure 5.**
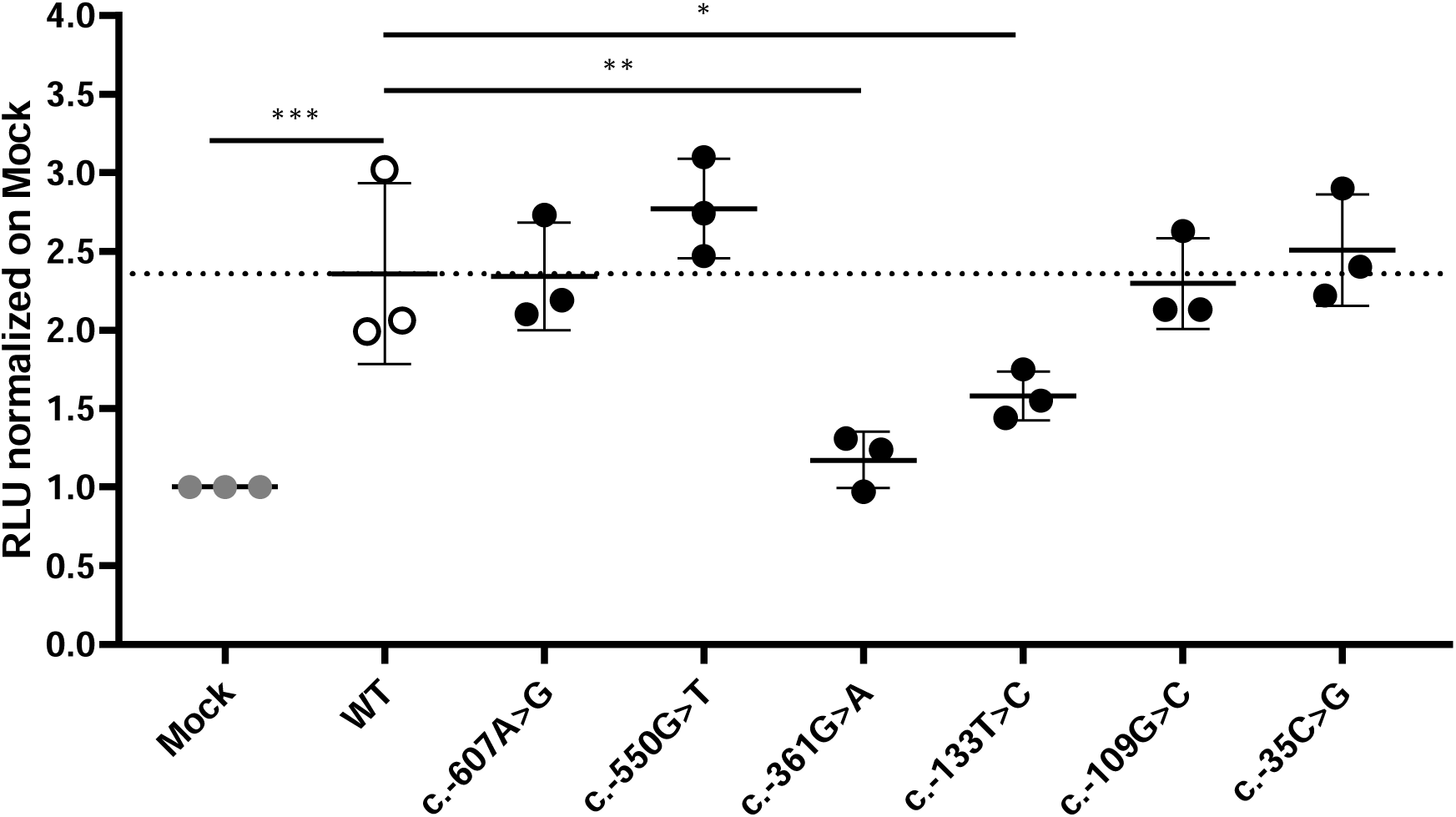
WT and mutant *PAM* promoter activity. A 5 kb *PAM* promoter-5’UTR WT sequence was cloned upstream of a luciferase reporter. Six mutant constructs were created by site-directed mutagenesis. The constructs were transiently transfected into HEK-293 AD cells together with a *Renilla* luciferase reporter for normalization. *Firefly* and *Renilla* luciferase activities were measured 24 h post-transfection. Two SNVs, c.-361G>A and c.-133T>C, have significantly lower transcriptional activity than the WT hybrid transcript. Mock, empty pRMT-Luc vector. Differences between experimental groups were analyzed by 1-way ANOVA with Dunnett’s post hoc test, using WT as the control group. RLU, relative luciferase activity. *, p = 0.0443; **, p = 0.0019; ***, p = 0.0005.

### Quantification of PHM and PAL catalytic activities in serum

Endoproteolytic cleavage of PAM-1 can occur within the regulated secretory pathway and after the endocytic retrieval of PAM-1 from the cell surface [16–18]. In addition, several *PAM* splice variants encode soluble bifunctional PAM proteins that are secreted along with the peptide hormones stored in secretory granules [10]. As a result, both PHM and PAL activity are readily detected in human serum [19, 20]. The anti-coagulants added during the collection of plasma make the accurate assessment of PHM activity in plasma very difficult. As a result, we could only assess PHM and PAL activity in the small number of serum samples collected from our cohort. As shown in Figure 6, we measured PHM and PAL activity and calculated the ratio of PAL to PHM activity for 13 subjects (11 affected individuals/carriers and two WT controls). To control for the fact that the serum samples assayed had been stored for varying periods of time, we evaluated the statistical significance of changes in activity by normalizing PAL activity to PHM activity.

**Figure 6.**
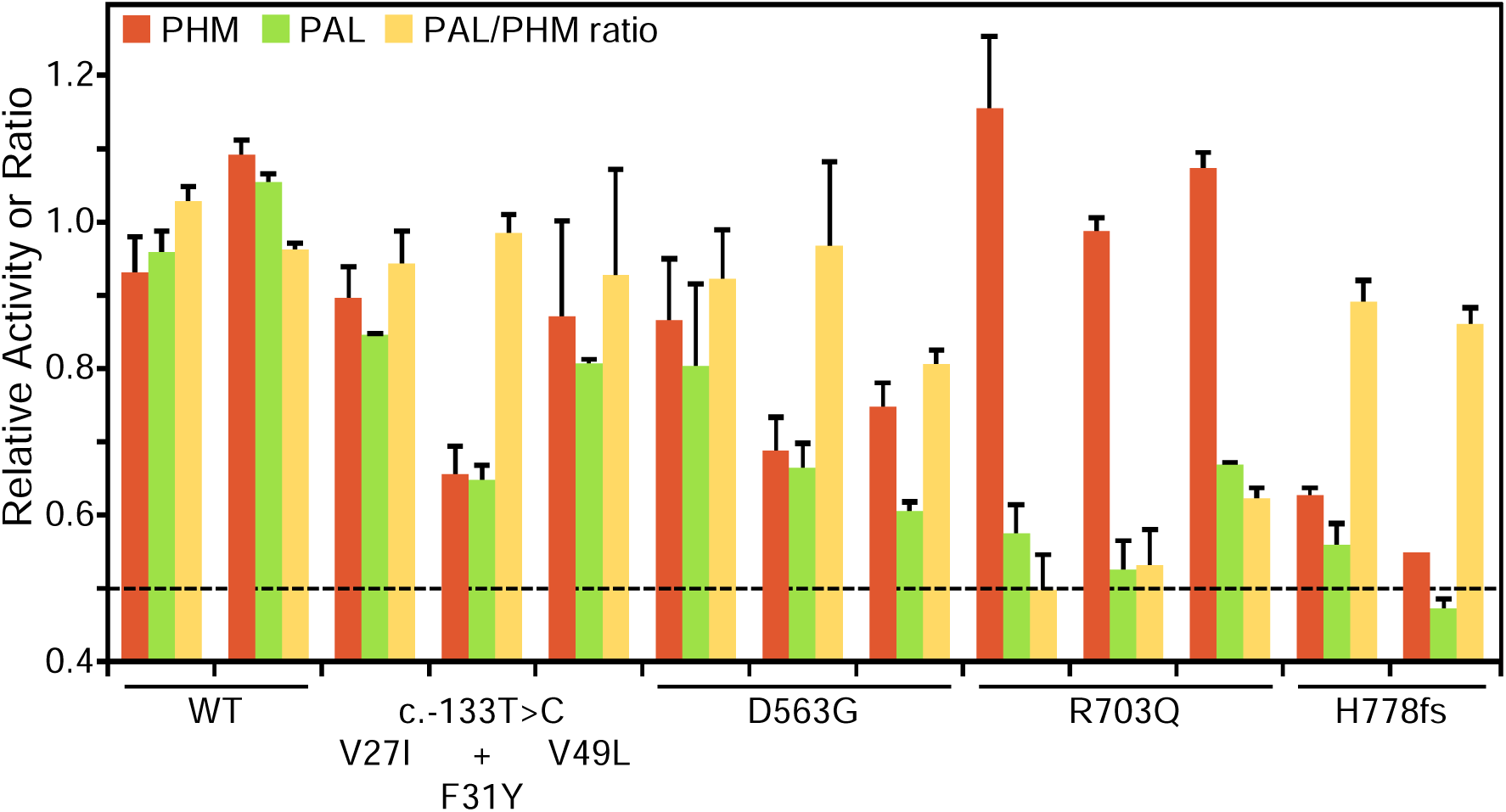
PHM and PAL activity in human sera. Assays for PHM and PAL activity were carried out on sera from individuals harboring variants showing deleterious effects in *in vitro* assays (p.Asp563Gly, p.Arg703Gln, and p.His778fs), subjects with more benign variants (p.Val27Ile and p.Val49Leu) and controls (WT). In NIH36, the p.Phe31Tyr variant occurs along with the 5’UTR c.-133T>C variant. The dashed line at 0.5 indicates the activity level expected with one completely inactive allele. WT, wild-type.

Several serum samples were available from patients with two SNVs that resulted in the almost total loss of the PAL activity of PAM-1, p.Arg703Gln (three samples) and p.His778fs (two samples); consistent with our *in vitro* activity assays of transiently expressed PAM-1 and the total elimination of one allele, serum PAL activity fell by a factor of two for both variants. Also consistent with the data shown in Table 3, serum PHM activity was unaffected in the p.Arg703Gln variant and was reduced substantially in the p.His778fs variant. Three of the serum samples available came from subjects carrying SNVs located in PHMcc, but none significantly reduced the PHM or PAL activity of transiently expressed PAM-1. Strikingly, both PHM and PAL activities were reduced in the serum sample from NIH36, who harbors an expression-inhibiting 5’UTR SNV (c.133T>C) in addition to the p.Phe31Tyr SNV. Taken together, our data indicate that the observed reduction in serum PHM and PAL activity can be attributed to the 5’UTR SNV, thus confirming the *in vitro* finding. Sera from three different subjects harboring the p.Asp563Gly variant showed substantial, but variable, reductions in both PHM and PAL activity. A better interpretation of serum PHM and PAL activity requires further insight into its multiple potential sources. Altogether, analysis of the limited set of serum samples available emphasizes the utility of using a wide variety of *in vitro* systems to evaluate the functional effects of PAM SNVs.

### Loss of heterozygosity (LOH) studies

Next, we explored whether PAM acts as a tumor suppressor gene and requires a second somatic hit affecting the WT allele, as would be expected by the Knudson two-hit hypothesis [21]. Therefore, we extracted genomic DNA from the PAs surgically removed from individuals harboring variants with demonstrated deleterious effects and studied LOH by Sanger sequencing. The available tumors that we analyzed harbored the following variants: c.-133T>C(;)p.Phe31Tyr, p.Ser539Trp, p.[Gly552Arg];[Asp563Gly], and p.Asp563Gly (n = 1 tested for each SNV). We did not identify loss of the WT allele, suggesting that *PAM* does not behave like a typical tumor suppressor but may rather be a haploinsufficient gene. Unexpectedly, in the individual compound heterozygous for p.[Gly552Arg];[Asp563Gly], we observed loss of the mutant allele causing the change at codon 552 (Figure S9). This likely represents an instance of *in vivo* reversion to normal of an inherited variant (revertant mosaicism) [22].

### *PAM* SNVs are significantly associated with a hyperfunctioning pituitary gland

Having detected multiple SNVs with deleterious effects on PAM protein function in our cohort, we decided to test whether SNVs within the *PAM* locus are statistically associated with a pituitary disease phenotype in a larger and more heterogeneous cohort. To this end, we interrogated the UKBB, a powerful resource for genetic association analyses that contains exome sequencing data of 200,643 adult individuals with different and well-annotated pathologies. First, we performed a gene-based association study using the sequence kernel association test (SKAT) [23, 24]. For this analysis, we considered ICD-10 codes associated with sellar lesions (D352, E220, E221, E222, E229, E237, E240, covering the following conditions: benign neoplasm of pituitary gland; acromegaly and pituitary gigantism; hyperprolactinemia; syndrome of inappropriate secretion of antidiuretic hormone; hyperfunction of pituitary gland, unspecified; disorder of pituitary gland, unspecified; and pituitary-dependent Cushing’s disease, respectively) and SNVs at the *PAM* locus for which we assigned a weight based on their detrimental effect on protein (see Methods for details). We set a MAF cutoff of 0.1% for rare variants. *PAM* was identified as significantly associated (exome-wide) with a diagnosis of hyperfunction of the pituitary gland (E229, p = 1.71×10^-77^, Figure 7A). Next, we performed single-variant analyses restricted to rare (MAF < 1%) *PAM* variants with a predicted deleterious effect (CADD_PHRED score ≥15). We identified nine missense heterozygous variants (Figure 7B and Figure S10, Table S6), four in PHMcc (p.Ser97Arg, p.Arg190His, p.Pro192Arg, p.Val351Met), four in PALcc (p.Gly531Cys, p.Ile554Val, p.Val597Met, p.Pro693Leu) and one in the C-terminal non-catalytic region (p.Leu856Pro). The p.Ser97Arg variant is located in the loop connecting the β-strand containing the vicinal His residues that bind Cu_H_ to the preceding β-strand and the p.Arg190His and p.Pro192Arg variants are both in the short linker that connects the two homologous domains of PHMcc. PHMcc terminates with Val351; although the final three C-terminal amino acids are not identified in the crystal structure, removal of four C-terminal residues produces a protein lacking PHM activity. Two of the PAL variants (p.Gly531Cys and p.Ile554Val) are situated in or near the long loop that forms its hydrophobic substrate binding pocket and connects β-strands 2 and 3 of Blade 1. The other two PAL variants (p.Val597Met and p.Pro693Leu) are contained within the structural repeats that form Blades 2 and 4. A study aimed at understanding the effects of luminal pH on the trafficking of rat PAM identified the juxtamembrane region that encompasses p.Leu856Pro as a key determinant [25].

**Figure 7.**
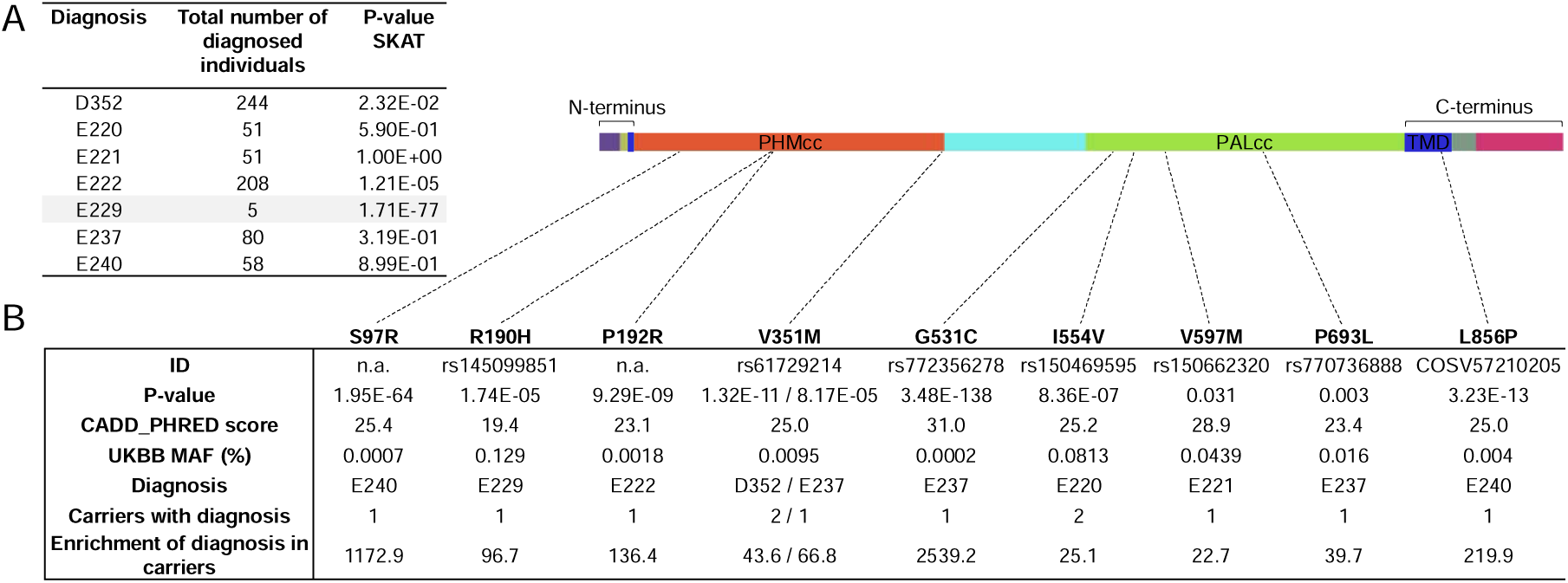
Gene- and variant-based association analyses for *PAM* in the UKBB. (A) Results of SKAT analysis of *PAM* variants for diagnoses of hyperfunction of the pituitary gland (identified from UKBB fields 41270 and 41204 - primary and secondary ICD-10 diagnoses from hospitalization records - and 20002 - self-reported diagnosis) in 200,000 UKBB participants. The SKAT CommonRare algorithm was used for analysis. (B) Significant enrichment for *PAM* pathological missense *PAM* variants in subjects diagnosed with hyperfunction of the pituitary gland.

To complement the analyses conducted on the UKBB, we examined the outputs of a gene-specific metric, the gene damage index (GDI) [26]. The GDI score predicts the liability of a gene to contain disease-causing mutations by considering the influences of selection and genetic drift [27]. For this analysis, we considered 20 genes that harbor germline or somatic variants known to predispose to PAs in order to estimate a GDI cutoff above which a gene is unlikely to cause a pituitary disease, i.e., it is considered a false positive. *PAM* was reported to have a Phred-scaled GDI score of 3.98, below the calculated cutoff of 4.84 (Table S7). Therefore, our analysis indicates an intolerance to mutational changes for *PAM* in large population samples, supporting the notion that the observed LOF variants are more likely to contribute to disease.

### Functional analysis of splicing in other rare variants

Among the rare SNVs identified in the UKBB, an intronic SNV (c.2746+3A>T) was predicted to disrupt the canonical splice donor site of intron 24 (Figure S11A and Table S6); this SNV was associated with a diagnosis of acromegaly and pituitary gigantism (E220). Our splicing analysis ascertained that this SNV caused skipping of exon 24 in most transcript molecules; the expected outcome is the in-frame deletion of 19 residues located within the cytoplasmic domain (p.(Ala897_Arg915del)). While the absence of exon 24 does not alter the PHM or PAL activity of PAM, previous studies indicate that tissue-specific splicing at this site occurs in the rat and could affect the trafficking of membrane PAM [28–30]. PAM mRNA expression levels for the mutant allele were about half of WT levels in *in vitro* studies (Figure S11B). Therefore, an individual harboring this variant is predicted to make 25% less biologically active PAM protein than healthy controls; if trafficked correctly, a reduced level of this variant might support normal PAM functions.

The finding of two non-coding SNVs affecting *PAM* splicing (albeit with different outcomes) and recent studies underscoring the importance of evaluating splicing defects as a disease mechanism for both conditions of hyper and hypopituitarism [31, 32], prompted us to extend the corresponding functional studies to selected synonymous and intronic variants found in our cohort (Figure S12A and Table S8). The selection criteria were analogous to those applied for other prioritized variants. This analysis did not reveal any splicing impairment. We observed only a possible reduction in mRNA expression by the c.195G>T allele (Figure S12B), but this was not subsequently confirmed by qPCR studies (Figure S12C).

Taken together, these findings indicate that impairment of PAM function or expression via abnormal splicing is not frequently associated with PAs.

## Discussion

Studies over the last 30 years have identified a small number of germline and somatic genetic defects associated with the development of different types of PAs. However, the majority of tumors still do not have a known genetic cause [8]. Here, we report that several rare variants in the *PAM* gene are associated with pituitary gland hypersecretion and that most have a deleterious effect on protein function.

We initially identified a deleterious missense *PAM* variant (p.Arg703Gln) in a FIPA family with a severe form of childhood-onset pituitary gigantism in which other established causes, such as X-linked acrogigantism (X-LAG) due to duplications involving *GPR101*, germline *AIP* mutations/deletions or others, had been ruled out [33, 34]. Among the genes filtered from the WES screening, the high level of *PAM* expression in the pituitary gland and its known functions in the pituitary and other endocrine cells made *PAM* the prioritized candidate. A bifunctional enzyme that is essential for the biosynthesis of multiple pituitary and hypothalamic peptide hormones, PAM affects regulated secretion and is involved in the biogenesis of secretory granules [10, 11, 35, 36]. Based on site-directed mutagenesis and crystallographic studies of rat PALcc, this Arg residue (Arg 706 in rat PAM) is located at the active site and participates in substrate binding; site-directed mutagenesis of this residue to Gln reduced the activity of PALcc by a factor of four, and mutagenesis to Ala decreased activity by 97% versus WT [13]. Or functional studies on bifunctional human PAM with the Arg703 to Gln mutation revealed an even greater decrease in its PAL activity (1% relative to WT human PAM), while retaining fully normal PHM activity.

This intriguing finding prompted us to explore whether LOF *PAM* variants (SNVs and CNVs) were associated with other types of anterior pituitary adenomas, occurring both in familial and sporadic settings. No other likely deleterious *PAM* SNVs were observed in the familial cases examined, making the estimated frequency of *PAM* LOF variants in our FIPA kindreds 5.56% (1/18). A very rare VUS, p.Pro412Leu, located within the linker region connecting the two enzymatic domains was identified in a heterogeneous FIPA kindred.

We detected six germline SNVs with deleterious effects on protein function/expression in sporadic PA cases. Two variants (p.Ser539Trp, p.Asp563Gly) are relatively common in the general population (global MAF range: 0.42-3.11%). The remaining four variants are very rare (MAF < 0.01%), were found in one subject each, and were significantly enriched in our cohort compared to the general population. Two of these variants (p.Gly552Arg and p.Phe759Ser) were identified in subjects with GH excess (2/173, frequency: 1.16%) and two (c.-133T>C and p.His778fs) in cases with pediatric CD (2/81, frequency: 2.47%).

The p.Ser539Trp and p.Asp563Gly mutants were previously implicated as major risk factors for type 2 diabetes [37, 38], and an important question is whether these proteins are enzymatically active. When introduced into human PAM-1 and assayed in cell lysates, the p.Ser539Trp variant had 50% of normal PHM activity, lacked detectable PAL activity, and was not normally sialylated, suggesting its abnormal intracellular trafficking. When expressed as a soluble protein containing both PHMcc and PALcc, secretion of the p.Ser539Trp protein was not detected, precluding assessment of its catalytic activity [15]. We prepared two independent p.Ser539Trp plasmid constructs, sequenced both and verified that both constructs were correct. The p.Ser539Trp mutation disrupted the normal trafficking of membrane PAM; when present in the soluble protein, the same variant may inhibit its secretion [15]. Using an assay to quantify PAM activity (PHM followed by PAL) by measuring glyoxylate production from hippuric acid, *Thomsen et al.* reported that the p.Asp563Gly mutation was about 50% active [15]. Our assays, which assess PHM and PAL activity separately, showed that the PHM activity of p.Asp563Gly was slightly less than half that of WT PAM, while its PAL activity was only one seventh that of WT PAM.

Our *in vitro* functional tests indicated an impairment in enzymatic activity for five missense SNVs and the splicing variant (in PAL). *PAM* expression was decreased for two 5’UTR variants and the COOH-terminal region splicing variant. In addition, glycosylation was abnormal for four of the five missense SNVs. Experimental truncations used to define the C-terminus of rat PALcc are consistent with the lack of PAL activity observed for p.His778fs, with the same inability of the truncated rat protein to fold correctly, thereby preventing its secretion [39]. Our functional data indicate a similar behavior for the human variant, along with a significant reduction in PHM activity.

We also showed that measurement of PHM and PAL activity in the serum of subjects harboring *PAM* variants correlates well with the corresponding values for each single variant determined using the PEAKrapid assay system. The CD case that harbored a regulatory variant reducing PAM expression was instructive in this regard. In that case, both PHM and PAL activities were lowered to the same extent (about 40% compared to WT), with the PAL to PHM ratio showing a normal value, suggesting that enzymatic activity was not affected. These observations indicate that it is valuable to explore whether assays of PHM and PAL activity in sera could be used as a biomarker of pituitary disease. The recent development of an efficient high-throughput amidation assay [20] could aid in this endeavor.

The association of loss-of-function PAM SNVs with PAs was further explored by screening 200,000 exomes from the UKBB. Supporting our findings, *PAM* was significantly associated with a diagnosis of pituitary hyperfunction (ICD-10 code E229) and nine rare (MAF < 1%) missense variants, all predicted to be pathogenic, were associated with diagnoses of sellar lesions. Although the UKBB cohort has an inherent selection bias towards healthy volunteers, and disease annotation may be self-reported in some cases or only after hospital admission in others (thereby potentially excluding mild or undiagnosed pituitary hypersecretion diagnoses), the validation of our findings related to *PAM* variants in an independent cohort more representative of the general population strengthens the observed associations. Based on previous studies, each of the nine *PAM* variants identified in the UKBB (four in PHMcc, four in PALcc, and one in the juxtamembrane region of the luminal domain) is expected to disrupt enzymatic activity or trafficking of membrane PAM bearing these mutations.

We did not observe LOH for *PAM* variants. This suggests that *PAM* does not behave as a classical tumor suppressor gene, leading us to propose that pathogenic LOF *PAM* variants act as haploinsufficient alleles. This hypothesis is supported by *in silico* predictions (HI and HIPred scores for *PAM* are 0.588 and 0.7057, respectively) [40, 41], and by previous observations that *Pam* haploinsufficiency impairs several interdependent organ systems in PAM^+/-^ mice. Although a complete lack of *Pam* is not compatible with life [42], mice with a single functional copy of *Pam* developed increased adiposity and mild glucose intolerance with age [42], were deficient in their ability to maintain body temperature in the cold [43], and showed an increase in anxiety-like behavior [44]; these findings are consistent with defects in β-cell function, the vascular system, and neuronal/neuroendocrine function, respectively. Production of the few amidated peptides tested was not greatly altered by PAM heterozygosity [42, 45, 46], but the production of amidated TRH in the hypothalamus was compromised and the mice were deficient in their pituitary response to hypothyroidism [43]. Moreover, pituitary levels of POMC-derived amidated peptides (diAc- α-MSH, α-MSH, and JP) were significantly increased in PAM^+/-^ mice [46]. These data, together with our observation that rare and functionally deleterious *PAM* SNVs are associated with anterior pituitary hyperfunction, suggest that the pituitary gland is particularly sensitive to PAM haploinsufficiency. Moreover, they indicate that changes in gene expression associated with PAM heterozygosity are tissue-specific and that both amidation-dependent and amidation-independent mechanisms (for instance, the secretagogue-mediated translocation of the soluble cytoplasmic PAM fragment to the nucleus affecting gene expression in a cell-type specific manner) [11, 47, 48] likely underlie the described deficits. It will be interesting to study in further detail whether any of the phenotypes observed in PAM^+/-^ mice are also present in individuals with PAs that harbor LOF *PAM* variants.

Limitations to our current study include the fact that only a few subjects with sporadic non-functioning tumors and families with multiple endocrine neoplasia were screened. To better estimate the prevalence of *PAM* variants and their possible contribution to pituitary tumorigenesis, the search for *PAM* variants needs to be extended to those subtypes as well as cohorts with hyperprolactinemia (tumoral or non-tumoral), the most common form of pituitary hypersecretion. In particular, it would be interesting to better assess the risk that the relatively common p.Asp563Gly variant confers to the development of pituitary hyperfunction, particularly in the homozygous state. The variable PAL activity measured in PA subjects that were heterozygous for this variant suggests that other factors, such as genetic, biochemical or dietary (e.g., copper or vitamin C availability, hypoxia) [19], contribute to the regulation of PAM expression and/or activity and, hence, predisposition to pituitary dysfunction and potentially tumorigenesis. Second, our LOH and serum activity analyses were limited to the small number of available PAs and serum samples, respectively. It will be important to screen more tumors harboring deleterious *PAM* variants to definitively exclude the absence of LOH. Likewise, assessing circulating PHM and PAL activity in more *PAM* variant carriers will better define their risk profile. Third, the question of how disordered PAM function leads to increased hormonal secretion and/or pituitary tumor development remains to be determined. PAM is a complex, multifunctional protein involved in several pathways related to the regulation of secretion; disruption of one or more of these pathways could play a role in hormonal hypersecretion (Figure 8).

**Figure 8.**
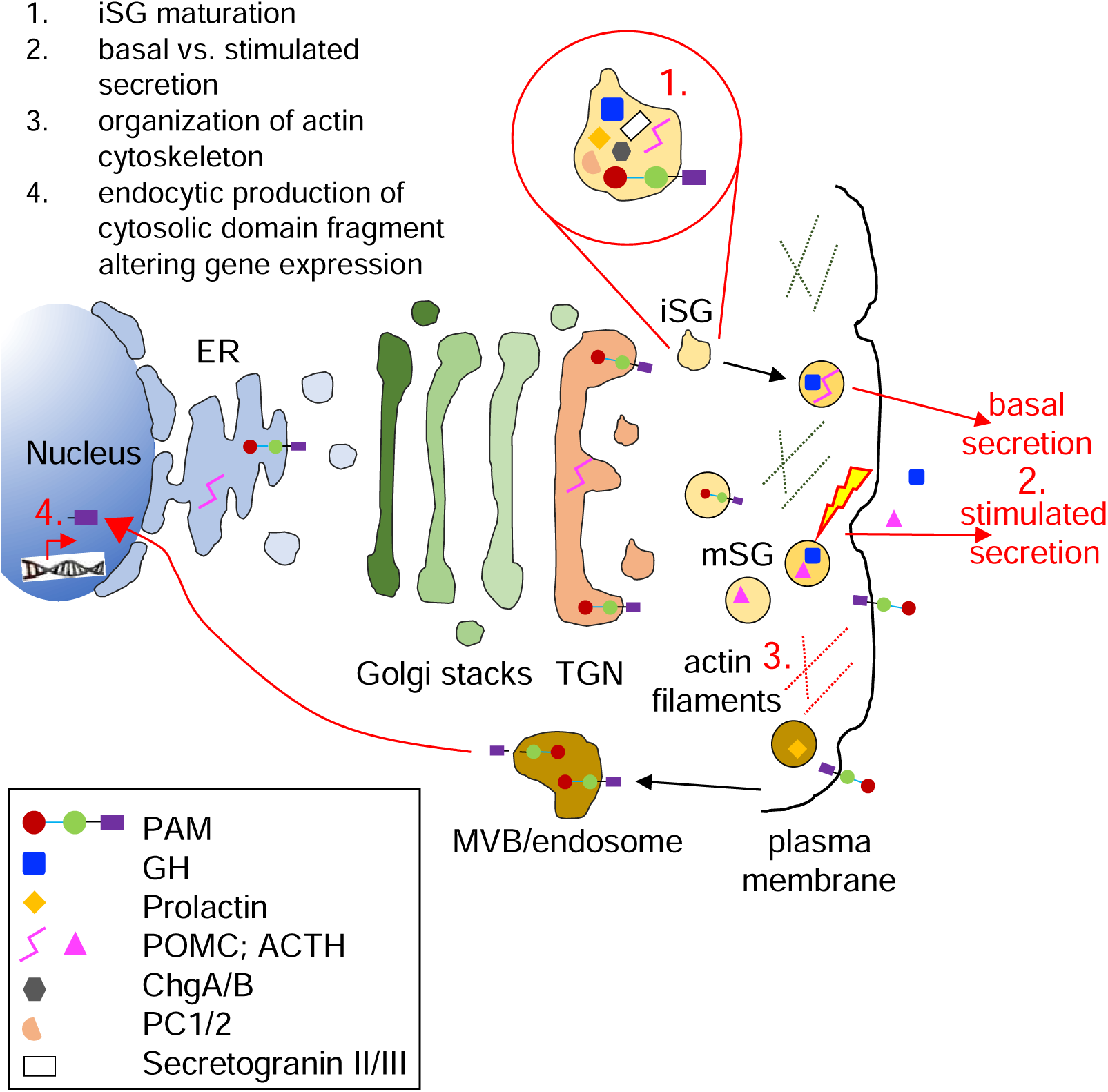
Biological processes and cellular components that might be affected by PAM haploinsufficiency in pituitary hormone-secreting cells. The trafficking of integral membrane PAM and pituitary hormones through the regulated secretory pathway is depicted. Immature secretory granules (iSG) budding from the *trans*-Golgi network (TGN) contain prohormones and processing enzymes like the prohormone convertases (PCs), granins [chromogranins A and B (ChgA/B) and secretogranins II and II], and PAM. Mature secretory granules (mSG) release their soluble content during regulated exocytosis (the yellow lightning bolt represents the external stimulus triggering secretion). Basal rates of hormone secretion from pituitary cells are less than 1% of the cell content/hour. Although PAM appears on the cell surface during exocytosis, its rapid endocytosis means that very little PAM remains on the plasma membrane at steady-state. Cleavage by secretase-like enzymes can generate a soluble, cytosolic fragment of PAM that can enter the nucleus, where it relays information about the status of the secretory granule pool. Several *in vitro* studies have shown that PAM misexpression can affect a variety of steps in the regulated secretory pathway; these are highlighted in red in the cartoon and include SG formation, basal and stimulated secretion, actin cytoskeleton organization, and gene expression. ER, endoplasmic reticulum; GH, growth hormone; MVB, multivesicular body.

In our study pathological PAM variants were associated with hypersecretion of both amidated and non-amidated pituitary hormones, which suggests that the altered PAM function may affect a different target than the hormones themselves. Granin proteins, including chromogranin A and B are crucial factors in hormonal secretion. Chromogranin A-derived peptides such as pancreastatin are themselves targets for amidation by PAM [49], such that changes in PHM and PAL function could alter the packaging and release of hormones contained in secretory granules. Separately, the linker region connecting the PHM and PAL enzymes is O-glycosylated and pH sensitive and is known to influence PAM function and its effects on vesicular trafficking [50].

In AtT-20 cells, expression of the PAM-1 isoform without soluble PHM and PAL leads to accumulation of uncleaved POMC in the trans-Golgi network; however, while PAM-1 expression decreased secretagogue-stimulated secretion by mature granules, it increased basal secretion of larger POMC peptides [35]. More recently, it has been shown that in atrial myocytes proANP storage is a highly PAM-dependent process even though neither proANP nor ANP itself are amidated proteins. Also, in murine atrial cells devoid of PAM that have depleted proANP storage, the reintroduction of PAM facilitated proANP storage. Crucially, this effect occurred irrespective of whether normal or enzymatically inactivated PAM was used [51, 52]. Interestingly, the depletion of proANP in the absence of PAM was unaccompanied by decreased proANP mRNA or protein biosynthesis, but rather occurred due to 3-fold increased basal ANP secretion. Bäck, Mains & Eipper recently suggested that PAM may also have a role as a cargo receptor that directly binds to and protects proANP soon after its synthesis in the rough endoplasmic reticulum through to newly formed granules [9, 51]. A potential role for PAM as a cargo receptor in pituitary cells remains to be explored. Taken together, these data underline the multiple roles that PAM can play in secretion of hormones. Studies of emerging PAM actions will be needed to address the influence of genetic variants in this study on PAM-mediated pituitary hormone release.

In conclusion, *PAM* was identified as a candidate gene associated with pituitary hypersecretion. Our comprehensive analysis of the impact of multiple *PAM* variants on protein function can also improve our understanding of other endocrine disorders recently linked to PAM defects, such as type 2 diabetes. Together, these new results raise the prospect of therapeutically targeting PAM function to influence disease in the pituitary and elsewhere across the endocrine system.

## Materials and Methods

The study population consisted of index cases and a validation cohort.

The index cases were part of a three member FIPA acro-gigantism family referred to the National Institutes of Health Clinical Center (NIHCC) as part of the Undiagnosed Diseases Program (UDP) [53–55]. The presence of germline SNVs/CNVs in genes known to predispose to pituitary tumors (*AIP*, *GPR101*, *CDKN1B*, *MEN1*, *PRKAR1A*, *GNAS*, and *DICER1*) had been previously ruled out in all affected individuals.

The validation cohort consisted of 359 individuals with PAs (Table S1). Germline DNA was studied in 299 unrelated individuals with sporadic PAs, in 17 FIPA families (including 19 individuals), and in three families with syndromic presentations of PAs and other endocrine tumors (MEN-like syndromes) [7, 56]. Among the sporadic cases, 173 had somatotroph adenomas (40 had pituitary gigantism), 84 had Cushing disease associated with corticotroph adenomas (CD, 81 were pediatric - age < 18 years at diagnosis - and one had Nelson syndrome), four had silent corticotroph adenomas, 25 had lactotroph adenomas (six were pediatric, one was a carcinoma), and 13 had clinically non-functioning or silent adenomas (one was pediatric). Somatic PA DNA was available from additional 38 subjects with sporadic acromegaly; five of them had matched germline DNA [7, 57]. Twenty-two of the pediatric CD cases (one familial and 21 sporadic) had matched tumor DNA available [56]. Subjects for the study were recruited from the Centre Hospitalier Universitaire (CHU), University of Liège (231), the NIH (90), and the Bicêtre Hospital and Foch Hospital Paris (38).

UDP patients provided written informed consent under clinical protocol 76-HG-0238, “Diagnosis and Treatment of Patients with Inborn Errors of Metabolism and Other Genetic Disorders”, approved by the NHGRI Institutional Review Board. Other individuals from the NIH were recruited between 1997-2017 under research protocol 97-CH-0076 (ClinicalTrials.gov: NCT00001595). The study was approved by the *Eunice Kennedy Shriver* National Institute of Child Health and Human Development (NICHD) Institutional Review Board and by the Ethics Committee of the University of Liège, Belgium. Affected subjects or their relatives signed informed consent or assent forms approved by the local ethics committee. Parents and siblings of the affected individuals were also recruited, when appropriate and available.

### DNA sequencing

#### Index cases

DNA was extracted from peripheral whole blood samples of the twin probands affected with gigantism from generation II, their father, two unaffected siblings, and the three individuals from generation III (sons of II-2). Whole exome sequencing (WES) was performed by a commercial vendor (Axeq Technologies, Rockville, Maryland) using the TruSeq Human Exome 62 Mb capture kit for library preparation for individuals from generation I and II. Exome sequencing was completed using the Illumina HiSeq2000 platform resulting in raw sequence FASTQ files. WES in individuals from generation III (and in II-2 as control for kit-specific artifacts) was performed using the NimbleGen SeqCap EZ Exome Library v3.0 kit (Roche) for library preparation. Exome sequencing was completed using the Illumina NovaSeq 6000 platform. Alignments were made to human reference assembly GRCh38 using BWA-MEM [58]. Variant calling and joint genotyping were performed using GATK Best Practices (GATK ver. 4.2.3.0, HTSJDK ver. 2.24.1, Picard ver. 2.25.4) [59]. The VCF files were annotated with population data from the gnomAD database (ver. 3.1.2). Functional and predictive annotations were attached using Ensembl Variant Effect Predictor (ver. 102) [60] and the dbNSFP (ver. 4.3a) [61] database. Pathogenicity predictions used in filtering came from SIFT, ClinPred, PolyPhen-2, FATHMM, and Combined Annotation Dependent Depletion (CADD) score. No known disease-causing or damaging biallelic/hemizygous variants segregating to the affected status were identified. Slivar [62] was used to filter the variants and apply pedigree analysis. We prioritized rare heterozygous damaging variants with nonzero pituitary median expression from GTEx that appear in affected individuals for downstream validation and analysis, excluding variants with a population minor allele frequency (MAF) > 1%. Variants reported in the ClinVar and/or the UniProt databases as benign or likely benign were also excluded. PCR (GoTaq Green Master Mix, M7123, Promega) and Sanger sequencing (BigDye Terminator 3.1 Cycle Sequencing Kit, 4337456, ThermoFisher Scientific) were used to confirm the *PAM* variant identified by WES. Sequences were aligned and analyzed using the SeqMan Pro 11.1.0 (DNASTAR Lasergene) software. Sequence chromatograms were visualized using SnapGene Viewer ver. 5.0.4.

#### Validation cohort

WES data on germline and tumor DNA samples from subjects with CD were generated as described previously [56]. These WES datasets were manually assessed using Integrative Genomics Viewer ver. 2.12.3 platform (Broad Institute) [63]. For all other subjects in the validation cohort, genomic DNA was extracted from peripheral blood and screened for *PAM* variants by Sanger sequencing. Whenever a *PAM* pathogenic/likely pathogenic variant was identified and family members were available for analysis, cascade screening was initiated. This led to two instances of genotype-positive family members. In 38 individuals with sporadic acromegaly, tumor DNA was available for analysis. However, due to the limited amount of DNA available, Sanger sequencing was targeted to specific regions of *PAM* where the majority of likely pathogenic variants were previously observed (Figure 2), i.e., the proximal promoter and exons 9, 16, 18, 19, 20, and 21. The primers included in Table S2 were used to amplify the promoter and 5’UTR regions, the coding regions and exon–intron junctions by PCR, and for Sanger sequencing. In the available adenomas, DNA was extracted from unstained sections using the Pinpoint Slide DNA Isolation System (D3001, Zymo Research) and loss of heterozygosity (LOH) was investigated by Sanger sequencing. The NC_000005.9(NM_000919.3) reference sequence was used to annotate *PAM* variants.

#### Single cell RNA sequencing (scRNA-seq) analysis

Figure S2A was derived from data published in the NCBI Gene Expression Omnibus (GEO) Series (accession number GSE132224) using methods described in detail [64]. Briefly, scRNA-seq data were derived from freshly dispersed rat anterior pituitary cells from postpubertal males (3,562 cells) and diestrus females (3,334 cells). Cell type clusters were identified using known genetic markers for secretory cell types, folliculostellate cells, erythrocytes, leukocytes, and endothelial cells. Here, we examined the expression levels of the *Pam* gene and the percentage of cells of each type expressing this gene, plotted as tSNE map [65] and percent-expressing heatmaps using Matlab (R2018b).

### CNV analysis

Sixteen pituitary gigantism patients and 121 other individuals with different subtypes of PAs were tested for germline *PAM* deletions/duplications by droplet digital PCR (ddPCR) using five FAM-labeled TaqMan CNV assays spanning the entire *PAM* gene (Hs03560663_cn in intron 2, Hs03041534_cn in intron 5, Hs06720527_cn in intron 13, Hs06071620_cn in intron 16, Hs06028580_cn in exon 25, all ThermoFisher Scientific). The VIC-labeled *RPP30* (*Rnase P*) assay (4403326, ThermoFisher Scientific) was used as an internal control. All PCR reactions were prepared using the ddPCR SuperMix for Probes (no dUTP, 1863024, Bio-Rad) and the HindIII restriction enzyme in a QX200 Droplet Digital PCR System (Bio-Rad). Results were analyzed with Quanta Soft software ver. 1.7.4.0917 (Bio-Rad).

### Allele phasing

The PacBio long seq run was employed to determine the phasing of *PAM* variants c.718C>T (p.His240Tyr) and c.2276T>C (p.Phe759Ser), located 60 kb apart (exons 9 and 20, respectively), in a person with sporadic gigantism. His parents were not available for genetic studies. Briefly, we employed Single Molecule, Real Time (SMRT) Sequencing technology and a Sequel sequencer (Pacific Biosciences, Menlo Park, CA). The general strategy was to generate a whole-genome library, then perform a hybridization-based pulldown to enrich for the genomic region including the two variants. The probes were targeted against chr5:102,939,611-103,019,811 (hg38 coordinates). This encompasses the positions of the two variants plus an extra 10 kb upstream and downstream for a total of about 83 kb. The biotinylated xGen Lockdown Probes were designed and synthesized by IDT (Coralville, IA) with one probe approximately every 1 kb, for a total of 86 probes. The long reads enable heterozygous variant walking to determine phasing of the variants. The library was constructed according to the Pacific Biosciences protocol given at https://www.pacb.com/wp-content/uploads/Procedure-Checklist-%E2%80%93-Multiplex-Genomic-DNA-Target-Capture-Using-IDT-xGen-Lockdown-Probes.pdf, except that SeqCap EZ reagents and the SeqCap Library SR User’s Guide (Roche, Indianapolis, IN) were used for the hybridization pulldown. Briefly, the genomic DNA was fragmented using a g-TUBE (Covaris, Woburn, MA) and Pacific Biosciences linear adapters were ligated to the ends to generate a library with an average insert length of about 10 kb. The region of interest was pulled down using a pool of 400 attomoles of each of the 86 probes described above. The resulting material was amplified and SMRTbell adapters were ligated to the ends. This library was sequenced on a Sequel (Pacific Biosciences) on a SMRT Cell 1M v2 cell with Sequencing kit 2.1 and a 10 hr acquisition time. The resulting reads were analyzed using the program “Targeted Phasing Consensus” from Pacific Biosciences (https://github.com/PacificBiosciences/targeted-phasing-consensus) to perform the read phasing and alignment using CCS reads (circular consensus sequences from 3+ passes), basic subreads, and the hg38 genome for the target region.

A male with sporadic acromegaly harbored *PAM* variants c.1654G>A (p.Gly552Arg) and c.1688A>G (p.Asp563Gly), both located in exon 16. To determine the phasing of these closely located SNVs, exon 16 was PCR-amplified from his germline DNA and the PCR product cloned into the TOPO-TA vector. Transformed bacterial colonies were screened for the presence of the variants by PCR and Sanger sequencing.

### Bioinformatic analyses

#### Variant pathogenicity and gene damage predictions

Besides the bioinformatic tools used in the variant prioritization process, Alamut Visual ver. 2.9 software (Interactive Biosoftware) was used for annotation, *in silico* prediction, and for determining the frequency in public databases of all the *PAM* variants identified. For nonsynonymous variants, unless specified, the computational verdict was based on the combined outputs of 19 software packages available in Varsome (accessed on 11/24/2022. Five algorithms (Splice Site Finder, MaxEnt, NNSplice, GeneSplicer and Human Site Finder) integrated in Alamut were used for splicing variants. Variants were considered probably damaging or affecting splicing when most of the algorithms agreed; otherwise, they were considered variants of uncertain significance (VUS). The gene damage index (GDI) [26] score for *PAM* and other genes known to predispose to pituitary tumors was retrieved from The Gene Damage Index Server.

#### Spatial clustering analysis

The spatial clustering of a set of variants is obtained by calculating the geometric mean distance between all pairs of variants and normalizing them to the gene’s cDNA length (2,922 bp, isoform P19021-5). An empirical p-value is calculated by randomly-generating 100,000 permutations of the variants and comparing their clustering distance against the clustering of the actual variants. As input, we used the 16 prioritized missense and truncating variants found in the index cases and validation cohort and reported in Table 2. We then classified the number of variants located in particular domains of the protein (converted to cDNA coordinates) and counted the number of variants appearing in each domain across all the permutations to calculate an empirical p-value comparing the actual number of variants found in each domain to the number of variants in each domain of the 100,000 permutation analysis.

#### Promoter analysis

The computational search for transcription factor binding sites within the promoter region of *PAM* was executed using Genomatix. The MatInspector ver. 3.1 tool of Genomatix (Matrix Library 11.0) was used with a core similarity threshold of 0.75 and an optimized matrix similarity threshold to search for the presence of transcription factor binding sites. We limited our search to vertebrate general core promoter elements and the + strand.

#### Protein sequence alignment and structural analysis

Protein sequence alignments were performed using the following sequences from UniProt and Clustal Omega: human PAM-1 (P19021-5), chimpanzee PAM-1 (A0A2I3SM67-1), rat PAM-1 (P14925-1), Aplysia PAM-1 (Q9NJI4-1), *Drosophila* PHM (O01404-1) and PAL2 (Q9W1L5-1), and *Chlamydomonas* PAM (A0A0S2C767-1). The enzymatically active cores for PHM and PAL were aligned. Enzyme activity has been demonstrated for each of these proteins except *Aplysia* and chimpanzee [39, 66–71]. We included chimpanzee as this species is evolutionarily close to human, rat because both its PHM and PAL domains have been crystalized and extensively analyzed [13, 72], *Aplysia* and *Drosophila* to cover other major families, and *Chlamydomonas* as the most evolutionarily distant species at the boundary of the animal/plant kingdoms. Full-length alignments are available in the Supplemental Methods. Ribbon diagrams of the crystal structures of the catalytic cores of rat PHM (PHMcc) (PDB: 1OPM) and PAL (PALcc) (PDB: 3FW0) were used to contextualize the identified variants. 3D models were generated using the PyMOL molecular graphics system (Schrödinger, LLC). Rat PHMcc and PALcc are, respectively, 93% and 92% identical to human PHMcc and PALcc.

#### Analysis of *PAM* SNV enrichment in pituitary diagnoses from the UK Biobank (UKBB) cohort UKBB dataset and disease annotation

The UKBB is a prospective study that recruited 502,611 participants aged 38 to 73 years from 22 sites across the UK with baseline measures collected between 2006 and 2010 [73]. This research was conducted using the UKBB Resource (Application Number 48008). The UKBB obtained ethics approval from the North West Multi-centre Research Ethics Committee (approval number: 11/NW/0382) and obtained informed consent from all participants. Diagnoses of hyperfunctioning pituitary diseases were retrieved from hospital inpatient diagnosis fields 41270, 41202, 41204, with International classification of diseases (ICD-10) codes E220 (acromegaly and pituitary gigantism), E221 (hyperprolactinemia), E222 (syndrome of inappropriate secretion of antidiuretic hormone), E229 (unspecified hyperfunction of the pituitary gland), E240-E248-E249 combined as E240 (pituitary-dependent Cushing disease), D352-D359 combined as D352 (benign neoplasm of pituitary gland), E237 (disorder of pituitary gland), or as self-reported medical condition with codes 1237 (disorder of pituitary gland combined with ICD-10 code E237), 1238 (pituitary adenoma/tumor combined with ICD-10 code D352), 1239 (Cushing’s syndrome combined with ICD-10 code E240), 1431 (hyperprolactinemia combined with ICD-10 code E221), 1429 (acromegaly combined with ICD-10 code E220).

#### SKAT analysis

Gene-based diagnosis-association analysis was performed on 200,643 individuals for whom exome sequencing data were available using the Sequence Kernel Association Test (SKAT). The R package SKAT was employed [24]. We used the CommonRare algorithm for binary traits with the adaptive sum method to account for common and rare variants within the *PAM* gene, using a MAF cutoff of 0.1% and considering the weight of the SNVs based on their detrimental effect on protein. SNV weights were assigned as previously described [74] and adapted as follows:

- weight of 5 for UTR variants, synonymous variants, splice region variants;
- weight of 10 for protein altering variants;
- weight of 20 for start lost, stop lost, in-frame deletions, in-frame insertions;
- for missense variants, weight of 20 + score from combined PolyPhen-2 and SIFT predictions: PolyPhen-2 +10 if possibly damaging or +20 if probably damaging or +5 if unknown; SIFT +20 if deleterious, +0 if tolerated;
- weight of 75 for frameshift variants, nonsense, splice acceptor variants, splice donor variants;
- weight of 100 for transcript ablation.

Statistical significance for gene-based tests was set at a Bonferroni-corrected threshold of p < 2×10^−6^ (threshold for 25,000 genes).

#### Analysis of SNVs in UKBB

Analysis of SNVs in UKBB individuals with pituitary diagnoses was performed with PLINK and a custom code in Python ver. 3.7. The CADD v1.660 VEP plugin was used to provide prediction scores for their detrimental effect. A CADD_PHRED score of ≥15 and a MAF < 1% were considered as pathogenicity criteria.

### Functional characterization of SNVs

#### Cell lines

The PEAKrapid (ATCC CRL-2828) and HEK-293 AD (ATCC CRL-1573) cells are lines derived from the Human Embryonic Kidney (HEK)-293 cell line. The PEAKrapid cells were maintained in Dulbecco’s modified Eagle’s medium (DMEM)-F12, with 10% fetal bovine serum (FBS, Hyclone), pen-strep, and HEPES to net 25 mM, while the HEK-293 AD cells were maintained in DMEM containing low glucose, pyruvate, 2 mM glutamine (Gibco), with 10% FBS (Gemini Bio Products), and 1% antibiotic–antimycotic (Gibco). Both cell lines were kept in a humidified atmosphere at 37°C with 5% CO_2_.

#### Vectors and mutagenesis

All cloning reactions were performed using the In-Fusion cloning system (Clontech). Cloning primers are reported in Table S2. Tissue-specific alternative splicing generates multiple PAM isoforms [75]. The longest human isoform is a 974 amino acid integral membrane protein identified as P19021-5 in UniProt. Since a GenBank search revealed that this isoform is more prevalent than the one chosen as the canonical sequence (UniProt P19021-1/NM_001177306.2, 973 amino acid-long), we regarded it as human PAM-1, corresponding to what was previously extensively characterized as rat PAM-1, a 976 amino acid integral membrane protein. The ORF expression clone for wild-type (WT) human PAM-1 (NM_000919.3) cloned into pReceiver-M02 (EX-A3104-M02-GS, Genecopoeia) was subcloned into the pCMV-FLAG-C vector (635688, Clontech) using the EcoRI and XhoI restriction sites. The resulting PAM-1 protein has an in-frame C-terminal FLAG tag (DYKDDDDK) separated from its normal C-terminus by a five amino acid spacer (LEVPA). A 5,373 bp sequence including the human WT *PAM* promoter (5,000 bp) and 5’UTR (373 bp) was generated by *de novo* gene synthesis (Blue Heron Biotech) and cloned into the pRMT-Luc vector (PR100001, Origene) using the EcoRI and MluI restriction sites located upstream of *Firefly* luciferase. The WT and mutant (harboring the c.*1455C>T variant) 3’UTRs of human *PAM* (2,040 bp) were PCR-amplified from human genomic DNA and cloned into psiCHECK-2 (Promega) using the XhoI and NotI restriction sites located downstream of *Renilla* luciferase. WT and mutant *PAM* sequences consisting of seven different human *PAM* exons (exon 2, 6, 16, 18, 19, 20, 21) together with their flanking intronic sequences (100/300-bp-long) were PCR-amplified from human genomic DNA and cloned into the EcoRI-digested pSPL3 vector (Invitrogen). See Supplemental Methods for more details.

Mutagenic primers (reported in Table S2) were used to introduce *PAM* SNVs using the QuikChange II XL site-directed mutagenesis kit (Agilent Technologies). Mutagenesis products were verified by Sanger sequencing. Since our findings (see the Results section) showed a discrepancy with what was previously reported by *Thomsen et al*. [15] for the p.Ser539Trp variant, we prepped the construct harboring this variant twice and sequence-verified it again. To introduce the deletion caused by the splice-altering c.2332-2A>T variant into the PAM-1_pCMV-FLAG-C vector, the PCR splicing technique was used. A pair of primers flanking the region where the deletion occurs (entire exon 21) and a pair of complementary primers comprising a region of -15 bp to +15 bp related to the junction point are reported in Table S2.

#### Antibodies

Antibodies used for the various experiments included the following: an affinity-purified rabbit polyclonal antibody (Ab JH629) raised to purified bacterially expressed Exon A (exon 16 in rat *Pam*) [76]; a rabbit polyclonal antibody (Ab JH246) raised to a synthetic peptide corresponding to human PAM(111-126), which is identical to rat PAM(116-131) [77]; a mouse anti-FLAG monoclonal antibody (F1804, Sigma-Aldrich); and an anti-GM130 antibody (#610822, BD Biosciences).

#### Biochemical analysis of nonsynonymous and splice site-affecting SNVs

Transient transfection of PAM-1 WT and mutant vectors was performed 24 h after plating using PEAKrapid cells and TransIT-2020 (MIR 5400, Mirus Bio) [78]. Cells were harvested 24-36 h after transfection. Protein extraction and western blotting were performed as described previously [76]. After rinsing with serum-free medium, cells were scraped into serum-free medium and pelleted. Cell pellets were extracted into ice cold 20 mM Na TES, 10 mM mannitol, 1% TX-100, pH 7.4 (TMT) containing protease inhibitors and subjected to three freeze/thaw cycles. Particulate material was removed by centrifugation and protein concentrations were determined using the bicinchoninic acid assay with bovine serum albumin (BSA) as the standard. For enzyme assays, lysates were diluted using TMT containing 1 mg/ml BSA. For Western blot analysis, lysates were denatured using 2X or 4X Laemmli Sample Buffer (Bio-Rad).

PHM and PAL assays were performed using [^125^I]-Ac-Tyr-Val-Gly and [^125^I]-Ac-Tyr-Val-α-hydroxy-Gly with 0.5 μM unlabeled substrate, as described previously [79]. Samples were assayed in triplicate, within the linear range of the assay. Data for mutants were normalized to data for human PAM-1 analyzed in parallel. Normalized data from multiple different transfections were averaged to ensure that each mutant was assessed in at least two independent transfections. Human serum samples were diluted 10-fold into 20 mM Na TES, 10 mM mannitol, 1 mg/ml bovine serum albumin (PHM diluent). Assays for PHM and PAL activity were carried out in duplicate or triplicate at pH 5.5; PHM assays included 4 μM CuSO_4_ and PAL assays contained 1 mM CoCl_2_.

Analysis of the glycosylation patterns for PAM WT and mutants was performed as described previously [39]. Cells were harvested 24 h after transfection. Cell pellets were frozen and thawed in 20 mM Na TES, 10 mM mannitol, pH7.4 containing a protease inhibitor mix. Following centrifugation at 17,000 g for 20 min at 4°C, the pellets were extracted in TMT with protease inhibitors and the supernatants were denatured using SDS and digested with PNGase F or neuraminidase as described by the manufacturer (NEB). For PAM and for each PAM variant, one aliquot remained on ice (In, Input) and two aliquots were prepared for enzymatic digestion; both were incubated at 37°C for 60 min, but one received enzyme (N-Gly or Neur) and one did not (Con). After SDS-PAGE and transfer to PVDF membranes, epitope-tagged PAM and its variants were visualized using a FLAG antibody. Neuraminidase removes sialic acid from both N-linked and O-linked oligosaccharides. Samples treated with neuraminidase were analyzed on two separate gels, with the p.Arg703Gln samples appearing in part on both gels.

#### Analysis of splicing by minigene assay

Transient transfection of WT and mutant pSPL3 vectors was performed using HEK-293 AD cells and TurboFect (ThermoFisher Scientific). Cells were transfected 24 h after plating; 24 h after transfection, total RNA was extracted using the RNeasy Plus Mini Kit (Qiagen). RNA (500 ng) was reverse transcribed to cDNA using the Superscript III Kit (ThermoFisher Scientific). mRNA synthesis from the plasmids using the cells’ own transcription and splicing machinery led to mRNA products containing the tested *PAM* exon flanked by two exons from the pSPL3 vector. Splicing products were analyzed by RT-PCR using the vector-specific SD6 and SA2 primers (Table S2). The empty pSPL3 vector was used as negative control.

#### Analysis of SNVs in regulatory regions by luciferase-based reporters

SNVs located upstream (promoter and 5’UTR) and downstream (3’UTR) of *PAM* CDS, were investigated using luciferase-based reporter assays transfected into HEK-293 AD cells. The empty pRMT-Luc and psiCHECK-2 vectors were used as negative controls. For the 3’UTR experiments, four microRNA (miRNA) mimics (hsa-miR-138-2-3p, MC12814; hsa-miR-192-3p, MC12893; hsa-miR-3143, MC17293; hsa-miR-556-3p, MC12806; all ThermoFisher Scientific) were transfected alongside the WT and mutant reporter vectors. Cells were lysed 24 h after transfection and *Firefly* and/or *Renilla* luciferase activities were measured using the Dual-Luciferase Reporter Assay System (Promega) following the manufacturer’s protocol. When appropriate, ratios of *Firefly*/*Renilla* luminescence signals, serving as a measure for reporter activity normalized for transfection efficiency, were determined using a FLUOstar Omega microplate reader (BMG Labtech).

### Immunohistochemistry

Immunofluorescence analysis was performed on histological sections from non-pathological pituitary cells (control). The full protocol has been detailed elsewhere [80]. Briefly, all slides were blocked with 10% normal donkey serum in 1X PBS for 1 h at room temperature and then incubated overnight at 4°C with affinity-purified 1:500 anti-PAM JH629 and anti-GM130. All slides were incubated for 1 h with 1:500 donkey anti-rabbit 488 and donkey anti-mouse 594 (respectively, A-11055 and A-21207, ThermoFisher Scientific). Slides were imaged at 40× on a Keyence BZ-X710 microscope.

### Statistical analysis

All graphs were plotted as mean ± standard error of the mean (SEM). Data distributions were assessed for approximate normality. Differences between experimental groups were analyzed by two-tailed Student’s t-test or 1-way ANOVA with Dunnett’s *post hoc* test, or corresponding non-parametric tests, as appropriate. *PAM* SNV allele frequencies in the study population were compared with frequencies in the general population reported in the gnomAD public database using Fisher’s exact test or the chi-square test, as appropriate. Data were analyzed using GraphPad Prism (GraphPad, San Diego, CA, USA). p-values < 0.05 were considered statistically significant.

## Data Availability

All data produced in the present study are available upon reasonable request to the authors

https://github.com/NICHD-BSPC/spatial_clustering

## Acknowledgments

The authors would like to thank Steven Coon of the Molecular Genomics Core of NICHD/NIH for conducting the phasing experiment, the late Mario Amzel (Johns Hopkins University School of Medicine) for useful comments on the impact of *PAM* variants on its 3D structure, Lyssikatos Charalampos and María de la Luz Sierra (NICHD/NIH) for their help with the collection and preparation of DNA and histopathological samples used in this study; Emilie Castermans and Leonor Palmeira (CHU de Liège) for genetic and bioinformatic analyses of FIPA kindreds, and Jean-Francois Bonneville (CHU de Liège, Belgium) for discussions on the neuroradiological images.

## Data and Code Availability

The code used to conduct the spatial clustering analysis can be found at https://github.com/NICHD-BSPC/spatial_clustering.

## Web Resources

BWA, https://github.com/lh3/bwa

CADD, https://cadd.gs.washington.edu/

ClinVar, https://www.ncbi.nlm.nih.gov/clinvar/

Clustal Omega, https://www.ebi.ac.uk/Tools/msa/clustalo/

dbNSFP (ver. 4.3a), http://database.liulab.science/dbNSFP

dbSNP, https://www.ncbi.nlm.nih.gov/snp/

Ensembl VEP (ver. 102), https://grch37.ensembl.org/info/docs/tools/vep/index.html

GATK (ver. 4.2.3.0), https://gatk.broadinstitute.org/hc/en-us/articles/4409678362139-GATK-4-2-3-0-release

Genomatix, https://www.genomatix.de/

gnomAD (ver. 3.1.2), https://gnomad.broadinstitute.org/

Gene Structure Display Server (GSDS 2.0), http://gsds.cbi.pku.edu.cn/

GTEx Project, https://www.gtexportal.org/home/

HIPred score, https://github.com/HAShihab/HIPred

HTSJDK (ver. 2.24.1), https://samtools.github.io/htsjdk/

OMIM, https://www.ncbi.nlm.nih.gov/omim

PCR splicing technique, http://www.methods.info/Methods/Mutagenesis/PCR_splicing.html

Picard (ver. 2.25.4), http://broadinstitute.github.io/picard/

PolyPhen-2, http://genetics.bwh.harvard.edu/pph2/

Primer3, https://www.ncbi.nlm.nih.gov/omim

Protein Molecular Weight, https://www.bioinformatics.org/sms/prot_mw.html

Slivar, https://github.com/brentp/slivar

The Gene Damage Index (GDI) Server, http://pec630.rockefeller.edu:8080/GDI/

The Human Protein Atlas, https://www.proteinatlas.org/

UniProt, https://www.uniprot.org/

Varsome, https://varsome.com/

