## Supplemental Data for "Germline loss-of-function *PAM* variants are enriched in subjects with pituitary hypersecretion"

### **Overview of Supplemental Information**

### **Supplemental Figures**

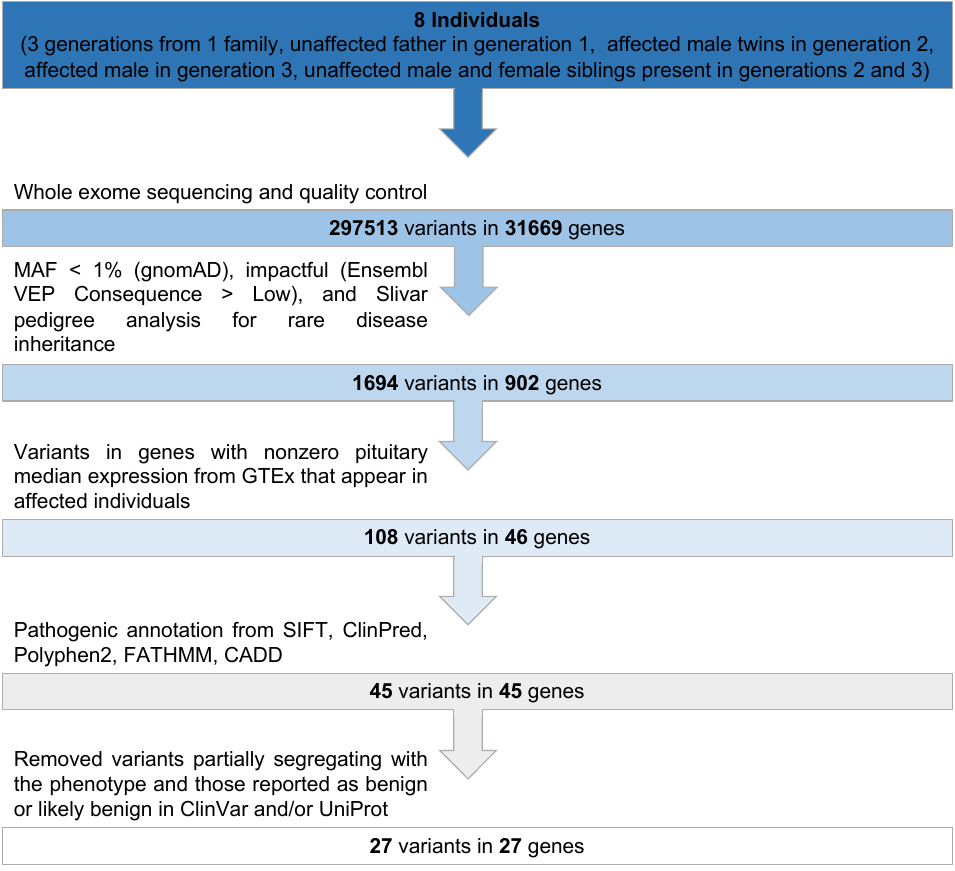

**Figure S1.** **WES analysis pipeline**

Analytic pipeline detailing the SNV filtering steps applied in the WES analysis of the index cases.

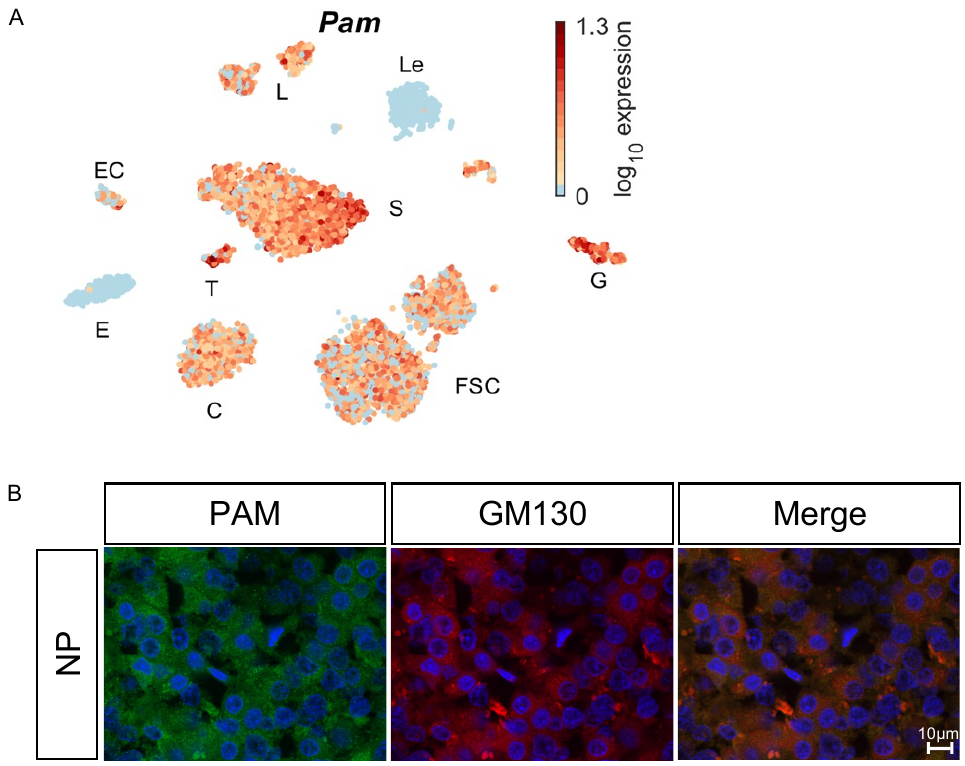

**Figure S2.** **PAM mRNA and protein expression studies in pituitary cells**

(A) tSNE map showing the expression of *Pam* gene in freshly dispersed adult rat anterior pituitary cells.  E, erythrocytes, Le, leukocytes (both negative), L, lactotropes; EC, endothelial cells; S, somatotropes; G, gonadotropes, T, thyrotropes; C, corticotropes; and FSC, folliculostellate cells (all expressing *Pam*). Data are shown on Log10 scale. (C) Immunofluorescence staining for PAM (green) and GM130 (red) in non-pathological pituitary cells (NP). PAM was visualized using an antibody (JH629) raised against the linker region of PAM-1 that recognizes intact PAM-1, soluble PHM, and membrane PAL. Magnification for all images: 40×.

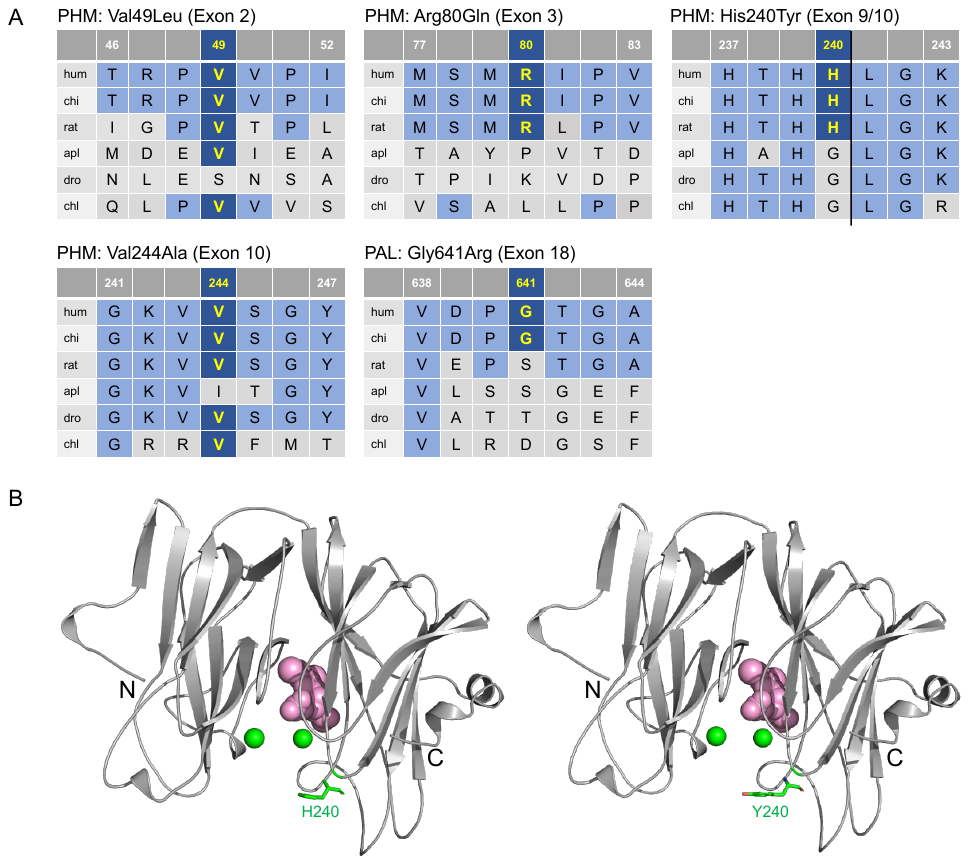

**Figure S3.** **Evolutionary conservation and structure-based analyses for six missense *PAM* variants**

(A) Protein sequence alignments for five *PAM* variants in which no significant change in PHM or PAL activity was observed. Based on *in silico* and *in vitro* data, four variants are considered likely non-pathogenic, while we classified p.His240Tyr as a variant of uncertain significance (VUS). The alignments were performed using Clustal Omega with default settings using the UniProt alignment tool. Conserved affected residues are given in yellow. (B) The crystal structure of rat PHMcc (PDB entry 1OPM) was used to contextualize the p.His240Tyr *PAM* variant with respect to the active site; the WT residue is shown on the left and the mutant residue on the right. The two copper ions are depicted as green spheres, while the substrate, N-acetyl-3,5-diiodotyrosylglycine, is shown in pink.

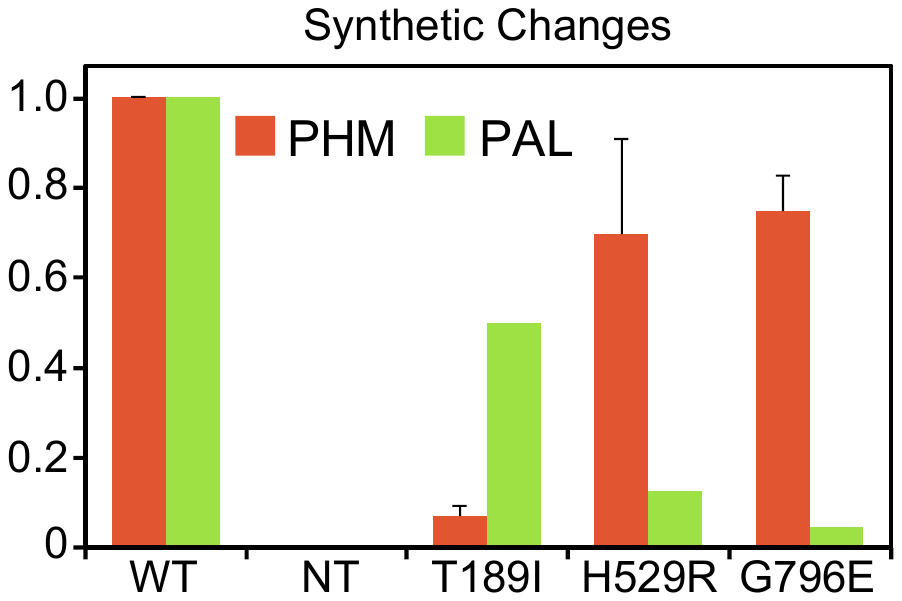

**Figure S4.** **PHM and PAL activity for three engineered PAM variants**

As described in Methods, TMT lysates prepared from transiently transfected PEAKrapid cells were assayed for PHM activity and for PAL activity. Data for the level of expression of WT PAM and each variant were determined by quantifying the FLAG-tag signal. Data for variants expressed in any single transfection were normalized to WT PAM expressed and processed at the same time. p.Thr189Ile is located in PHMcc and p.His529Arg and p.Gly796Glu in PALcc.

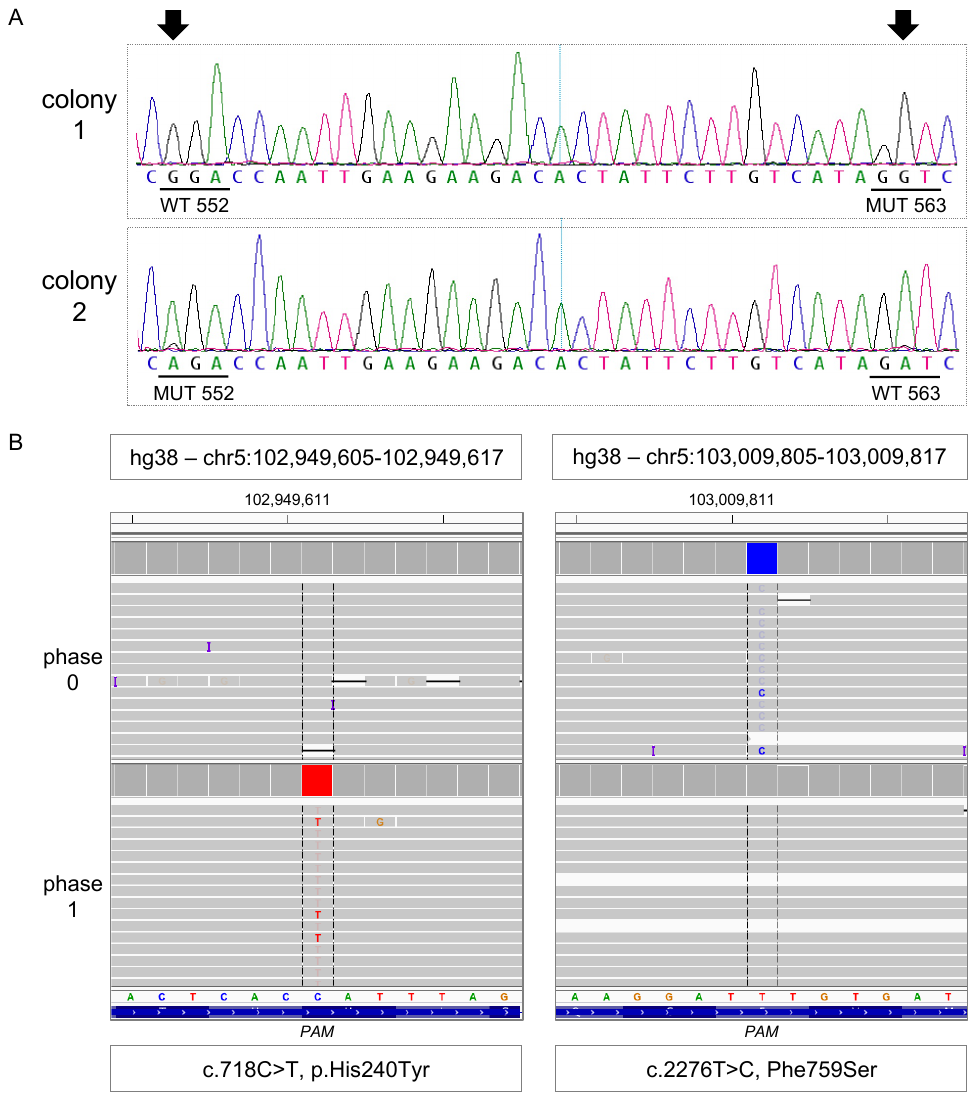

**Figure S5. Allele phasing analyses**

(A) A subject with sporadic acromegaly (Belgium128) harbored two missense variants in exon 16 of *PAM*: c.1654G>A (p.Gly552Arg) and c.1688A>G (p.Asp563Gly), indicated by black solid arrows. To determine the phasing of these closely located SNVs, exon 16 was PCR-amplified from his germline DNA and the PCR product cloned into the TOPO-TA vector. Transformed bacterial colonies were screened for the presence of the variants by PCR and Sanger sequencing. Two colonies showing a different variant each are shown, confirming the subject is a compound heterozygote. MUT, mutant; WT, wild-type. (B) A subject with sporadic gigantism harbored two missense variants – c.718C>T (p.His240Tyr) and c.2276T>C (p.Phe759Ser) – located 60 kb apart (exons 9 and 20, respectively). To determine the phasing of these distantly located SNVs, we employed Single Molecule, Real Time (SMRT) Sequencing technology and a Sequel sequencer (Pacific Biosciences, Menlo Park, CA). The reads were split into the two alleles based on the heterozygous mutation walking. The coverage was continuous across the capture region with low-points of about 15x fold in either allele. The top track is the “phase 0” allele and the bottom track the “phase 1” allele. The c.718C>T variant was assigned to the phase 1 allele, while the c.2276T>C variant was assigned to the phase 0 allele, confirming that the subject is a compound heterozygote.

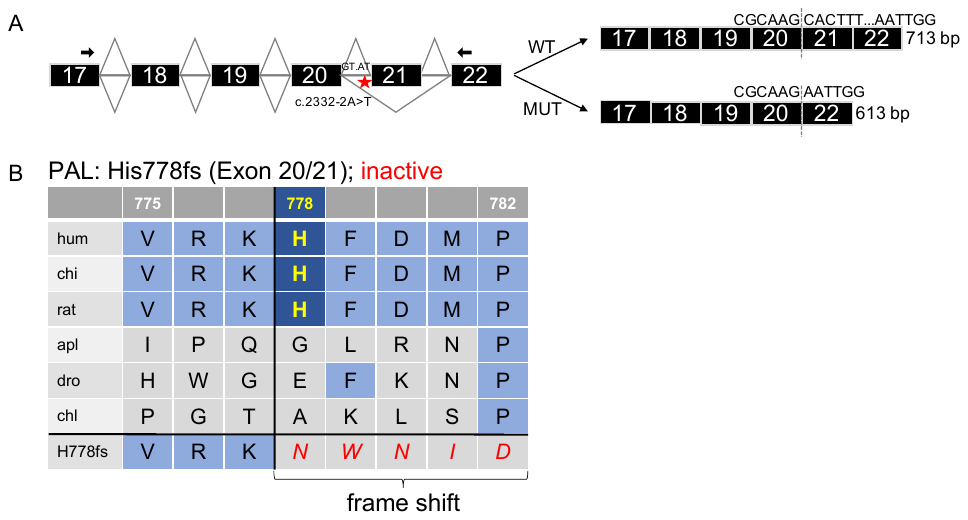

**Figure S6. Predicted splicing transcripts and introduced frame shift for the c.2332-2A>T variant**

(A) Diagram depicting the location of the c.2332-2A>T variant in intron 20 of *PAM* (red star) and the expected splicing events generating a normal transcript (WT) and an abnormal one lacking exon 21 (MUT). The location of the primers used for the RT-PCR analysis from blood-extracted RNA is given by the black solid arrows. MUT, mutant; WT, wild-type. (B) Protein sequence alignment for the encoded p.His778fs variant that abolishes PAL activity and reduces PHM activity (Table 3). The alignment was performed using Clustal Omega with default settings using the UniProt alignment tool. The affected residue is given in yellow when conserved. The beginning of the predicted frame shift is visible in the last row and the newly introduced amino acids are given in red.

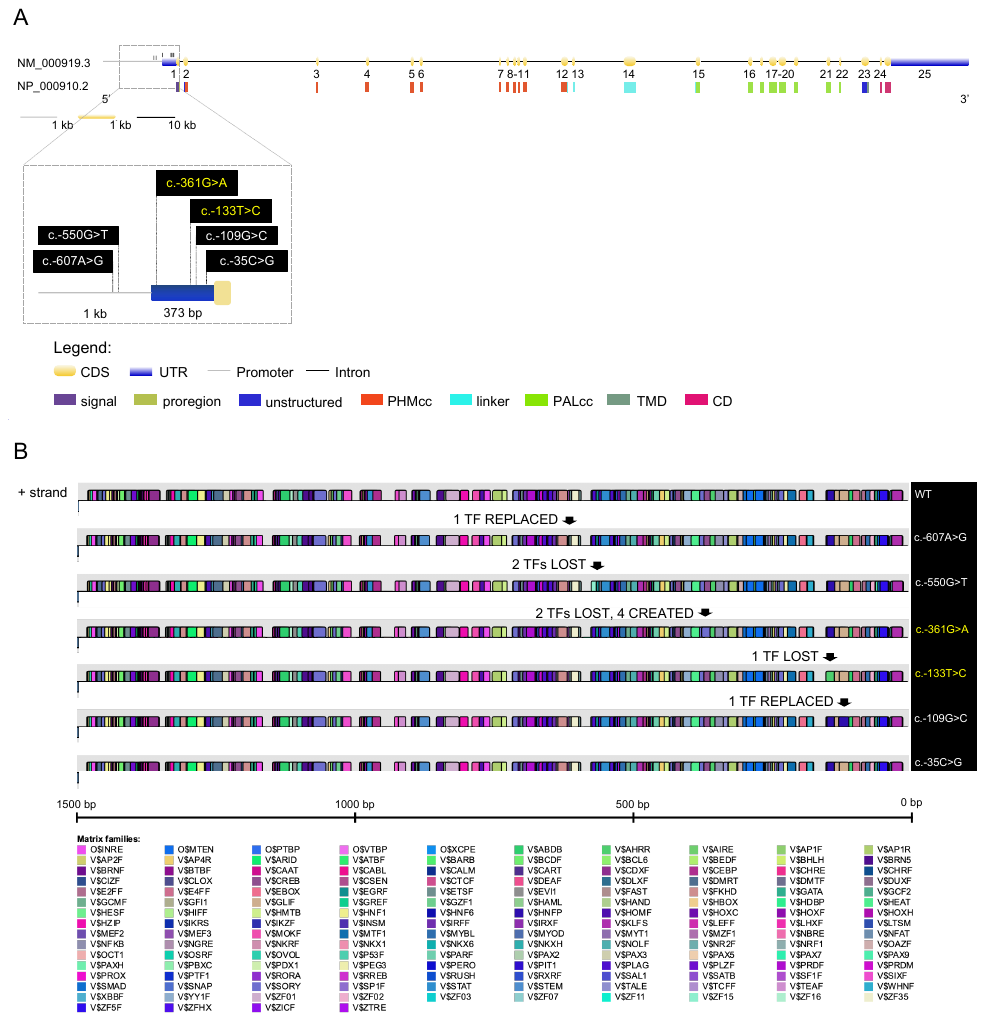

**Figure S7. Location and *in silico* predictions for six *PAM* variants located in regulatory regions**

(A) Schematic representation of the *PAM* gene (GenBank: NM_000919.3, PAM-1, 25 exons) and encoded protein (NP_000910.2, 974 amino acids), including functional domains. Six functionally tested SNVs located in the promoter region and 5’UTR are shown in the zoom in panel. Variants shown to reduce PAM transcription are given in yellow, while those without an identified effect are given in white. CD, cytosolic domain; CDS, coding sequence; PAL, peptidyl-α-hydroxyglycine α-amidating lyase; PHM, peptidylglycine α-hydroxylating monooxygenase; TMD, transmembrane domain; UTR, untranslated region. (B) Genomatix prediction depicting the transcription factor (TF) binding sites perturbed by the six variants. The MatInspector v3.1 tool of Genomatix (Matrix Library 11.0) was used with a core similarity threshold of 0.75 and an optimized matrix similarity threshold to search for the presence of TF binding sites. We limited our search to vertebrate general core promoter elements and the + strand. WT, wild-type.

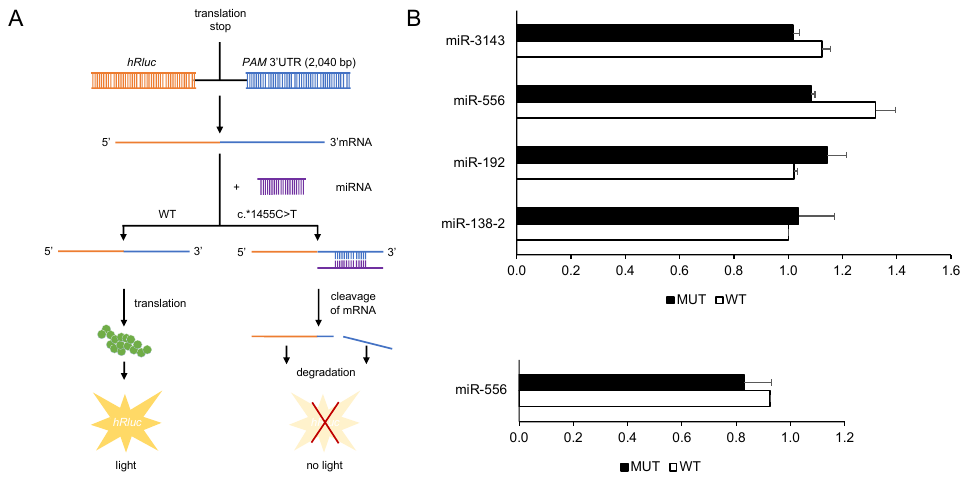

**Figure S8. Functional study for the c.*1455C>T 3’UTR variant**

(A) Cartoon depicting the experimental setup and the expected outcome based on the *in silico* predictions. The image was adapted from the siCHECK^TM^ Vectors Technical Bulletin (TB329, Promega). The WT and mutant 3’UTRs of *PAM* were PCR-amplified from human genomic DNA and cloned into the psiCHECK-2 vector, downstream of the *Renilla* Luciferase CDS. The constructs were transiently transfected into HEK-293 AD cells together with a *Firefly* reporter for normalization and each of the four miRNAs mimics predicted to bind the mutated 3’UTR. *Firefly* and *Renilla* activities were then measured. MUT: mutant; UTR, untranslated region; WT, wild-type. (B) miRNAs were initially transfected at 30 nM (n = 2 biological experiments each with three technical replicates, upper panel); the concentration of miR-556 was subsequently increased to 50 nM (n = 2 biological experiments each with three technical replicates, lower panel). This analysis failed to find a significant decrease in luciferase activity, suggesting that the mutant 3’UTR of *PAM* is not bound by the predicted miRNAs.

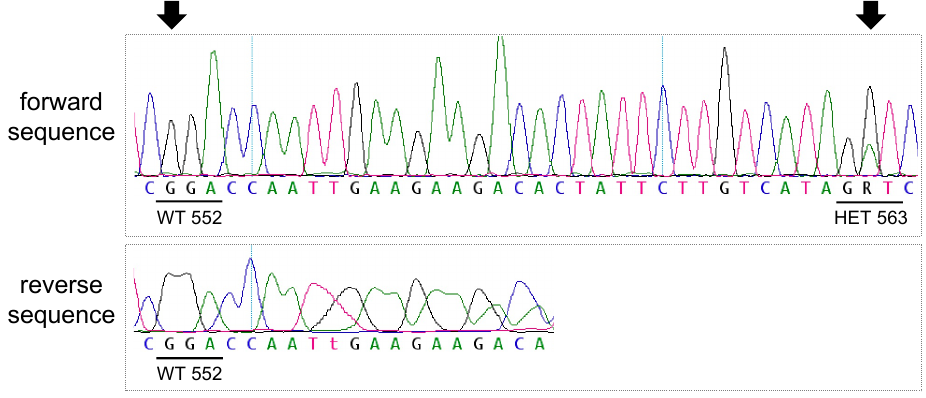

**Figure S9. Loss of heterozygosity (LOH) analysis for *PAM* in the pituitary adenoma of the individual Belgium197 compound heterozygote for the p.[Gly552Arg];p.[Asp563Gly] variants**

The DNA was extracted from unstained sections and LOH was investigated by Sanger sequencing. The primers used for the PCR amplification of exon 16, which harbors both variants observed in the germline DNA (Figure 2A), are reported in Table S1. The black solid arrows indicate the affected nucleotides. HET, heterozygous; WT, wild-type.

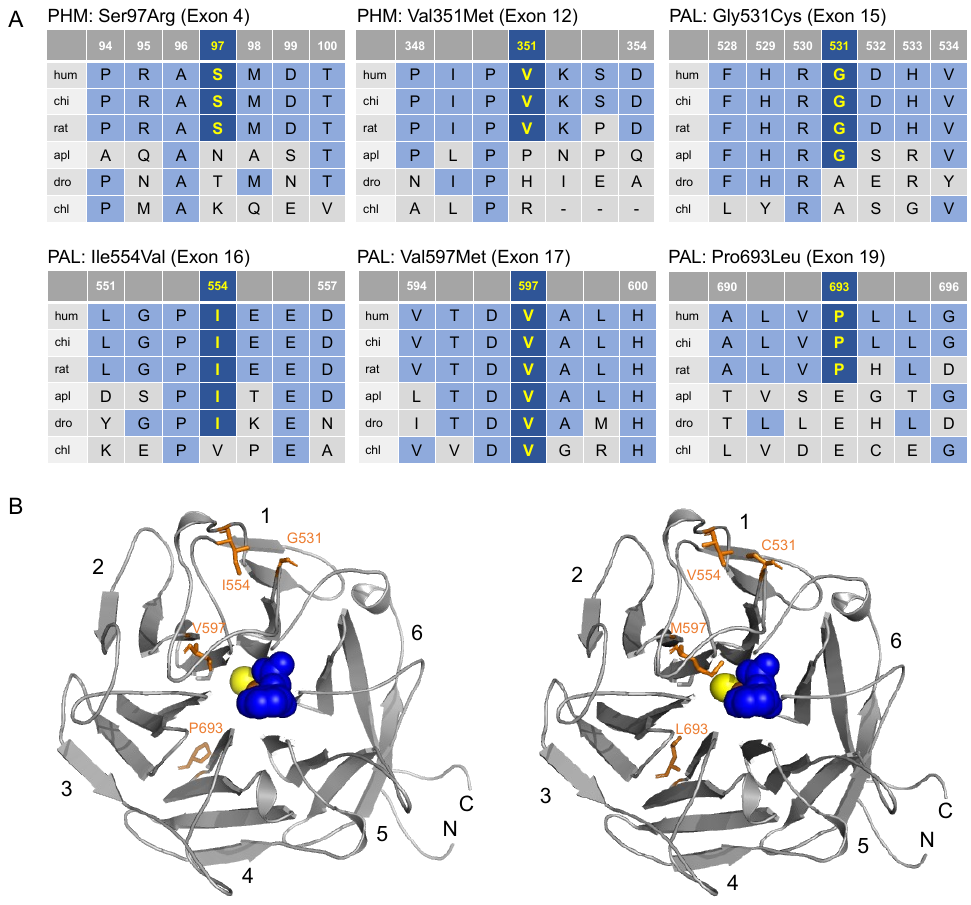

**Figure S10. Rare missense variants identified in the UKBB as significantly associated to diagnoses of tumoral/hyperfunctioning pituitary gland**

(A) Protein sequence alignments were performed using Clustal Omega with default settings using the UniProt alignment tool. Conserved affected residues are given in yellow. Only the variants located in PHMcc and PALcc are shown. (B) The crystal structure of rat PALcc (PDB entry 3FW0) was used to contextualize the missense variants; the WT residue is shown on the left and the mutant residue on the right. PAL folds as a β-propeller, with six blades (numbered 1 to 6) positioned around a central cavity. The affected residues are visualized as sticks and highlighted in dark orange, along the ribbon visualization of WT rat PALcc in grey. C, C-terminus; N, N-terminus.

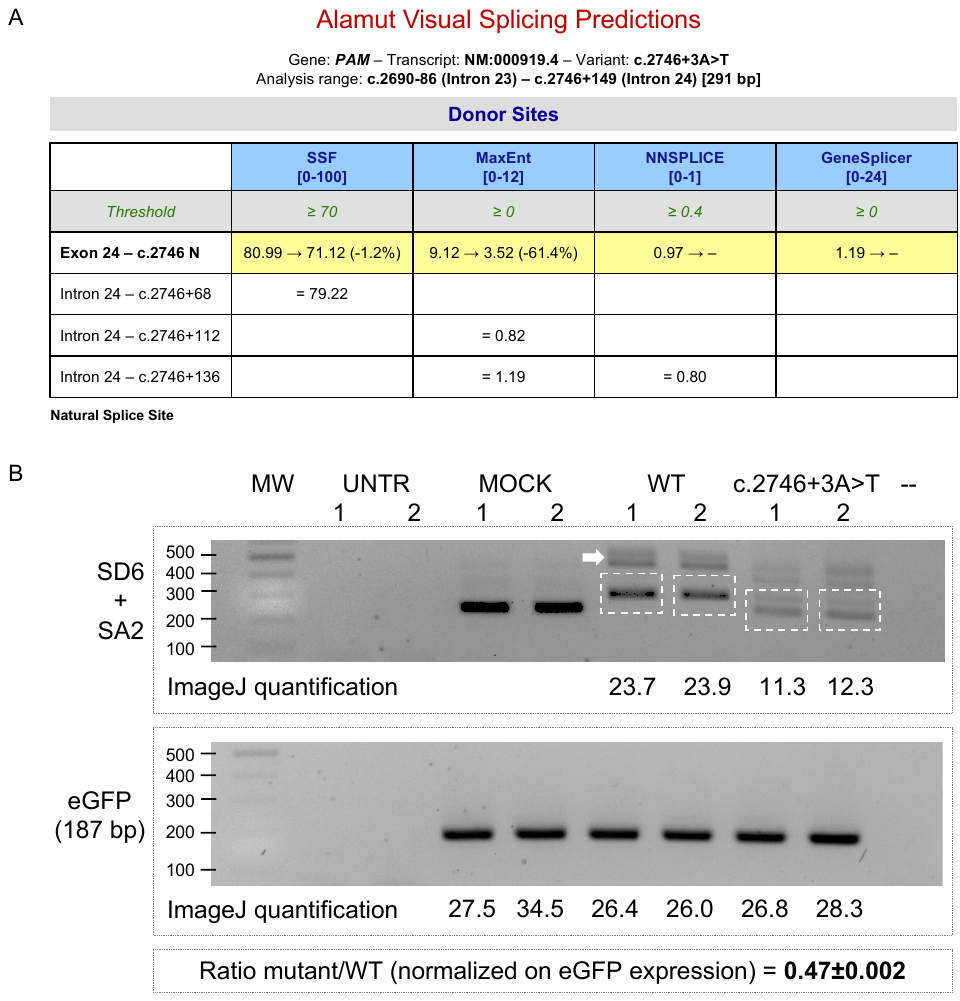

**Figure S11. Bioinformatic and *in vitro* analyses for the intronic c.2746+3A>T (p.(Ala897_Arg915del))**

***PAM* variant identified in the UKBB**

(A) The Alamut Visual 2.9 software was employed for splicing assessment. The variant was predicted to reduce the usage of the natural splice donor site in intron 24 by all four splicing tools. (B) The variant was functionally tested with a minigene splicing assay. *PAM* exon 24 and flanking intronic sequences were cloned into the pSPL3 vector. The full-length insert is reported in the Supplemental Methods. Then, the c.2746+3A>T variant was introduced by site-directed mutagenesis. The WT and mutant vectors were transfected into HEK-293 AD cells alongside an eGFP plasmid. 24h post-transfection, RNA was extracted and RT-PCR was performed using both pSPL3-specific (SD6 + SA2) and eGFP primers. Two replicates are shown for each condition. In both the WT and mutant constructs, the two upper bands represent non-specific splicing (white arrow), while the lower band in WT represents correct splicing. The variant causes a reduction in the amount of correctly spliced transcripts, favoring exon skipping (260 bp band, same as in MOCK). Densitometric quantification of both bands (enclosed by white dotted squares) was evaluated using the ImageJ software. The results were normalized on eGFP expression. All bands were confirmed by sequencing. MOCK, cDNA from empty vector-transfected cells consisting of a 260 bp band made up of fragments of pSPL3 exons; MW, molecular weight marker; UNTR, untransfected; WT, wild-type; --, negative control (RT-PCR without cDNA).

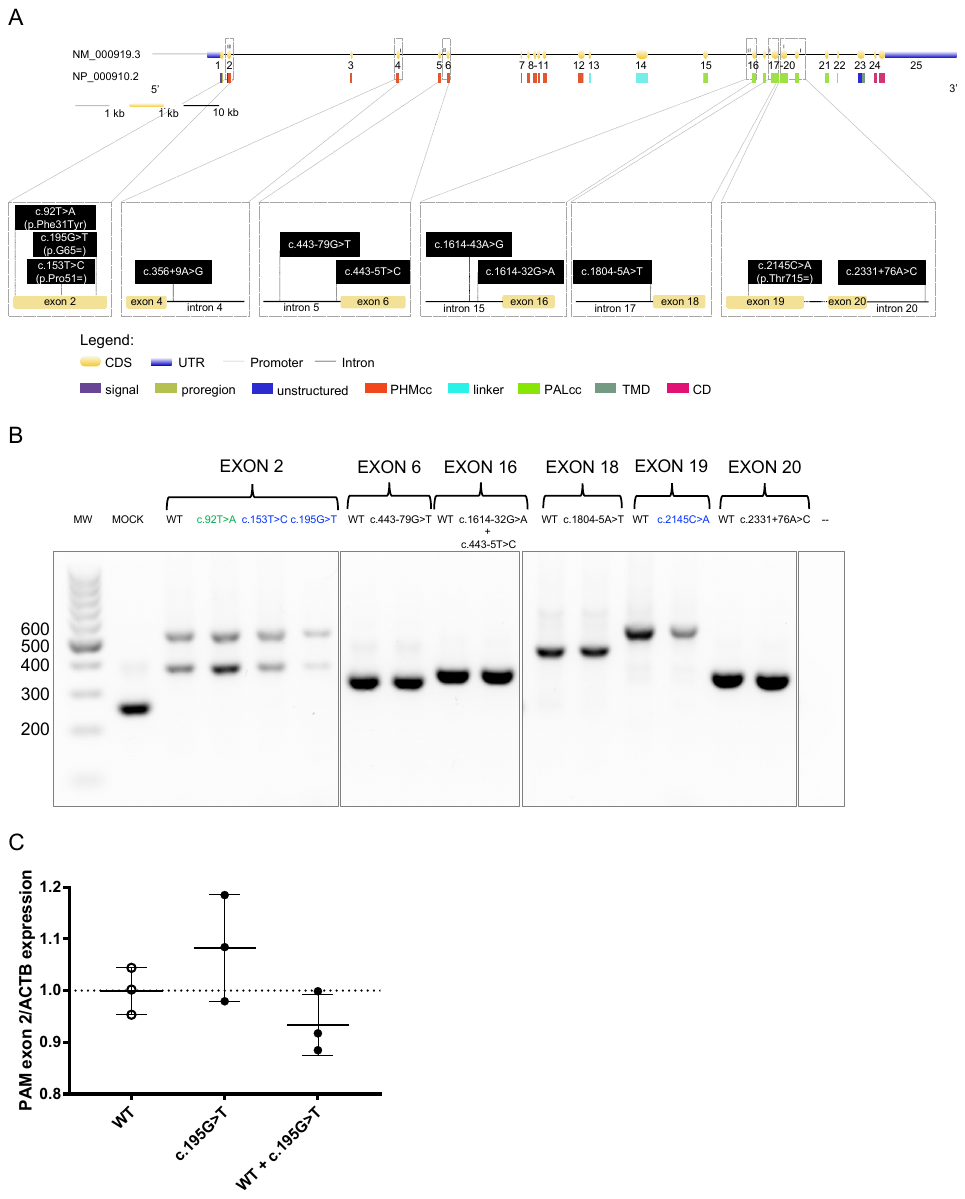

**Figure S12. Splicing and mRNA expression studies for selected *PAM* variants found in our cohort**

(A) Schematic representation of the *PAM* gene (GenBank: NM_000919.3, PAM-1, 25 exons) and encoded protein (NP_000910.2, 974 amino acids), including functional domains. Eleven functionally tested SNVs located along the gene are shown in the zoom in panels. CD, cytosolic domain; CDS, coding sequence; PAL, peptidyl-α-hydroxyglycine α-amidating lyase; PHM, peptidylglycine α-hydroxylating monooxygenase; TMD, transmembrane domain; UTR, untranslated region. (B) Nine *PAM* SNVs were evaluated for their effect on splicing using a minigene assay. The variants were cloned into the pSPL3 vector and transfected into HEK-293 AD cells. The full-length inserts are reported in the Supplemental Methods. 24h post-transfection, RNA was extracted and RT-PCR was performed using vector-specific primers (SD6 + SA2). The three synonymous variants are given in blue, the missense variant is given in green, and the remaining five intronic variants are given in black. None of these variants affected splicing. The effect on splicing of the missense c.92T>A (p.Phe31Tyr) and the synonymous c.2145C>A (p.Thr715=) SNVs was also determined in lymphocytes-extracted RNA, which confirmed the minigene outcomes (not shown). The missense c.92T>A (p.Phe31Tyr) in exon 2 was tested because it is located 3 bp downstream of a splice acceptor site and was predicted to activate a cryptic splice donor site. In all exon 2 constructs, the upper band represents normal splicing, while the lower band represents non-specific splicing. All bands were confirmed by sequencing. MW, molecular weight marker; MOCK, cDNA from empty vector-transfected cells consisting of a 260 bp band made up of fragments of pSPL3 exons; WT, wild-type; --, negative control (RT-PCR without cDNA). The lines separating c.1614-32G>A and negative control from exons 18-20 constructs indicate that data for intervening samples were removed. (C) The c.195G>T (p.Gly65=) variant was subsequently studied by RT-qPCR to assess whether it affects *PAM* mRNA expression. HEK-293 AD cells were transfected with WT, mutant, and WT plus mutant (to mimic a heterozygous state) plasmids. 24h post-transfection, RNA was extracted and RT-qPCR was performed using a TaqMan assay designed to specifically amplify the correctly spliced human *PAM* exon 2 transcript. The variant did not affect *PAM* mRNA expression. WT, wild-type.

### **Supplemental Tables**

**Table S2. Primers used in this study**

| **Application** | **Primer name** | **Sequence (5’-3’)** | **Product size (bp)** |
| --- | --- | --- | --- |
| Sanger sequencing  screening | PAM_prom1_F | GGGGCAGCTAAGATTCTCCC | 732 |
|  | PAM_prom1_R | GCAAATCCCAGCTGAGGTTC |  |
|  | PAM_prom2_F | GGCTGTGAGGGATAAAGGGG | 699 |
|  | PAM_prom2_R | ACAAGGAGAACTAGCAGGCT |  |
|  | PAM_ex1_F | GCCATGAAGTAGCGGCTG | 233 |
|  | PAM_ex1_R | CAACAGCATGCACAGAAAGC |  |
|  | PAM_ex2_F | CCTTTCTTTCCCCTTCTCATGT | 285 |
|  | PAM_ex2_R | AGTTCCTACAGCTGTAAATGCA |  |
|  | PAM_ex3_F | GCCACTCAGCCCTCAGAA | 239 |
|  | PAM_ex3_R | ACTGCCCTCCATTAAACTGC |  |
|  | PAM_ex4_F | TGTATTTTATGACTTTGGTGCACAG | 229 |
|  | PAM_ex4_R | GCACAGATACACTTTGCACTTG |  |
|  | PAM_ex5_F | CCCATCACCTCCCACCATG | 270 |
|  | PAM_ex5_R | CAGAACACTGTCAACACTGGT |  |
|  | PAM_ex6_F | ACTCTGGTTCTGAAGTGGGA | 342 |
|  | PAM_ex6_R | TGCTTAGTTCCAACACTTCAAAA |  |
|  | PAM_ex7_F | AGTATGTCTGCAGGAAAGTCAA | 392 |
|  | PAM_ex7_R | CTTCCCACTCTAAGTTCGCA |  |
|  | PAM_ex8_F | AGGAACCATTTAGAGTCTGCCT | 490 |
|  | PAM_ex8_R | ACACAAGGGCCTCAATTCCT |  |
|  | PAM_ex9_F | CCTCAGTCCCTAGCATCTTGT | 246 |
|  | PAM_ex9_R | TTTATGGCACCAGGAAGCAC |  |
|  | PAM_ex10_F | ACTCAGTCTTTGCAGCTCTG | 329 |
|  | PAM_ex10_R | AGCTCAGTTATACAGTTAAGGCA |  |
|  | PAM_ex11_F | ACCTCAACACAGTCAAACATGT | 249 |
|  | PAM_ex11_R | GCTGTCACCTGTAATCATGCC |  |
|  | PAM_ex12_F | TGGCAGATCTAAGGGCTTCA | 386 |
|  | PAM_ex12_R | TGTGACTGGTAGAAGAAGGGA |  |
|  | PAM_ex13_F | ACGGTAATAAGCACAAGACCC | 300 |
|  | PAM_ex13_R | TGAGGCAGATACAGAGAGACA |  |
|  | PAM_ex14_F | CCTATGCTTAAAAGTCTGAGTGC | 519 |
|  | PAM_ex14_R | TGCTAACGTGGATTGTAGCATTT |  |
|  | PAM_ex15_F | GAGGCAATTCGACCTTTCCC | 365 |
|  | PAM_ex15_R | ACACAAGACTTCCTCTCTGCT |  |
|  | PAM_ex16_F | TGTGAATGGTCTGCATTTCTCA | 291 |
|  | PAM_ex16_R | GTGCAGTTATACAAAACACTCGA |  |
|  | PAM_ex17_F | CTGTCACATATGCCTTGAGAGT | 208 |
|  | PAM_ex17_R | ACCCATACAACTCCAGAGCT |  |
|  | PAM_ex18_F | ACCATGAAAAGTAGGTAAGGCT | 297 |
|  | PAM_ex18_R | ACTGGCTAAGAATTACAACTGGA |  |
|  | PAM_ex19_F | AGAGAGTCAAACTTTGGGTGAT | 497 |
|  | PAM_ex19_R | TTTTAACCCCTTACAAACAGGTG |  |
|  | PAM_ex20_F | TTGTAGGGACCAGGTTGACC | 372 |
|  | PAM_ex20_R | ACACTCCCATCAACCTTGCT |  |
|  | PAM_ex21_F | TGTTGTGGGCTTTGCTTTGT | 424 |
|  | PAM_ex21_R | AGACAGTACTTCAGGCATCAAGA |  |
|  | PAM_ex22_F | TCCAGGATTCATGGACCACC | 286 |
|  | PAM_ex22_R | AGAATTCAACACAGCCAGACT |  |
|  | PAM_ex23_F | ATTTGACTGGGTAGGGGTGG | 366 |
|  | PAM_ex23_R | TCCCAATCAAACTCCTGAGGA |  |
|  | PAM_ex24_F | GAGCCAGGCCTTGCTTTATC | 223 |
|  | PAM_ex24_R | TGTGCTTGTTGACTTTGAACCA |  |
|  | PAM_ex25_F | CTTTCCCATCCCCACCTTCT | 397 |
|  | PAM_ex25_R | GTCCCACACAGACTAGTACAG |  |
| LOH | PAM_ex16LOH_F | TGTGAATGGTCTGCATTTCTCA | 173 |
|  | PAM_ex16LOH_R | GGAGTACTGCAGCATTATTTGG |  |
| Mutagenesis | PAM_V27I_F | CCACCTCTTAAAGATAGAAAGTGGGCTTCGGAAAG |  |
|  | PAM_V27I_R | CTTTCCGAAGCCCACTTTCTATCTTTAAGAGGTGG |  |
|  | PAM_F31Y_F | AAATGGTCTGGTAGTTTCTTTATACCTCTTAAAGACAGAAAGTGG |  |
|  | PAM_F31Y_R | CCACTTTCTGTCTTTAAGAGGTATAAAGAAACTACCAGACCATTT |  |
|  | PAM_V49L_F | TGAATCAATAGGAACTAGGGGTCTGGTGGTACCA |  |
|  | PAM_V49L_R | TGGTACCACCAGACCCCTAGTTCCTATTGATTCA |  |
|  | PAM_R80Q_F | CCTCATCCACTGGTATTTGCATAGACATGCAGAAG |  |
|  | PAM_R80Q_R | CTTCTGCATGTCTATGCAAATACCAGTGGATGAGG |  |
|  | PAM_T189I_F | GTGGCAGACGTATGAGGTGTAAGGACACACCAG |  |
|  | PAM_T189I_R | CTGGTGTGTCCTTACACCTCATACGTCTGCCAC |  |
|  | PAM_H240Y_F | CACTTACTACCTTACCTAAATAGTGAGTGTGAACTCTATAGGC |  |
|  | PAM_H240Y_R | GCCTATAGAGTTCACACTCACTATTTAGGTAAGGTAGTAAGTG |  |
|  | PAM_V244A_F | TTACTCTGTATCCACTTACTGCCTTACCTAAATGGTGAGTG |  |
|  | PAM_V244A_R | CACTCACCATTTAGGTAAGGCAGTAAGTGGATACAGAGTAA |  |
|  | PAM_P412L_F | CTCTGATTCTGCCTTTTCTGTAAGATTATATTTGTGCACATGAAC |  |
|  | PAM_P412L_R | GTTCATGTGCACAAATATAATCTTACAGAAAAGGCAGAATCAGAG |  |
|  | PAM_I452V_F | TTCTGTCTCTGACAAGAACAGCATTACCCCTCTCATG |  |
|  | PAM_I452V_R | CATGAGAGGGGTAATGCTGTTCTTGTCAGAGACAGAA |  |
|  | PAM_E491D_F | GGAAATCTCCTGTGTGGTCTGGTTCCCAGGTGC |  |
|  | PAM_E491D_R | GCACCTGGGAACCAGACCACACAGGAGATTTCC |  |
|  | PAM_S539W_F | CAAACTTGCTGTCAAACCAGTTTCCATCCCAGACA |  |
|  | PAM_S539W_R | TGTCTGGGATGGAAACTGGTTTGACAGCAAGTTTG |  |
|  | PAM_G552R_F | GTGTCTTCTTCAATTGGTCTGAGTCCTATTTGCTGGTAA |  |
|  | PAM_G552R_R | TTACCAGCAAATAGGACTCAGACCAATTGAAGAAGACAC |  |
|  | PAM_D563G_F | GTACTGCAGCATTATTTGGACCTATGACAAGAATAGTGTCTTCTTC |  |
|  | PAM_D563G_R | GAAGAAGACACTATTCTTGTCATAGGTCCAAATAATGCTGCAGTAC |  |
|  | PAM_G641R_F | GGCTCCAGTGCGTGGATCCACAGCC |  |
|  | PAM_G641R_R | GGCTGTGGATCCACGCACTGGAGCC |  |
|  | PAM_R703Q_F | GGATCCGACCATTTTCCTGGTCTGCCACACATAAT |  |
|  | PAM_R703Q_R | ATTATGTGTGGCAGACCAGGAAAATGGTCGGATCC |  |
|  | PAM_F759S_F | GAACCTGTACAAGGATCTGTGATGAACTTTTCC |  |
|  | PAM_F759S_R | GGAAAAGTTCATCACAGATCCTTGTACAGGTTC |  |
|  | PAM_P51=_F | CAAAATCTGATGAATCAATGGGAACTACGGGTCTGGTG |  |
|  | PAM_P51=_R | CACCAGACCCGTAGTTCCCATTGATTCATCAGATTTTG |  |
|  | PAM_G65=_F | GACTGTTTAGGTGTAACACCAGGCATGCGAATATC |  |
|  | PAM_G65=_R | GATATTCGCATGCCTGGTGTTACACCTAAACAGTC |  |
|  | PAM_T715=_F | CTTAATCTCTCTCACAAATTCTTTTGTGTCAGTTTTAAAACACTGGATC |  |
|  | PAM_T715=_R | GATCCAGTGTTTTAAAACTGACACAAAAGAATTTGTGAGAGAGATTAAG |  |
|  | PAM_c.92T>A_F | AAAATGGTCTGGTAGTTTCTTTATACCTTAAAAAAGAGGGCAATTTC |  |
|  | PAM_c.92T>A_R | GAAATTGCCCTCTTTTTTAAGGTATAAAGAAACTACCAGACCATTTT |  |
|  | PAM_c.443-79G>T_F | GTTAATCTCAAATTGCTGTTCTATGAAATCCAAAGGGAAATAACATATAACATAT |  |
|  | PAM_c.443-79G>T_R | ATATGTTATATGTTATTTCCCTTTGGATTTCATAGAACAGCAATTTGAGATTAAC |  |
|  | PAM_c.443-5T>C_F | AACTCTGAATCCAACACCTAAGGATTTACAAAAAAGTTAATATATTAGAAATGAGC |  |
|  | PAM_c.443-5T>C_R | GCTCATTTCTAATATATTAACTTTTTTGTAAATCCTTAGGTGTTGGATTCAGAGTT |  |
|  | PAM_c.1614-32G>A_F | GCATTAAATAGGACATGAAATAATCATAAATTCCAATGAAATTGAGAAATGCAGAC |  |
|  | PAM_c.1614-32G>A_R | GTCTGCATTTCTCAATTTCATTGGAATTTATGATTATTTCATGTCCTATTTAATGC |  |
|  | PAM_c.1804-5A>T_F | CAGTTTGAACACCTTTATAAAAAGGAACAAAAGCCTTACCTACTTT |  |
|  | PAM_c.1804-5A>T_R | AAAGTAGGTAAGGCTTTTGTTCCTTTTTATAAAGGTGTTCAAACTG |  |
|  | PAM_c.2331+76A>C_F | TCTTAAAAGTTTTTCTAAAGCTATTCAAGGACAGATTGAATGCCAAGATTTATATTTTA |  |
|  | PAM_c.2331+76A>C_R | TAAAATATAAATCTTGGCATTCAATCTGTCCTTGAATAGCTTTAGAAAAACTTTTAAGA |  |
|  | PAM_c.2332-2A>T_F | CATGAGGCATATCAAAGTGCAGCCAAAATAAGAAGCTACAC |  |
|  | PAM_c.2332-2A>T_R | GTGTAGCTTCTTATTTTGGCTGCACTTTGATATGCCTCATG |  |
|  | PAM_c.2746+3A>T_F | AGGTCATAGGCAAACCTCTAAATCTTCCCAGTACTC |  |
|  | PAM_c.2746+3A>T_R | GAGTACTGGGAAGATTTAGAGGTTTGCCTATGACCT |  |
|  | PAM_c.-607A>G_F | GCTCTTCCTCCCTCGGACCCACTCCTG |  |
|  | PAM_c.-607A>G_R | CAGGAGTGGGTCCGAGGGAGGAAGAGC |  |
|  | PAM_c.-550G>T_F | AAGATTTCAGGAACACCCACGGCGCAGCC |  |
|  | PAM_c.-550G>T_R | GGCTGCGCCGTGGGTGTTCCTGAAATCTT |  |
|  | PAM_c.-361G>A_F | AAACCCGCGTCAGTTATCATTCAGAACCTGCAAAAATAG |  |
|  | PAM_c.-361G>A_R | CTATTTTTGCAGGTTCTGAATGATAACTGACGCGGGTTT |  |
|  | PAM_c.-133T>C_F | CTGCGGCGGCCGCACACCATCCG |  |
|  | PAM_c.-133T>C_R | CGGATGGTGTGCGGCCGCCGCAG |  |
|  | PAM_c.-109G>C_F | GCCCGGGGACGGCGGGCGTCC |  |
|  | PAM_c.-109G>C_R | GGACGCCCGCCGTCCCCGGGC |  |
|  | PAM_c.-35C>G_F | GGGAGAGGACCCGGCAGCGCAGC |  |
|  | PAM_c.-35C>G_R | GCTGCGCTGCCGGGTCCTCTCCC |  |
| Cloning | PAM_ex2_pSPL3_F | GGGATCACCAGAATTCCAATCATCTTTAAGGTCATAGAATT | 321 |
|  | PAM_ex2_pSPL3_R | TCGAGCTCCAGAATTCACGTTTAAAGAAGTTCCTACAGCTG |  |
|  | PAM_ex6_pSPL3_F | GGGATCACCAGAATTCTTATCAGTCAATGGTATATTTCTGT | 384 |
|  | PAM_ex6_pSPL3_R | TCGAGCTCCAGAATTCAGTTCCAAATTAAAGGAAACAAAAG |  |
|  | PAM_ex16_pSPL3_F | GGGATCACCAGAATTCTTGAGCTCTTTTGATTTTGTATTTA | 317 |
|  | PAM_ex16_pSPL3_R | TCGAGCTCCAGAATTCAGTGCAGTTATACAAAACACTCGAA |  |
|  | PAM_ex18_pSPL3_F | GGGATCACCAGAATTCGCTTCTACTCCAATTGCTTG | 411 |
|  | PAM_ex18_pSPL3_R | TCGAGCTCCAGAATTCGGACAATTACAGCTTAAAGAA |  |
|  | PAM_ex19_pSPL3_F | GGGATCACCAGAATTCCATTTAATGGTTTACTATCATGAAT | 401 |
|  | PAM_ex19_pSPL3_R | TCGAGCTCCAGAATTCTAGCAAAGAAAATTTATAAAAAGAG |  |
|  | PAM_ex20_pSPL3_F | GGGATCACCAGAATTCAGACACTAAGTATTTTATATTCTAC | 416 |
|  | PAM_ex20_pSPL3_R | TCGAGCTCCAGAATTCAAATAGAACACTCCCATCAACCTTG |  |
|  | PAM_ex21_pSPL3_F | GGGATCACCAGAATTCTTTGTTGTTTTGTTGTGGGCTTTGC | 300 |
|  | PAM_ex21_pSPL3_R | TCGAGCTCCAGAATTCATCAGTAAAAAGATAAGGAAATGCA |  |
|  | PAM_ex24_pSPL3_F | GGCAGGAGTCAAACCCAGC | 554 |
|  | PAM_ex24_pSPL3_R | GACATGCCAACAGGAGTGACAG |  |
|  | PAM_3’UTR_psiCHECK2_F | TAGGCGATCGCTCGAGAAACCAAGCTTTGATTTAGATTGAG | 2040 |
|  | PAM_3’UTR_psiCHECK2_R | TTGCGGCCAGCGGCCGCTTTCCCTTTGCTTTTTTAATAAGTC |  |
|  | PAM_CDS_pCMV-FLAG-C_F | AGATCTGCCGCCGCGATCGCATGGCTGGCCGCGTCCCTAGCCTGC | 2922 |
|  | PAM_CDS_pCMV-FLAG-C_R | GCGGCCGCGTACGCGTGGAGGAGGAAGGTGCGAGCGCAGGC |  |
| Expression constructs sequencing | PAM_seq_F1 | AGGAAGCCTTCGTGATTGACT |  |
|  | PAM_seq_F2 | GGACACTGATTGGACGGCA |  |
|  | PAM_seq_F3 | GACCTGGTAGCTGAGATTGCA |  |
|  | PAM_seq_F4 | TGGGAATTATTGGGTCACAGAC |  |
|  | PAM_seq_F5 | GGTTATTCCGGTGGTTGTCC |  |
|  | PAM_seq_R1 | GGAACTACGGGTCTGGTGGT |  |
|  | PAM_seq_R2 | CTTCTCGTTTTGGCTGCTGT |  |
|  | PAM_seq_R3 | TGGTCCGAGTCCTATTTGCTG |  |
|  | PAM_seq_R4 | TTTCCTGGACCTCAATGCCA |  |
| qPCR | PAM_ex2_norm splic_F | GATTCATCAGATTTTGCATTGGAT | 79 |
|  | PAM_ex2_norm splic_R | GGGTCCCCTCGGGAGAT |  |
|  | PAM_ex2_norm splic_probe | TTACACCTAAACAGACCTGG |  |
| RT-PCR | eGFP_F | ATCATGGCCGACAAGCAGAA | 187 |
|  | eGFP_R | TCTCGTTGGGGTCTTTGCTC |  |
| PCR splicing to delete exon 21 | PAM_flank-ex21_F | GACCTGGTAGCTGAGATTGCA | 1086 |
|  | PAM_ex21del_R | ATCGATGTTCCAATTCTTGCGCACTGGCTT |  |
|  | PAM_ex21del_F | AAGCCAGTGCGCAAGAATTGGAACATCGAT | 63 |
|  | PAM_flank-ex21_R | TTTCCTGGACCTCAATGCCA |  |
| Minigene assay | SD6 | TCTGAGTCACCTGGACAACC |  |
|  | SA2 | ATCTCAGTGGTATTTGTGAGC |  |

**Table S3. Candidate variants in index family**

| **Gene** | **Chromosome** | **DNA**  **change** | **Protein change** | **RefSeq** | **SNP ID** | **Global Control MAF (%)** | ***In silico* prediction** |
| --- | --- | --- | --- | --- | --- | --- | --- |
| *ITGB5* | 3 | c.1262C>T | p.Thr421Met | NM_002213.5 | rs200415084 | 0.0039 | 7, 9, 2 |
| *THBS4* | 5 | c.1771C>T | p.Arg591Trp | NM_003248.6 | rs148589675 | 0.0368 | 2, 6, 9 |
| *AVL9* | 7 | c.243A>C | p.Arg81Ser | NM_015060.3 | rs142055726 | 0.1156 | 0, 7, 10 |
| *CEP350* | 1 | c.6130A>C | p.Thr2044Pro | NM_014810.5 | rs56173179 | 0.3654 | 2, 11, 3 |
| *ANKRD36C* | 2 | c.3910A>T | p.Asn1304Tyr | NM_001310154.3 | rs768682466 | 0.2027 | 0, 4, 9 |
| ***PAM*** | **5** | **c.2108G>A** | **p.Arg703Gln** | **NM_000919.3** | **rs761130902** | **0.0013** | **10, 9, 0** |
| *TRMT11* | 6 | c.869A>G | p.Asp290Gly | NM_001031712.2 | rs781238788 | 0.0007 | 10, 6, 2 |
| *PXDNL* | 8 | c.3352C>G | p.Leu1118Val | NM_144651.5 | rs145542518 | 0.3009 | 0, 4, 13 |
| *UHRF2* | 9 | c.869C>G | p.Ser290Cys | NM_152896.3 | rs373417432 | 0.0059 | 1, 8, 8 |
| *ZFYVE19* | 15 | c.1192C>T | p.Arg398Cys | NM_001077268.2 | rs72735636 | 0.1887 | 2, 1, 14 |
| *ZNF799* | 19 | c.1481A>G | p.His494Arg | NM_001080821.3 | rs151038912 | 0.1275 | 4, 5, 7 |
| *ABCB6* | 2 | c.1762G>A | p.Gly588Ser | NM_005689.4 | rs145526996 | 0.4192 | 5, 12, 0 |
| *ARAP2* | 4 | c.2050G>A | p.Asp684Asn | NM_015230.3 | rs200497411 | 0.0112 | 2, 5, 10 |
| *RPS3A* | 4 | c.598G>C | p.Ala200Pro | NM_001006.5 | rs781597345 | 0.0007 | 4, 9, 4 |
| *CCNO* | 5 | c.134G>T | p.Pro45His | NM_021147.5 | rs139606873 | 0.1459 | 0, 4, 13 |
| *SEPTIN14* | 7 | c.796T>C | p.Tyr266His | NM_207366.2 | rs368110863 | 0.0158 | 8, 5, 4 |
| *FBXO24* | 7 | c.17T>A | p.Val6Asp | NM_033506.3 | rs148663467 | 0.0269 | 0, 3, 14 |
| *CFTR* | 7 | c.2168G>T | p.Gly723Val | NM_000492.4 | rs200531709 | 0.0013 | 2, 10, 6 |
| *OR2A14* | 7 | c.736G>A | p.Val246Met | NM_001001659.1 | rs201108825 | 0.0177 | 1, 4, 12 |
| *CD163L1* | 12 | c.3163G>A | p.Gly1055Ser | NM_174941.5 | rs36206713 | 0.6261 | 1, 7, 9 |
| *CFAP251* | 12 | c.2578C>T | p.Arg860Cys | NM_001178003.1 | rs146415200 | 0.5433 | 2, 4, 10 |
| *SBNO1* | 12 | c.614A>G | p.Asn205Ser | NM_018183.3 | rs199607415 | 0.1925 | 0, 8, 8 |
| *ATP12A* | 13 | c.1883G>A | p.Arg628Gln | NM_001185085.2 | rs770792516 | 0.0013 | 5, 9, 4 |
| *HIF1A* | 14 | c.148G>C | p.Val50Leu | NM_181054.2 | rs61755705 | 0.2135 | 0, 13, 4 |
| *NRDE2* | 14 | c.1519G>A | p.Val507Met | NM_017970.3 | rs79341977 | 0.4136 | 2, 9, 5 |
| *GATM* | 15 | c.67G>A | p.Arg23Trp | NM_001482.3 | rs1240389133 | 0.0013 | 0, 10, 9 |
| *LIPC* | 15 | c.1214C>T | p.Thr405Met | NM_000236.2 | rs113298164 | 0.2839 | 5, 7, 5 |

Twenty-seven nonsynonymous variants passed the variant prioritization process in the individuals screened by WES. In light yellow are given variants that fully segregated with the phenotype in all three generations, in light green are given variants that fully segregated with the phenotype only in generation II, and in light orange are given variants that fully segregated with the phenotype only in generation III. The *PAM* variant is highlighted in bold. MAFs were retrieved from the gnomAD database ver. 3.1.2. For *in silico* predictions, the computational verdict was based on the combined outputs of 19 software available in Varsome: the first number represents the pathogenic verdicts, the second number the uncertain verdicts, and the third number the benign verdicts.

MAF, minor allele frequency; n.a., not available; SA, splice acceptor site; VUS, variant of uncertain significance.

**Table S4. *PAM* SNVs detected in our cohort of individuals with pituitary adenomas but not functionally tested**

| **DNA change** | **Protein change** | **SNP ID** | **Location in gene** | **Location in protein** | **Global Control MAF (%)** | **Highest Control Population MAF (%)** | **# of homozygotes in gnomAD** | ***In silico* prediction** |
| --- | --- | --- | --- | --- | --- | --- | --- | --- |
| c.-1069delT | n.a. | rs3834823 | promoter | n.a. | 9.088 | 26.27 (African) | 1560 | benign |
| c.-1009G>T | n.a. | rs1231126774 | promoter | n.a. | 0.0007 | 0.0024 (African) | 0 | benign |
| c.-310C>G | n.a. | rs249496 | 5’UTR (exon 1) | n.a. | 51.34 | 72.21 (African) | 21840 | benign |
| c.89+88G>A | n.a. | rs73192734 | intron 1 | n.a. | 8.920 | 25.33 (African) | 1486 | benign |
| c.211-78A>G | n.a. | rs73175443 | intron 2 | n.a. | 3.012 | 10.40 (African) | 222 | benign |
| c.269-185T>C | n.a. | rs149240405 | intron 3 | n.a. | 0.4472 | 2.083 (Amish) | 5 | benign |
| c.269-92A>G | n.a. | rs6876278 | intron 3 | n.a. | 26.94 | 43.31 (East Asian) | 5911 | benign |
| c.269-51G>T | n.a. | rs112746759 | intron 3 | n.a. | 1.352 | 2.115 (European) | 18 | benign |
| c.443-121A>G | n.a. | rs542812907 | intron 5 | n.a. | 0.0861 | 3.838 (Amish) | 1 (Amish) | benign |
| c.527-69T>A | n.a. | rs3776873 | intron 6 | n.a. | 28.75 | 43.13 (East Asian) | 6539 | benign |
| c.576-145G>A | n.a. | rs143408776 | intron 7 | n.a. | 0.6669 | 1.241 (European) | 8 (European) | benign |
| c.576-68G>A | n.a. | rs140557297 | intron 7 | n.a. | 1.099 | 1.701 (European) | 4 | benign |
| c.576-14A>G | n.a. | rs182339803 | intron 7 | n.a. | 0.6395 | 2.038 (African) | 10 | benign |
| c.906-75G>T | n.a. | rs2195272 | intron 11 | n.a. | 28.65 | 43.10 (East Asian) | 6510 | benign |
| c.1483+26G>A | n.a. | rs3733939 | intron 13 | n.a. | 40.91 | 54.11 (African) | 13363 | benign |
| c.1484-98A>G | n.a. | rs12656131 | intron 14 | n.a. | 31.17 | 43.81 (East Asian) | 7542 | benign |
| c.1614-125A>G | n.a. | rs1451051658 | intron 15 | n.a. | n.a. | n.a. | n.a. | benign |
| c.2014+27G>A | n.a. | rs114859298 | intron 18 | n.a. | 0.1295 | 4.434 (African) | 43 | benign |
| c.2015-171T>C | n.a. | rs45518833 | intron 18 | n.a. | 25.14 | 43.66 (East Asian) | 5278 | benign |
| c.2215+139G>A | n.a. | rs73179842 | intron 19 | n.a. | 3.573 | 12.60 (African) | 358 | benign |
| c.2307A>C | p.Ile769= | rs2230457 | exon 20 | PAL | 25.01 | 43.65 (East Asian) | 5256 | benign |
| c.2431+147G>A | n.a. | rs11960579 | intron 21 | n.a. | 3.875 | 13.55 (African) | 385 | benign |
| c.2431+154T>A | n.a. | rs30719 | intron 21 | n.a. | 3.017 | 7.723 (South Asian) | 76 | benign |
| c.2432-142A>G | n.a. | rs77585944 | intron 21 | n.a. | 1.470 | 4.539 (Finnish) | 27 | benign |
| c.2432-23C>G ^†^ | n.a. | rs183956071 | intron 21 | n.a. | 0.6695 | 3.719 (Finnish) | 15 | benign |
| c.2486-125A>C | n.a. | rs79989892 | intron 22 | n.a. | 1.081 | 3.681 (African) | 22 (African) | benign |
| c.2486-88A>C | n.a. | rs17296280 | intron 22 | n.a. | 27.44 | 43.83 (Amish) | 6177 | benign |
| c.2689+46G>C | n.a. | rs41300805 | intron 23 | n.a. | 7.290 | 11.38 (Finnish) | 1532 | benign |
| c.2689+118C>T | n.a. | rs147272823 | intron 23 | n.a. | 1.313 | 9.226 (Amish) | 15 | benign |
| c.2690-28C>T | n.a. | rs26433 | intron 23 | n.a. | 64.72 | 84.54 (South Asian) | 32347 | benign |
| c.2747-49_2747-48insAT | n.a. | rs2067133 | intron 24 | n.a. | 74.80 | 86.52 (African) | 43021 | benign |
| c.*417G>A | n.a. | rs5855 | 3’UTR (exon 25) | n.a. | 68.90 | 84.65 (South Asian) | 36249 | benign |
| c.*941T>A | n.a. | rs26432 | 3’UTR (exon 25) | n.a. | 68.89 | 84.73 (South Asian) | 36237 | benign |
| c.*1025G>C | n.a. | rs26431 | 3’UTR (exon 25) | n.a. | 74.60 | 85.72 (African) | 42792 | benign |
| c.*1163G>A | n.a. | rs26430 | 3’UTR (exon 25) | n.a. | 68.90 | 84.71 (South Asian) | 36246 | benign |
| c.*1284G>A | n.a. | rs26429 | 3’UTR (exon 25) | n.a. | 68.91 | 84.69 (South Asian) | 36256 | benign |
| c.*1715C>T | n.a. | rs26428 | 3’UTR (exon 25) | n.a. | 68.90 | 84.66 (South Asian) | 36271 | benign |
| c.*1748A>C | n.a. | rs73181445 | 3’UTR (exon 25) | n.a. | 1.624 | 5.710 (African) | 63 | benign |
| c.*2014A>T | n.a. | rs26427 | 3’UTR (exon 25) | n.a. | 4.170 | 14.57 (African) | 441 | benign |

In addition to the prioritized variants, 38 other variants (10 in regulatory regions, one synonymous, and 27 intronic) were detected in our cohort. Variants were annotated using the NC_000005.9(NM_000919.3) reference sequence. MAFs were retrieved from the gnomAD database. “European” excludes the Finnish population; “African” includes African-American; “Latino” includes Admixed American. For *in silico* predictions, the computational verdict was based on the output reported in Varsome.

MAF, minor allele frequency; n.a., not applicable.

^†^The subject harbors the 5’UTR c.-361G>A *PAM* variant.

**Table S5. Spatial clustering analysis**

| **Domain** | **Variants #** | **p-value** |
| --- | --- | --- |
| N-terminus | 2 | 0.02 |
| PHMcc | 4 | 0.63 |
| linker | 3 | 0.20 |
| PALcc | 7 | 0.12 |
| C-terminus | 0 | 0.94 |

The spatial clustering of a set of variants is described by calculating the geometric mean distance between all pairs of variants and normalized to the gene's cDNA length (2,922 bp, isoform P19021-5). An empirical p-value is calculated by randomly-generating 100,000 permutations of the variants and comparing their clustering distance against the clustering of the actual variants. As input, we gave the 16 prioritized variants found in the cohort of persons with pituitary adenomas and reported in Table 2. We then classified the number of variants located in particular domains of the protein (converted to cDNA coordinates) and counted the number of variants appearing in each domain across all the permutations to calculate an empirical p-value comparing the actual number of variants found in each domain to the number of variants in each domain in the 100,000 permutations analysis.

**Table S6. Rare (MAF < 1%) and likely pathogenic *PAM* variants that are significantly associated with diagnoses of tumoral/hyperfunctioning pituitary gland in the UKBB**

| **SNP ID** | **DNA/Protein change** | **Location in protein** | **UKBB MAF (%)** | **CADD_PHRED Score** | **Carriers – No diagnosis** | **Non carriers – No diagnosis** | **Carriers - Diagnosis** | **Non carriers - Diagnosis** | **P-value** | **Diagnosis** |
| --- | --- | --- | --- | --- | --- | --- | --- | --- | --- | --- |
| rs61729214 | p.Val351Met | PHMcc | 0.0095 | 25.00 | 36 | 200300 | 2 | 242 | 1.32E-11 | D352 |
| rs150469595 | p.Ile554Val | PALcc | 0.0813 | 25.20 | 324 | 200201 | 2 | 49 | 8.36E-07 | E220 |
| rs368974389 | c.2746+3A>T | PALcc | 0.0401 | 23.50 | 160 | 200364 | 1 | 50 | 0.0232 | E220 |
| rs150662320 | p.Val597Met | PALcc | 0.0439 | 28.90 | 175 | 200141 | 1 | 50 | 0.0314 | E221 |
| n.a. | p.Pro192Arg | PHMcc | 0.0018 | 23.10 | 6 | 194611 | 1 | 204 | 9.29E-09 | E222 |
| rs145099851 | p.Arg190His | PHMcc | 0.1290 | 19.39 | 516 | 200009 | 1 | 4 | 1.74E-05 | E229 |
| rs772356278 | p.Gly531Cys | PALcc | 0.0002 | 31.00 | 0 | 200519 | 1 | 79 | 3.48E-138 | E237 |
| rs1201940345 | c.2689+34G>A | n.a. | 0.0007 | 15.93 | 2 | 200189 | 1 | 79 | 4.36E-47 | E237 |
| rs61729214 | p.Val351Met | PHMcc | 0.0095 | 25.00 | 37 | 200463 | 1 | 79 | 8.17E-05 | E237 |
| rs770736888 | p.Pro693Leu | PALcc | 0.0160 | 23.40 | 63 | 200459 | 1 | 79 | 0.0029 | E237 |
| n.a. | p.Ser97Arg | PHMcc | 0.0007 | 25.40 | 2 | 200516 | 1 | 57 | 1.95E-64 | E240 |
| COSV57210205 * | p.Leu856Pro | C-ter | 0.0040 | 25.00 | 15 | 200528 | 1 | 57 | 3.23E-13 | E240 |

Variants were annotated using the NC_000005.9(NM_000919.3) reference sequence. MAFs were retrieved from the UKBB database. Only variants with a CADD_PHRED score ≥ 15 and a MAF < 1% are listed. The selected diagnoses are given using the International Classification of Diseases, Tenth Revision (ICD-10) coding: D352, benign neoplasm of pituitary gland; E220, acromegaly and pituitary gigantism; E221, hyperprolactinemia; E222, syndrome of inappropriate secretion of antidiuretic hormone; E229, hyperfunction of the pituitary gland, unspecified; E237, disorder of pituitary gland (unspecified); E240, pituitary-dependent Cushing disease. n.a., not applicable; *, variant ID reported in the COSMIC database.

**Table S7: Gene damage index (GDI) for genes harboring germline and somatic variants known to predispose to pituitary tumors**

|  | **Gene** | **GDI (Phred homogenized)** | **Mean** | **Median** | **Max** | **Min** | **SD** | **95% CI** | **Upper bound (GDI cutoff)** | **Lower bound** |
| --- | --- | --- | --- | --- | --- | --- | --- | --- | --- | --- |
| Germline | *AIP* | 1.160 | 3.829 | 2.161 | 11.375 | 0.196 | 3.991 | 3.8±2.3 | 6.087 | 1.571 |
|  | *MEN1* | 1.434 |  |  |  |  |  |  |  |  |
|  | *CDKN1B* | 9.581 |  |  |  |  |  |  |  |  |
|  | *PRKAR1A* | 0.219 |  |  |  |  |  |  |  |  |
|  | *GPR101* | 11.375 |  |  |  |  |  |  |  |  |
|  | *CABLES1* | 8.032 |  |  |  |  |  |  |  |  |
|  | *DICER1* | 2.685 |  |  |  |  |  |  |  |  |
|  | *SDHA* | 6.694 |  |  |  |  |  |  |  |  |
|  | *SDHB* | 2.675 |  |  |  |  |  |  |  |  |
|  | *SDHC* | 0.196 |  |  |  |  |  |  |  |  |
|  | *SDHD* | 1.647 |  |  |  |  |  |  |  |  |
|  | *MAX* | 0.248 |  |  |  |  |  |  |  |  |
| Somatic | *GNAS* | 3.644 | 2.828 | 2.434 | 5.593 | 0.391 | 1.620 | 2.8±1.1 | 3.950 | 1.705 |
|  | *PIK3CA* | 4.246 |  |  |  |  |  |  |  |  |
|  | *USP8* | 5.593 |  |  |  |  |  |  |  |  |
|  | *USP48* | 2.337 |  |  |  |  |  |  |  |  |
|  | *BRAF* | 1.627 |  |  |  |  |  |  |  |  |
|  | *SF3B1* | 0.391 |  |  |  |  |  |  |  |  |
|  | *ATRX* | 2.251 |  |  |  |  |  |  |  |  |
|  | *NR3C1* | 2.531 |  |  |  |  |  |  |  |  |
| All |  |  | 3.428 | 2.434 | 11.375 | 0.196 | 3.231 | 3.4±1.4 | **4.844** | 2.012 |
|  | ***PAM*** | **3.979** |  |  |  |  |  |  |  |  |

The GDI score for *PAM* is below the upper limit of the calculated confidence interval (GDI cutoff) above which a gene is considered a false positive. GDI scores were retrieved from <http://pec630.rockefeller.edu:8080/GDI/>.

CI, confidence interval; Max, highest GDI within the subgroup; Min, lowest GDI within the subgroup; SD, standard deviation.

**Table S8. *PAM* variants tested to assess their effect on splicing**

| **DNA change** | **Protein change** | **SNP ID** | **Location in gene** | **Location in protein** | **MAF in our cohort (%)** | **Global Control MAF (%)** | **Highest Control Population MAF (%)** | **# of homozygotes in gnomAD** | **P-value** | ***In silico* prediction** | **Individuals harboring the variant** |
| --- | --- | --- | --- | --- | --- | --- | --- | --- | --- | --- | --- |
| c.92T>A | p.Phe31Tyr | rs114014768 | exon 2 | proregion | 0.63 | 0.8654 | 2.814 (African) | 19 | n.s. | activates cryptic SD | 2 acro, 1 gigantism, 1 pediatric CD |
| c.153T>C | p.Pro51= | rs200231533 | exon 2 | PHMcc | 0.16 | 0.0026 | 0.0827 (South Asian) | 0 | <0.05 | benign | 1 gigantism |
| c.195G>T | p.Gly65= | rs145666758 | exon 2 | PHMcc | 0.16 | 0.1908 | 1.995 (Finnish) | 7 | n.s. | benign | 1 acro |
| c.443-79G>T ^†^ | n.a. | rs75448780 | intron 5 | n.a. | 1.56 | 6.366 | 18.20 (East Asian) | 606 | <0.05 | activates cryptic SA | 5 acro, 2 gigantism, 1 NFPA, 2 pediatric CD |
| c.443-5T>C ^†^ | n.a. | rs112626709 | intron 5 | n.a. | 0.16 | 0.4462 | 1.581 (African) | 4 (African) | n.s. | disrupts canonical SA | 1 acro |
| c.1614-32G>A ^†^ | n.a. | rs74633711 | intron 15 | n.a. | 0.15 | 0.4943 | 1.776 (African) | 7 (African) | n.s. | benign | 1 acro |
| c.1804-5A>T | n.a. | rs79621119 | intron 17 | n.a. | 0.31 | 0.7847 | 2.622 (African) | 20 | n.s. | VUS | 2 pediatric CD |
| c.2145C>A | p.Thr715= | rs34746026 | exon 19 | PALcc | 0.46 | 0.2947 | 1.782 (South Asian) | 3 | n.s. | benign | 2 acro, 1 pediatric CD |
| c.2331+76A>C | n.a. | rs139754655 | intron 20 | n.a. | 0.15 | 1.225 | 4.352 (African) | 35 (African) | <0.05 | activates cryptic SA | 1 PRL |

Nine SNVs (three synonymous, one missense, and five intronic) were functionally evaluated for their effect on splicing. Variants were annotated using the NC_000005.9(NM_000919.3) reference sequence. All variants were observed in heterozygosis. MAFs were retrieved from the gnomAD database. When available, combined exome and genome frequencies were used. “European” excludes the Finnish population; “African” includes African-American; “Latino” includes Admixed American. Individual variant allele frequencies in the study population were compared with the global frequencies reported in gnomAD using the Fisher’s exact test or the chi-square test, as appropriate. For *in silico* predictions, the computational verdict was based on the combined outputs of four softwares (Splice Site Finder, MaxEnt, NNSplice, and GeneSplicer) available in Alamut.

acro, acromegaly; CD, Cushing disease; MAF, minor allele frequency; n.a., not applicable; n.s., not significant; PAs, pituitary adenomas; SA, splice acceptor site; SD, splice donor site; VUS, variant of uncertain significance.

^†^One acromegalic harbors the c.443-79G>T and c.443-5T>C variants in the same allele as well as the c.1614-32G>A variant (phase unknown).

### **Supplemental Materials and Methods**

**CLUSTAL Omega v1.2.4 multiple sequence alignment**

```
tr|A0A0S2C767|A0A0S2C767_CHLRE      ---MAPGRL-----FGLLVAVALAL------------------------QAYA---QLPV	25
sp|Q9W1L5|PAL2_DROME                ------------------------------------------------------------	0
sp|O01404|PHM_DROME                 MPRISEIAASVGL-LLLIGVIS-------------VDGLVKE--GDYQNSLYQQNLESNS	44
tr|Q9NJI4|Q9NJI4_APLCA              ---MC-VRAPM----TRLGV--LAVCCVV-SLVMSPTLS--------ADPTTRVMDEVIE	41
sp|P14925|AMD_RAT                   ---MA-GRARSGLLLLLLGLLALQSSCLAFRSPLSVFKRFKETTRSFSNECLGTIGPVTP	56
sp|P19021-5|AMD_HUMAN               ---MA-GRV-----PSLLVLLVFPSSCLAFRSPLSVFKRFKETTRPFSNECLGTTRPVVP	51
tr|A0A2I3SM67|A0A2I3SM67_PANTR      ---MA-GRV-----PSLLVLLVFPSSCLAFRSPLSVFKRFKETTRPFSNECLGTTRPVVP	51

tr|A0A0S2C767|A0A0S2C767_CHLRE      VVSHSIEVNITVPPFKVDQDDAYICVSALLPPHP-HKLVGIIPHAKQEVVHHILLYGCTE	84
sp|Q9W1L5|PAL2_DROME                ------------------------------------------------------------	0
sp|O01404|PHM_DROME                 ATGATASFPFLMPNVSPQTPDLYLCTPIKVDPTTTYYIVGFNPNATMNTAHHMLLYGCGE	104
tr|Q9NJI4|Q9NJI4_APLCA              ADPTTHSMSLLMRGAKPSQPDAYLCTAYPV-TDLETYIYKFQAQANASTAHHMLLYGCDG	100
sp|P14925|AMD_RAT                   LDASDFALDIRMPGVTPKESDTYFCMSMRLPVDEEAFVIDFKPRASMDTVHHMLLFGCNM	116
sp|P19021-5|AMD_HUMAN               IDSSDFALDIRMPGVTPKQSDTYFCMSMRIPVDEEAFVIDFKPRASMDTVHHMLLFGCNM	111
tr|A0A2I3SM67|A0A2I3SM67_PANTR      IDSSDFALDIRMPGVTPKQSDTYFCMSMRIPVDEEAFVIDFKPRASMDTVHHMLLFGCNM	111

tr|A0A0S2C767|A0A0S2C767_CHLRE      PHMASKDGKPVAWRCDMKPV----------CNGPSSTILYGWGRNAPDLRLPEGVGFSVG	134
sp|Q9W1L5|PAL2_DROME                ------------------------------------------------------------	0
sp|O01404|PHM_DROME                 PGTS-K----TTWNCGEMNRASQEESASPCGPHSNSQIVYAWARDAQKLNLPEGVGFKVG	159
tr|Q9NJI4|Q9NJI4_APLCA              PAYSTA----DIWHC-PSVC------------RGQQTILFAWAKNAPPTELPRDVGHRVG	143
sp|P14925|AMD_RAT                   PSST-G----SYWFCDEGTC------------TDKANILYAWARNAPPTRLPKGVGFRVG	159
sp|P19021-5|AMD_HUMAN               PSST-G----SYWFCDEGTC------------TDKANILYAWARNAPPTRLPKGVGFRVG	154
tr|A0A2I3SM67|A0A2I3SM67_PANTR      PSST-G----SYWFCDEGTC------------TDKANILYAWARNAPPTRLPKGVGFRVG	154

tr|A0A0S2C767|A0A0S2C767_CHLRE      ERTGVKYIVAQVHYLKVR----PPDDHSGVTLLLKPHAVPYAAGLVSF-ASWFTIPPGKK	189
sp|Q9W1L5|PAL2_DROME                ------------------------------------------------------------	0
sp|O01404|PHM_DROME                 KNSPIKYLVLQVHYAHIDKFKDGSTDDSGVFLDYTEEPRKKLAGTLLL-GTDGQIPA-MK	217
tr|Q9NJI4|Q9NJI4_APLCA              QRSNVKTLVLQVHYAKG-FVRNESPDHSGIIVHMTDRRPKFVAGIFLMMSTWFQVPPHRE	202
sp|P14925|AMD_RAT                   GETGSKYFVLQVHYGDISAFRDNHKDCSGVSVHLTRVPQPLIAGMYLMMSVDTVIPPGEK	219
sp|P19021-5|AMD_HUMAN               GETGSKYFVLQVHYGDISAFRDNNKDCSGVSLHLTRLPQPLIAGMYLMMSVDTVIPAGEK	214
tr|A0A2I3SM67|A0A2I3SM67_PANTR      GETGSKYFVLQVHYGDISAFRDNNKDCSGVSLHLTRLPQPLIAGMYLMMSVDTVIPAGEK	214


tr|A0A0S2C767|A0A0S2C767_CHLRE      SHPIVNTCCYKGYEALTMFAVRVHTHGLGRRVFMTRETWNKTGT---EELVSRDPQLPQS	246
sp|Q9W1L5|PAL2_DROME                ------------------------------------------------------------	0
sp|O01404|PHM_DROME                 TEHLETACEVNEQKVLHPFAYRVHTHGLGKVVSGYRVRTNSDGEQEWLQLGKRDPLTPQM	277
tr|Q9NJI4|Q9NJI4_APLCA              SYPVDMSCVYLEQKPMYPFAFRTHAHGLGKVITGYL-YN-----GTYQLIGKGNPQWPQA	256
sp|P14925|AMD_RAT                   VVNADISCQYK-MYPMHVFAYRVHTHHLGKVVSGYRVRN-----GQWTLIGRQNPQLPQA	273
sp|P19021-5|AMD_HUMAN               VVNSDISCHYK-NYPMHVFAYRVHTHHLGKVVSGYRVRN-----GQWTLIGRQSPQLPQA	268
tr|A0A2I3SM67|A0A2I3SM67_PANTR      VVNSDISCHYK-NYPMHVFAYRVHTHHLGKVVSGYRVRN-----GQWTLIGRQSPQLPQA	268

tr|A0A0S2C767|A0A0S2C767_CHLRE      FVPTTR-HTIWPGDRLTVTCLFDSSSKTAPVNAGGTHNDEMCNMYTMVYGKTPYL---TM	302
sp|Q9W1L5|PAL2_DROME                ------------------------------------------------------------	0
sp|O01404|PHM_DROME                 FYNTSNTDPIIEGDKIAVRCTMQST-RHRTTKIGPTNEDEMCNFYLMYYVDHGETLNMKF	336
tr|Q9NJI4|Q9NJI4_APLCA              FYPVEDVIEVKPGDSLAARCTYDSTHMDQRVGVGATGSDEMCNFYIMYYTDSSVQRPGAE	316
sp|P14925|AMD_RAT                   FYPVEHPVDVTFGDILAARCVFTGEGRTEATHIGGTSSDEMCNLYIMYYMEAKYALSFMT	333
sp|P19021-5|AMD_HUMAN               FYPVGHPVDVSFGDLLAARCVFTGEGRTEATHIGGTSSDEMCNLYIMYYMEAKHAVSFMT	328
tr|A0A2I3SM67|A0A2I3SM67_PANTR      FYPVEHPVDVSFGDLLAARCVFTGEGRTEATHIGGTSSDEMCNLYIMYYMEAKHAVSFMT	328

tr|A0A0S2C767|A0A0S2C767_CHLRE      CDNNVQDI------RDDSPGALPR------------------------------------	320
sp|Q9W1L5|PAL2_DROME                ---------------------------------------------------------MSR	3
sp|O01404|PHM_DROME                 CFSQGAPYYFWSN-PDSGLHNIPHIEASTL------------------------------	365
tr|Q9NJI4|Q9NJI4_APLCA              CMNDQLPELTGAHFPSSVSQPLPPNPQLEEVASGHHHAH---------------------	355
sp|P14925|AMD_RAT                   CTKNVAPDMFRT-IPAEANIPIPVKPDMVMMHGHHKEAENKEKSALMQQPKQGEEEVLEQ	392
sp|P19021-5|AMD_HUMAN               CTQNVAPDMFRT-IPPEANIPIPVKSDMVMMHEHHKETEYKDKIPLLQQPKREEEEVLDQ	387
tr|A0A2I3SM67|A0A2I3SM67_PANTR      CTQNVAPDMFRT-IPPEANIPIPVKSDMVMMHEHHKETEYKDKIPLLQQPKREEEVL---	384

tr|A0A0S2C767|A0A0S2C767_CHLRE      ------------------------------------------------------------	320
sp|Q9W1L5|PAL2_DROME                LLFVALLAISLGYVASSSSNH-------------LPAGL-----AMDLGP-----GVNLN	40
sp|O01404|PHM_DROME                 ------------------------------------------------------------	365
tr|Q9NJI4|Q9NJI4_APLCA              ------------------HQH---------------------------GMSGGQ------	364
sp|P14925|AMD_RAT                   GDFYSLLSKLLGERED-VHVHKYNPTEKTESGSDLVAEIANVVQKKDLGRSDAREGAEHE	451
sp|P19021-5|AMD_HUMAN               GDFYSLLSKLLGEREDVVHVHKYNPTEKAESESDLVAEIANVVQKKDLGRSDAREGAEHE	447
tr|A0A2I3SM67|A0A2I3SM67_PANTR      ------------------------------------------------------------	384

tr|A0A0S2C767|A0A0S2C767_CHLRE      ------HSTLVVDPMP-------NWRPPAP---------------------AGTP-----	341
sp|Q9W1L5|PAL2_DROME                ERFFDQVRALIKRRLQE-----KGLAKPEQPELAMPLTDDDAVALQNQRSYDNVPLPAAS	95
sp|O01404|PHM_DROME                 ------------------------------------------------------------	365
tr|Q9NJI4|Q9NJI4_APLCA              -G---GVAQVHTTQVHT------T-QTPT----------------TQTPTTQTPTMQEIH	397
sp|P14925|AMD_RAT                   EW---GNAILVRDRIHRFHQLESTLRPAES----------RAFSFQQ--PGEGPWEPEPS	496
sp|P19021-5|AMD_HUMAN               -R---GNAILVRDRIHKFHRLVSTLRPPES----------RVFSLQQPPPGEGTWEPEHT	493
tr|A0A2I3SM67|A0A2I3SM67_PANTR      -----------------------------------------------------------D	385


tr|A0A0S2C767|A0A0S2C767_CHLRE      GD---------ALGAEAVGDATSVTTGPDGTLWVLYRASGVWKSDTFDRKEVITR--KEP	390
sp|Q9W1L5|PAL2_DROME                VP-TPVLVENWPTEQHSFGQVTAVAVDPQGSPVVFHRAERYWDVNTFNESNIYYLIEYGP	154
sp|O01404|PHM_DROME                 ------------------------------------------------------------	365
tr|Q9NJI4|Q9NJI4_APLCA              FDHPLQLNTSWPGVELTVGQVGGVSVDQRGNLYVFHRGSRVWNAASFDIDNNFQF-QDSP	456
sp|P14925|AMD_RAT                   GDFHVEEELDWPGVYLLPGQVSGVALDSKNNLVIFHRGDHVWDGNSFDSKFVYQQRGLGP	556
sp|P19021-5|AMD_HUMAN               GDFHMEEALDWPGVYLLPGQVSGVALDPKNNLVIFHRGDHVWDGNSFDSKFVYQQIGLGP	553
tr|A0A2I3SM67|A0A2I3SM67_PANTR      QDFHMEEALDWPGVYLLPGQVSGVALDPKNNLVIFHRGDHVWDGNSFDSKFVYQQIGLGP	445

tr|A0A0S2C767|A0A0S2C767_CHLRE      VPEAVVLNMNPDTGKILARWGADVFYLPHSISVDQYGNVWVVDVGRHQVLKFDS--KGKQ	448
sp|Q9W1L5|PAL2_DROME                IKENTIYVLDAKTGAIKSGWGSNMFYMPHGLTIDLHGNYWITDVAMHQAFKFKP-FSNKP	213
sp|O01404|PHM_DROME                 ------------------------------------------------------------	365
tr|Q9NJI4|Q9NJI4_APLCA              ITEDVVLVTDSTG-HKIRSFGAGRYFLPHGIQVDHKDNIWLTDVALHQVFKIPA-GSDTP	514
sp|P14925|AMD_RAT                   IEEDTILVIDPNNAEILQSSGKNLFYLPHGLSIDTDGNYWVTDVALHQVFKLDPHSKEGP	616
sp|P19021-5|AMD_HUMAN               IEEDTILVIDPNNAAVLQSSGKNLFYLPHGLSIDKDGNYWVTDVALHQVFKLDPNNKEGP	613
tr|A0A2I3SM67|A0A2I3SM67_PANTR      IEEDTILVIDPNNAAVLQSSGKNLFYLPHGLSIDKDGNYWVTDVALHQVFKLDPNNKEGP	505

tr|A0A0S2C767|A0A0S2C767_CHLRE      LLVVGKDRQPGAGKDKFCKPTQVAVL-RDGSFIVADGYCNSRVVWFDKTGKYIAESGAIK	507
sp|Q9W1L5|PAL2_DROME                LLTIGKRFRPGSSVKHLCKPTSIAVA-TTGEFFIADGYCNSRILKFNAAGKLLRTIPQ--	270
sp|O01404|PHM_DROME                 ------------------------------------------------------------	365
tr|Q9NJI4|Q9NJI4_APLCA              TLTIGHRFQHGEELTFFCKPTDVAVL-SSGEFFVSDGYCNSRVVKFSADGKVIKAWGEKN	573
sp|P14925|AMD_RAT                   LLILGRSMQPGSDQNHFCQPTDVAVEPSTGAVFVSDGYCNSRIVQFSPSGKFVTQWGEES	676
sp|P19021-5|AMD_HUMAN               VLILGRSMQPGSDQNHFCQPTDVAVDPGTGAIYVSDGYCNSRIVQFSPSGKFITQWGEES	673
tr|A0A2I3SM67|A0A2I3SM67_PANTR      VLILGRSMQPGSDQNHFCQPTDVAVDPGTGAIYVSDGYCNSRIVQFSPSGKFITQWGEES	565

tr|A0A0S2C767|A0A0S2C767_CHLRE      ------------AVVHGVLVDECEGLVYVASREGRKVVALDITDTKKRLQLKATYDMAAA	555
sp|Q9W1L5|PAL2_DROME                -----PPEFLSLQVPHAITLLEHLDLLCIADRENMRVVCPKAGLISSHGEGEPAATIQEP	325
sp|O01404|PHM_DROME                 ------------------------------------------------------------	365
tr|Q9NJI4|Q9NJI4_APLCA              LEFGVSPPPGTFDVPHSVTVSEGTGQLCVADRENGRVQCFDLEG-N------FDHLIRHK	626
sp|P14925|AMD_RAT                   --SGSSPRPGQFSVPHSLALVPHLDQLCVADRENGRIQCFKTDTKE------FVREIKHA	728
sp|P19021-5|AMD_HUMAN               --SGSSPLPGQFTVPHSLALVPLLGQLCVADRENGRIQCFKTDTKE------FVREIKHS	725
tr|A0A2I3SM67|A0A2I3SM67_PANTR      --SGSSPLPGQFTVPHSLALVPPLGQLCVADRENGQIQCFKTDTKE------FVREIKHS	617

tr|A0A0S2C767|A0A0S2C767_CHLRE      GHG-QVWALRFGPYGEQLALTWDEGKDAHLVNVRFPTQFWTLPGTAKLSPHDFTLGGAAT	614
sp|Q9W1L5|PAL2_DROME                DLGR-VFGV--AS---------------------FGDIVFAVNGPTS----MLPVRGFTI	357
sp|O01404|PHM_DROME                 ------------------------------------------------------------	365
tr|Q9NJI4|Q9NJI4_APLCA              EFGPRLFAVEVCP---------------------LQGVLYAVNGPAYDGPSDLTVQGFTV	665
sp|P14925|AMD_RAT                   SFGRNVFAISY-----------------------IPGFLFAVNGKPYFGDQ-EPVQGFVM	764
sp|P19021-5|AMD_HUMAN               SFGRNVFAISY-----------------------IPGLLFAVNGKPHFGDQ-EPVQGFVM	761
tr|A0A2I3SM67|A0A2I3SM67_PANTR      SFGRNVFAISY-----------------------IPGLLFAVNGKPHFGDQ-EPVQGFVM	653


tr|A0A0S2C767|A0A0S2C767_CHLRE      ELSGAGDRFFSVYLASVGVACDTKCGA-LHKFVVVPTGFKLPAQWELETAVAVPSKAKDV	673
sp|Q9W1L5|PAL2_DROME                ---------------------DPRSETIIGHWGE----FKNPHSMAVS------------	380
sp|O01404|PHM_DROME                 ------------------------------------------------------------	365
tr|Q9NJI4|Q9NJI4_APLCA              ---------------------DMSSGQLLESWNIP-QGLRNPHDLAVD------------	691
sp|P14925|AMD_RAT                   ---------------------NFSSGEIIDVFKPVRKHFDMPHDIV--------------	789
sp|P19021-5|AMD_HUMAN               ---------------------NFSNGEIIDIFKPVRKHFDMPHDIV--------------	786
tr|A0A2I3SM67|A0A2I3SM67_PANTR      ---------------------NFSNGEIIDIFKPVRKHFDMPHDIV--------------	678

tr|A0A0S2C767|A0A0S2C767_CHLRE      KPGDAKELPHGAVAANHTANVVAKAIAK-GVPAPAKPPAGEEEDEGGAMVADDYDEDVED	732
sp|Q9W1L5|PAL2_DROME                -VN-GSALYVTEIGTNHQTNRVWKYVLA--------------------------------	406
sp|O01404|PHM_DROME                 ------------------------------------------------------------	365
tr|Q9NJI4|Q9NJI4_APLCA              -PTTCGSVYVGE----LNPRVVWKLTRADRSTTPTPH-LHM-------------------	726
sp|P14925|AMD_RAT                   -ASEDGTVYIGD----AHTNTVWKFTLTEKMEHRSVKKAGI-------------------	825
sp|P19021-5|AMD_HUMAN               -ASEDGTVYIGD----AHTNTVWKFTLTEKLEHRSVKKAGI-------------------	822
tr|A0A2I3SM67|A0A2I3SM67_PANTR      -ASEDGTVYIGD----AHTNTVWKFTLTEKLEHRSVKKAGI-------------------	714

tr|A0A0S2C767|A0A0S2C767_CHLRE      DTEDAEDYENTKKVLDEEMEAEEEAMADYEEMITGVRPNTTHDKGELVPLDKERYQVLMQ	792
sp|Q9W1L5|PAL2_DROME                ------------------------------------------------------------	406
sp|O01404|PHM_DROME                 ------------------------------------------------------------	365
tr|Q9NJI4|Q9NJI4_APLCA              ---DTQIVNNNKKN--Q--------------------------IGQAGPSD----TQ---	748
sp|P14925|AMD_RAT                   ---EVQEIKEAEAVVEP--------------------------KVENKPTSSE--LQKMQ	854
sp|P19021-5|AMD_HUMAN               ---EVQEIKEAEAVVET--------------------------KMENKPTSSE--LQKMQ	851
tr|A0A2I3SM67|A0A2I3SM67_PANTR      ---EVQEIKEAEAVVET--------------------------KMENKPTSSE--LQKMQ	743

tr|A0A0S2C767|A0A0S2C767_CHLRE      DKKAATKTMGSGWANVLVVVLVLALTVISVYYARATLMHYIQSITVSSSNPGGSGAGIKG	852
sp|Q9W1L5|PAL2_DROME                ------------------------------------------------------------	406
sp|O01404|PHM_DROME                 ------------------------------------------------------------	365
tr|Q9NJI4|Q9NJI4_APLCA              ------------------------------------------------------------	748
sp|P14925|AMD_RAT                   EKQKLSTEPGSGVSVVLITTLLVIPV--LVLLAIVMFIRWKKSRAFGD------H-DR--	903
sp|P19021-5|AMD_HUMAN               EKQKLIKEPGSGVPVVLITTLLVIPV--VVLLAIAIFIRWKKSRAFGA------DSEH--	901
tr|A0A2I3SM67|A0A2I3SM67_PANTR      EKQKLIKEPGSGVPVVLITTLLVIPV--VVLLAIAIFIRWKKSRAFGA------DSEH--	793

tr|A0A0S2C767|A0A0S2C767_CHLRE      GPSAGASTPAGGGLRKTFGAWASAAEGLLGRFSGRGGAAAAAGLAGTTGAG---------	903
sp|Q9W1L5|PAL2_DROME                ------------------------------------------------------------	406
sp|O01404|PHM_DROME                 ------------------------------------------------------------	365
tr|Q9NJI4|Q9NJI4_APLCA              ------------------------------------------------------------	748
sp|P14925|AMD_RAT                   -------------------KLESSSGRVLGRFRGKGSGGLNLGNFFASRKGYSRKGFDRV	944
sp|P19021-5|AMD_HUMAN               -------------------KLETSSGRVLGRFRGKGSGGLNLGNFFASRKGYSRKGFDRL	942
tr|A0A2I3SM67|A0A2I3SM67_PANTR      -------------------KLETSSGRVLGRFRGKGSGGLNLGNFFASRKGYSRKGFDRL	834


tr|A0A0S2C767|A0A0S2C767_CHLRE      ---GAKRSTAEVEAARERERL----LRSGP---	926
sp|Q9W1L5|PAL2_DROME                ---------------------------------	406
sp|O01404|PHM_DROME                 ---------------------------------	365
tr|Q9NJI4|Q9NJI4_APLCA              ---------------------------------	748
sp|P14925|AMD_RAT                   STEGSDQEKDEDDGTESEEEYSAPLPKPAPSS-	976
sp|P19021-5|AMD_HUMAN               STEGSDQEK-EDDGSESEEEYSAPLPALAPSSS	974
tr|A0A2I3SM67|A0A2I3SM67_PANTR      STEGSDQEK-EDDGSESEEEYSAPLPALAPSSS	866
```

The PHM and PAL catalytic cores are underlined underneath the human sequence. All missense human variants reported in the study are given in bold. 15 SNVs identified in our cohort of individuals with pituitary adenomas are given in red with light blue highlight, while seven missense SNVs identified in the UKBB are given in dark blue. APLCA, Aplysia californica; CHLRE, *Chlamydomonas reinhardtii*; DROME, *Drosophila melanogaster*; PANTR, *Pan troglodytes*.

***PAM* PCR products cloned into the pSPL3 minigene vector**

Exons are given in uppercase and introns in lowercase

*PAM*-specific primer sequences are underlined

Tested SNVs are given in red

**PAM exon 2 and 100 bp of flanking intronic sequences**

PCR product: 321 bp

SNVs: c.92T>A, c.153T>C, c.195G>T

caatcatctttaaggtcatagaattcctttctttccccttctcatgttgagtttagattccattatgttcaagaatattgaaattgccctcttttttaaggtttaaagaaactaccagaccattttccaatgaatgtcttggtaccaccagacccgtagttcctattgattcatcagattttgcattggatattcgcatgcctggggttacacctaaacaggtgagaggaattttgtctttcattattcacttgttctggatacaatctataattaaagtaaggaaactgcatttacagctgtaggaacttctttaaacgt

**PAM exon 6 and 150 bp of flanking intronic sequences**

PCR product: 384 bp

SNVs: c.443-79G>T, c.443-5T>C

ttatcagtcaatggtatatttctgttttcatataatataattttatatgttatatgttatttccctttggagttcatagaacagcaatttgagattaactccattttagatgctcatttctaatatattaacttttttgtaaatctttagGTGTTGGATTCAGAGTTGGAGGAGAGACTGGAAGTAAATACTTTGTACTACAGGTACACTATGGGGATATTAGTGCTTTTAGAGgtaagttttgaagtgttggaactaagcaaaacttctagtactatattgtttaaaatacatgcacaaactattatctacatcttaaaaagaattgcccacaccctctgcccactaggggataacagcttttgtttcctttaatttggaact

**PAM exon 16 and 100 bp of flanking intronic sequences**

PCR product: 317 bp

SNVs: c.1614-43A>G, c.1614-32G>A

ttgagctcttttgattttgtatttatgtgaatggtctgcatttctcaatttcattggaatttatgattgtttcatgtcctatttaatgctttttgtttagctcgtttgacagcaagtttgtttaccagcaaataggactcggaccaattgaagaagacactattcttgtcatagatccaaataatgctgcagtactccagtccagtggaaaaaatctgtgagttaaatgacttatgttgttaagacttgtactacatatattgagtataaagtgtatcatagagtcttgaaattcgagtgttttgtataactgcact

**PAM exon 18 and 100 bp of flanking intronic sequences**

PCR product: 411 bp

SNV: c.1804-5A>T

gcttctactccaattgcttgtgaaagtggtcttagtcatgtgggtgtaagtcactaataaccatgaaaagtaggtaaggcttttgttcctttttaaaaagGTGTTCAAACTGGATCCAAACAATAAAGAAGGCCCTGTATTAATCCTGGGAAGGAGCATGCAACCAGGCAGTGACCAGAATCACTTCTGTCAACCCACTGATGTGGCTGTGGATCCAGGCACTGGAGCCATTTATGTATCAGATGGTTACTGCAACAGCAGGATTGTGCAGTTTTCACCAAGTGGAAAGTTCATCACACAGTGGGGAGAAGgtacccaataagactcttaatctccagttgtaattcttagccagtatcactgggaactaaatattggccaacactcacattctttaagctgtaattgtcc

**PAM exon 19 and 100 bp of flanking intronic sequences**

PCR product: 401 bp

SNV: c.2145C>A

catttaatggtttactatcatgaatttagaaacatactagagagtcaaactttgggtgatttttaacattgtgtatctaaggcttttttttgttctgcagAgtcttcagggagcagtcctctgccaggccagttcactgttcctcacagcttggctcttgtgcctcttttgggccaattatgtgtggcagaccgggaaaatggtcggatccagtgttttaaaactgacaccaaagaatttgtgagagagattaagcattcatcatttggaagaaatgtatttgcaatttcatatataccaggtatttcatcttaatatgtttgttgtcttctgtcctactgtactctatccttaaaaattgtgttatcttaatgtgctctttttataaattttctttgcta

**PAM exon 20 and 150 bp of flanking intronic sequences**

PCR product: 416 bp

SNV: c.2331+76A>C

agacactaagtattttatattctacttttgttgccaattgtagggaccaggttgaccctaaatttattctttgcataatatacatggatatattagaagtccattcttacccatattttaaagaaaggttaactggatttgctgttgcagGCTTGCTCTTTGCAGTGAATGGGAAGCCTCATTTTGGGGACCAAGAACCTGTACAAGGATTTGTGATGAACTTTTCCAATGGGGAAATTATAGACATCTTCAAGCCAGTGCGCAAGgtatttacacacattgtctaggtttcaattttcatgagaagaagactaaaatataaatcttggcattcaatctgtacttgaatagctttagaaaaacttttaagaagatgtgataactttcttagcaaggttgatgggagtgttctattt

**PAM exon 21 and 100 bp of flanking intronic sequences**

PCR product: 300 bp

SNV: c.2332-2A>T

tttgttgttttgttgtgggctttgctttgtttttctgtcttttcatgaaggattcaagtaaaggctcaccactctaatgtgtagcttcttattttggcagCACTTTGATATGCCTCATGATATTGTTGCATCTGAAGATGGGACTGTGTACATTGGAGATGCTCATACCAACACCGTGTGGAAGTTCACCTTGACTGAGAgtatggttttcacagtattattgttcacattttccctcagtttgattgcttaaaagcatatgaaacaaaaacatttgcatttccttatctttttactgat
